# Linked electronic health records for research on a nationwide cohort including over 54 million people in England

**DOI:** 10.1101/2021.02.22.21252185

**Authors:** CVD-COVID-UK consortium, Manuscript drafting and revising, Angela Wood, Rachel Denholm, Sam Hollings, Jennifer Cooper, Samantha Ip, Venexia Walker, Spiros Denaxas, Ashley Akbari, Jonathan Sterne, Cathie Sudlow, Data wrangling, QA and analysis (including generating phenotype definitions), Angela Wood (chair), Rachel Denholm, Sam Hollings, Jennifer Cooper, Samantha Ip, Venexia Walker, Spiros Denaxas, Amitava Banerjee, William Whiteley, Figures/graphics, Alvina Lai, Consortium coordination (BHF Data Science Centre core team), Rouven Priedon, Cathie Sudlow, Lynn Morrice, Debbie Ringham, Public and patient advisory panel, Suzannah Power, Lynn Laidlaw, Michael Molete, John Walsh, NHS Digital (coordination, data management/provision, data access request and information governance support Trusted Research Environment support, Garry Coleman, Cath Day, Elizabeth Gaffney, Tim Gentry, Lisa Gray, Sam Hollings, Richard Irvine, Brian Roberts, Estelle Spence, Janet Waterhouse

## Abstract

**Objectives:** Describe a new England-wide electronic health record (EHR) resource enabling whole population research on Covid-19 and cardiovascular disease whilst ensuring data security and privacy and maintaining public trust.

**Design:** Cohort comprising linked person-level records from national healthcare settings for the English population accessible within NHS Digital’s new Trusted Research Environment.

**Setting:** EHRs from primary care, hospital episodes, death registry, Covid-19 laboratory test results and community dispensing data, with further enrichment planned from specialist intensive care, cardiovascular and Covid-19 vaccination data.

**Participants:** 54.4 million people alive on 1^st^ January 2020 and registered with an NHS general practitioner in England.

**Main measures of interest:** Confirmed and suspected Covid-19 diagnoses, exemplar cardiovascular conditions (incident stroke or transient ischaemic attack (TIA) and incident myocardial infarction (MI)) and all-cause mortality between 1^st^ January and 31^st^ October 2020.

**Results:** The linked cohort includes over 96% of the English population. By combining person-level data across national healthcare settings, data on age, sex and ethnicity are complete for over 95% of the population. Among 53.2M people with no prior diagnosis of stroke/TIA, 98,721 had an incident stroke/TIA, of which 30% were recorded only in primary care and 4% only in death registry records. Among 53.1M people with no prior history of MI, 62,966 had an incident MI, of which 8% were recorded only in primary care and 12% only in death records. A total of 959,067 people had a confirmed or suspected Covid-19 diagnosis (714,162 in primary care data, 126,349 in hospital admission records, 776,503 in Covid-19 laboratory test data and 48,433 participants in death registry records). While 58% of these were recorded in both primary care and Covid-19 laboratory test data, 15% and 18% respectively were recorded in only one.

**Conclusions:** This population-wide resource demonstrates the importance of linking person-level data across health settings to maximize completeness of key characteristics and to ascertain cardiovascular events and Covid-19 diagnoses. Although established initially to support research on Covid-19 and cardiovascular disease to benefit clinical care and public health and to inform health care policy, it can broaden further to enable a very wide range of research.

## INTRODUCTION

The Covid-19 pandemic has increased awareness of the importance of population-wide person-level electronic health record (EHR) data from a range of sources for examining, modelling and reporting disease trends to inform healthcare and public health policy^1^. Key benefits of research using such data on nationwide cohorts include: (i) generalisability of findings across all age groups, ethnicities, geographical locations and socioeconomic, health and personal characteristics and (ii) inclusion of very large numbers of people and events, enhancing the precision of findings and enabling a wide spectrum of novel research studies (e.g., characterising shapes of relationships between risk factors and disease or studying minority sub-populations and rare disease sub-types). Whilst EHRs for whole country cohorts for Wales, Scotland, Denmark and Sweden (populations approximately 3 to 10 million) have been used for research for several years,^2,3,4,5,6^ at the start of the COVID-19 pandemic, there was no access for bona fide researchers to national linked healthcare data across the population of England to enable critical research to support healthcare decisions and public health policy. There were two main reasons for this: there was no national collection of comprehensive, linkable primary care data; and there was no secure, privacy-protecting mechanism for researchers to access and conduct population-wide research using national datasets linked across different parts of the health data system (from primary care, hospitals, death registries, laboratories etc). EHR research in England to date has, therefore, not been able to take advantage of the statistical power of studying a population of almost 60 million people, while clinical, public health and policy insights have directly represented only a subset of the population. Hence, there remains a need for accessible, nationwide health data in England for research, whilst ensuring participant safety and maintaining public trust.

Motivated by the public health importance of fully understanding the relationship between Covid-19 and cardiovascular disease (CVD), the British Heart Foundation (BHF) Data Science Centre^7^ established the CVD-COVID-UK initiative^8^ to partner with NHS Digital^9^ in the development and secure provision for approved research of linked, nationally collated EHRs for the whole population of the UK. Here we describe key features of the new English component of this effort: a nationwide linked health data resource, provided within a new Trusted Research Environment (TRE) for England. We use descriptive analyses of the currently available data to illustrate the importance for whole population research studies of linking EHRs from across different health settings.

## METHODS

### Data resources

The newly established NHS Digital TRE for England provides secure, remote access for researchers to linked, person-level EHR data from national health settings. The data sources currently available include primary care data, hospital episodes (covering inpatient, outpatient, emergency department and critical care episodes), registered deaths (including cause of death), Covid-19 laboratory tests and community dispensed medicines (Table 1; CVD-COVID-UK Dataset dashboard;^10^ CVD-COVID-UK Dataset TRE asset in Health Data Research Innovation Gateway)^11^. Further incorporation of specialist intensive care, cardiovascular audit and Covid-19 vaccination data is planned in the near future. Datasets from each source include the same set of unique person-level master keys (or pseudo-identifiers) to enable linkage of peoples’ records between datasets.

**Table 1:**
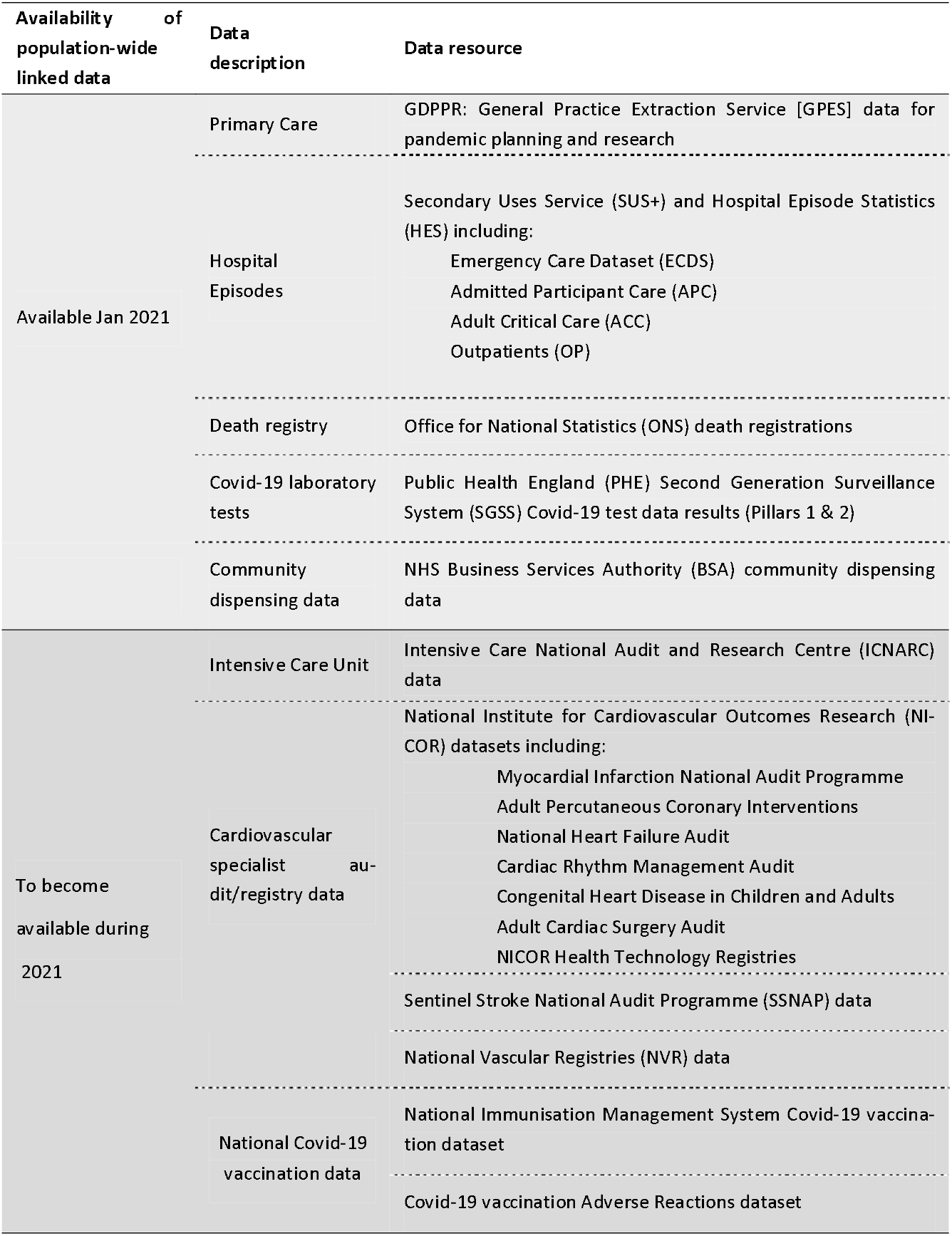
Overview of available and planned linked resources.

### Data linkage

Linkage between datasets is enabled by NHS Digital’s Master Person Service^12^, which uses a four-stage algorithm to match multiple records for each person from different clinical computer systems (e.g., hospitals and general practices) to a single unique identifier, the National Health Service (NHS) number representing a single person. The algorithm verifies and cross-checks the NHS numbers with associated demographic details including age, gender and postcode.^13^

### Data resource access – NHS Digital TRE for England

On behalf of the CVD-COVID-UK consortium, the BHF Data Science Centre requested access to the data sources via the NHS Digital online Data Access Request Service^14^ and received approval for the CVD-COVID-UK research programme (Ref no: DARS-NIC-381078-Y9C5K) following discussion with NHS Digital’s Independent Group Advising on the Release of Data (IGARD).^15^ A data sharing agreement with NHS Digital allows approved researchers based in UK research organisations (universities and NHS bodies) that jointly sign this agreement to access the data held within the NHS Digital TRE service for England.^16^ The BHF Data Science Centre coordinates an Approvals and Oversight Board (including representation from NHS Digital, participating research organisations and lay members) that ensures research projects undertaken fall within scope of the ethical and regulatory approvals for the CVD-COVID-UK consortium programme. The TRE provides secure storage and remote data access, avoiding the need for any person-level data to leave NHS Digital (Figure 1). An expanding suite of tools (currently including SQL, Python and R Studio) supports data management, visualization and analysis.

**Figure 1:**
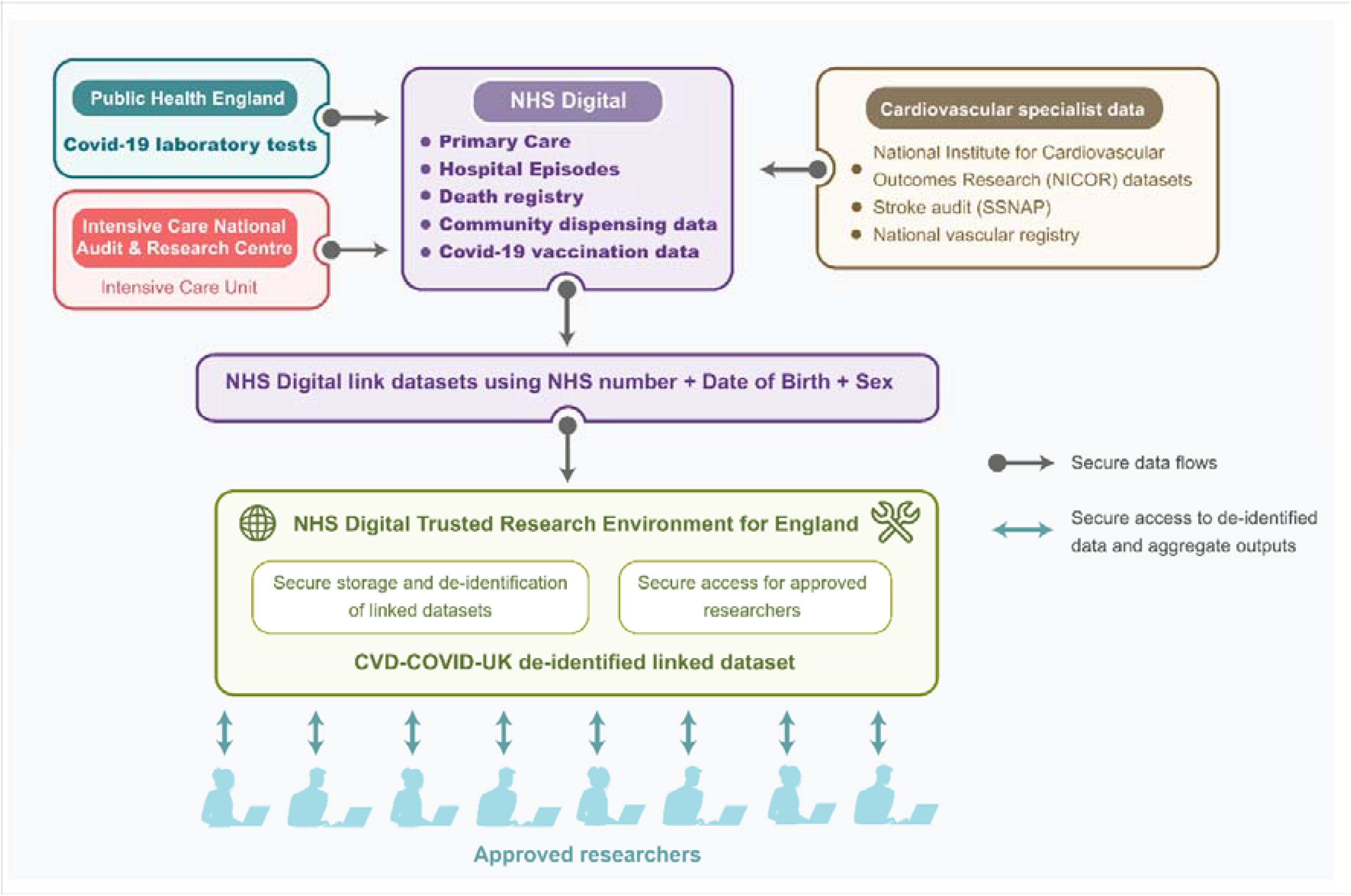
Overview of current (in bold text) and planned (regular text) data flows into the NHS Digital Trusted Research Environment for England.

### CVD-COVID-UK Consortium: Aims, Membership and Principles

The CVD-COVID-UK consortium aims to use analyses of UK population-wide linked EHR data to investigate: the effects of cardiovascular diseases, their risk factors and medications on susceptibility to and poor outcomes from Covid-19; the direct impact of SARS-CoV-2 infection on acute cardiovascular complications and longer-term cardiovascular risk; and the indirect impact of the pandemic on the presentation, diagnosis, management and outcomes of cardiovascular diseases.^8^ Lay summaries of approved projects are published on the consortium’s web page^8^. All consortium members (currently over 130 people from around 40 research or NHS organisations, including NHS data custodians) commit to: conducting research according to the ‘Five Safes’;^17^ an inclusive approach that enables additional researchers to join the consortium as the work evolves; and the open sharing of research protocols and analysis code (via the BHF Data Science Centre Github repository)^18^ and of phenotype code lists and algorithms (via the HDR UK Phenotype Library).^19^

### Data updates

The datasets within the TRE are updated regularly from NHS Digital’s internal systems (between daily and fortnightly depending on the dataset) and have a variable lag behind real time at the point of update (Table 2). The datasets are currently refreshed on a synchronized monthly schedule, but more frequent updates (e.g. daily or weekly) can be requested according to clinical, public health and health policy research needs.

**Table 2:**
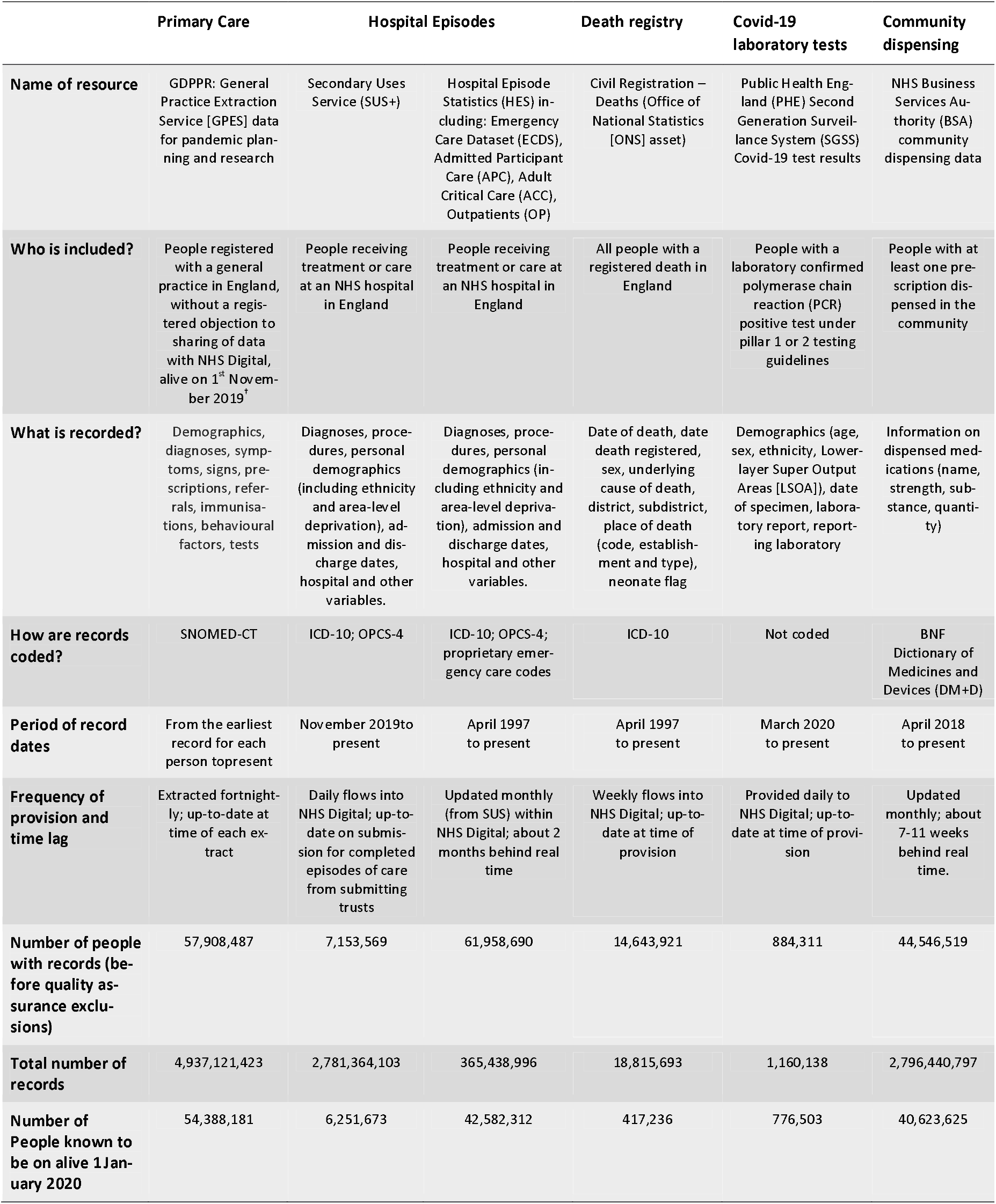
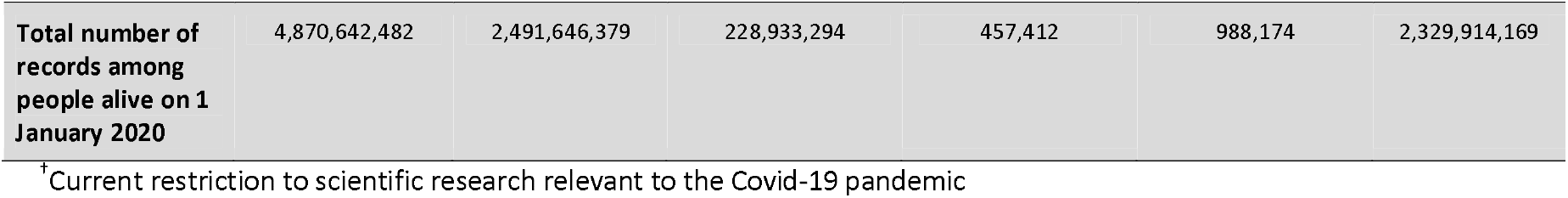
Key details of main data resources.

### Data security, privacy and confidentiality

The data within the TRE are de-identified (i.e., directly identifying data items, such as each person’s name, address, NHS number and exact date of birth, are removed) and pseudonymised (i.e., each unique person-specific NHS number is replaced with a non-identifying unique master key). Post-codes are replaced with lower layer super output areas which can be converted to indices of multiple deprivation.^20^ Further, NHS Digital operates a ‘safe outputs’ service: only summary, aggregate results can be extracted from the TRE by approved researchers, subject to approval through disclosure control processes and rules, following similar principles to those used by other established TREs, such as the Secure Anonymised Information Linkage (SAIL) Databank for Wales^21,22^ and the Scottish National Data Safe Haven.^23^ This ensures that no output that might be placed in the public domain contains information that could be used either on its own or in conjunction with other data to identify a person.

### Ethical approval

The North East - Newcastle & North Tyneside 2 Research Ethics Committee provided ethical approval for the CVD-COVID-UK research programme (REC number: 20/NE/0161).

### Patient and Public Involvement

The lay panel of the UK National Institutes for Health Research-BHF Cardiovascular Partnership reviews the CVD-COVID-UK programme every few months and provides feedback that informs ongoing and future research. In addition, lay people directly affected by cardiovascular disease are members of the consortium and its Approvals and Oversight Board, enabling co-generations of research ideas and providing valuable perspective and input on research proposals, lay summaries and research outputs.

### Derivation of participant characteristics and disease diagnoses

For descriptive analyses, we defined a linked cohort including all people in the primary care data known to be alive on 1^st^ January 2020, excluding those who had either died before or were born on or after that date (as recorded in the death registry and in the primary care records, respectively). We censored follow-up on 31^st^ October 2020, the latest record date common across the datasets. We defined eligible records within the hospital episodes, death registry and Covid-19 laboratory test results as those which could be linked by their unique master key to a person included in the primary care data.

We combined primary care and hospital episodes records (covering inpatient, outpatient, emergency department and critical care episodes) from before the index date of 1^st^ January 2020 to define key characteristics, including sex, age and ethnicity (categorised into White; Mixed; Asian and Asian British, Black and Black British and other ethnic groups). For each characteristic, we extracted the most recent record from the primary care data if available, otherwise we used the most recent record from the hospital episodes records. Characteristics were classified as “unknown” for people with no records. Using previously validated phenotypes from the CALIBER resource,^24^ we defined previous diagnoses of MI (yes/no), stroke/TIA (defined as ischaemic stroke, haemorrhagic stroke, unspecified stroke or TIA) (yes/no), diabetes (yes/no) and obesity (yes/no) from Systematized Nomenclature of Medicine Clinical Terms (SNOMED-CT) concept codes in the primary care data and from ICD-10 codes in the hospital episodes (main or secondary diagnostic code position in the admitted patient care component of the hospital episode statistics data) recorded prior to 1^st^ January 2020. For primary care phenotypes, we translated and expanded the phenotypes defined in Read Terms V2 to SNOMED-CT and cross referenced them with codes in the primary care dataset.^25^ Two clinicians independently reviewed all phenotype code lists and, where applicable, classified ICD-10 terms and SNOMED-CT concepts into prevalent or incident (**Supplementary Tables 1-4**).^19^

We ascertained people with a first-ever incident MI or stroke/TIA as those with no diagnosis of MI or stroke/TIA (as defined above) prior to 1^st^ January 2020 and with a diagnosis SNOMED-CT or ICD-10 code appearing in the primary care data, hospital episodes (main or secondary diagnostic code position in the admitted patient care component of the hospital episode statistics data) or death registry (underlying or contributing cause of death) between 1^st^ January and 31^st^ October 2020 (phenotype algorithms provided in **Supplementary Tables 1-2**).^19^

We ascertained people with a confirmed or suspected Covid-19 diagnosis as follows: (i) a positive PCR or antigen test from the Covid-19 laboratory test data, with specimen date on or before 31^st^ October 2020; or (ii) a Covid-19 diagnosis SNOMED-CT concept code appearing in the primary care data, with event date on or before 31^st^ October 2020; or (iii) a diagnosis ICD-10 code appearing in the hospital episodes (main or secondary diagnostic code position in the admitted patient care component of the hospital episode statistics), with admission date on or before 31^st^ October 2020 or (iv) death registration including a diagnosis ICD-10 code (as underlying or contributing cause), with date of death on or before 31^st^ October 2020. All Covid-19 phenotype definitions are provided in **Supplementary Table 5**.)^19^

We followed the RECORD guidance in preparing this manuscript (Annexe 2).^26^

## RESULTS

### Overview of data resources

**Table 2** provides an overview of the currently available primary care, hospital episodes, death registrations, Covid-19 test data and community dispensing data sources.

The primary care dataset includes healthcare information coded with SNOMED-CT concepts for all people registered with an English NHS general practice (excluding around 1.3 million people with a registered objection to their general practice records being provided to NHS Digital).^25^ It includes data from 98% of all English general practices across all relevant general practice computer system suppliers (TPP, EMIS, InPracticeSystems and Microtest) and holds approximately 4.9 billion records on 54.4 million people alive on 1^st^ January 2020 (over 96% of the total population of England based on the UK Office for National Statistics mid-2019 population estimate for England of 56,286,961).^27^ Around 34,000 SNOMED codes are included (over 90% of all those currently extracted for a wide range of purposes by NHS Digital’s GP Extraction Service), covering a broad range of diagnoses and procedures (from as far back in time as records exist) along with laboratory results, physical measurements, clinical referrals and prescriptions. Of note, while over 900,000 SNOMED codes are listed in UK and international releases, large numbers of these are either inactive or hardly used.

Administrative and clinical hospital episode data are available from both the Secondary Uses Service (SUS+) and Hospital Episode Statistics (HES) resources.^27^ These data include information on length of stay, diagnoses and procedures during hospital admissions as well as on outpatient, emergency department and critical care episodes. Diagnoses are coded with ICD-10 codes and procedures with Operating Procedure Codes (OPCS-4).^28^ The SUS+ resource contains raw data collected from NHS healthcare providers, representing the most up-to-date hospital episodes within NHS Digital (amongst hospitals making prompt and complete returns). These data are consolidated, validated and cleaned on a monthly basis to form the HES database.^29^ As a result, each month’s HES data become available about two months behind real time. Thereafter, a fixed update is produced for each full year of HES data. Amongst the 54.4 million people included in our linked cohort, the SUS+ dataset holds 2.5 billion records for 6.3 million people (from November 2019 onwards) and the HES dataset holds 0.2 billion records on 42.3 million people (from 1997 onwards).

Death registration data^11^ flow daily to NHS Digital from the Office for National Statistics (ONS) Civil Registration dataset, including date, cause (coded with ICD-10) and place of death, and are available historically from April 1997. Deaths in England should be registered within five days of the date of death, although registration of a death is delayed in some situations.^30,31^ Amongst the 54.4 million people in our linked cohort, 417,236 died on or before 31^st^ October 2020.

The Second Generation Surveillance System (SGSS)^11^ is the national laboratory reporting system used in England to capture routine laboratory data on mainly infectious diseases and antimicrobial resistance, including the SARS-CoV-2 virus. SGSS provides reports daily to NHS Digital on positive Covid-19 results (including the test date) fed directly from Pillar 1 pathology labs (i.e., established labs in hospitals for patients as well as NHS key workers), and indirectly from Pillar 2 labs (i.e., new, centralised, mostly privately-run labs, created specifically for Covid-19 testing for the wider population). In total 884,341 participants have at least one positive Covid-19 test recorded in the SGSS Covid-19 laboratory test dataset, of which 776,503 participants (88%) are linkable to the 54.4 million person cohort.

The community dispensing dataset, provided to NHS Digital monthly by the NHS Business Services Authority, contains person-level information on NHS primary care prescriptions dispensed by community pharmacists, appliance contractors and dispensing doctors in England, including the name and strength of medication coded from the British National Formulary (BNF) Dictionary of Medicines and Devices (DM+D).^32^ Amongst the 54.4 million person cohort, there are over 40.6 million with dispensed medications and approximately 2.3 billion records (from April 2018).

### Demographic characteristics and cardiovascular disease incidence

Characteristics of the linked cohort of 54.4 million people alive on 1^st^ January 2020 are shown in **Table 3**; on 1^st^ January 2020, 51% were female and 14% aged 70 years or older, with a mean age of 40.0 years for males and 41.6 years for females. By linking and combining person-level records from primary care and hospital episodes, ethnicity information is available for over 95% of people, among whom 63% have their ethnic group recorded in primary care and 92% in hospital episodes data (**Figure 2a**). A previous diagnosis of stroke/TIA or MI is recorded for 2.2% and 2.1% of people, respectively, while 7% and 8% people have a record indicating a previous diagnosis of diabetes and obesity, respectively. Among 53.2 million people with no prior diagnosis of stroke/TIA, 98,721 had a first-ever incident stroke/TIA between 1 January and 31^st^ October 2020, of which 30% were recorded only in primary care (i.e. not in hospital episodes or death registry data) and 4% only in death registry records (**Figure 2b**). Among 53.1M people with no prior MI, 62,966 had an incident MI during follow up, of which 8% were recorded only in primary care and 12% only in death registry records (**Figure 2c**).

**Table 3:**
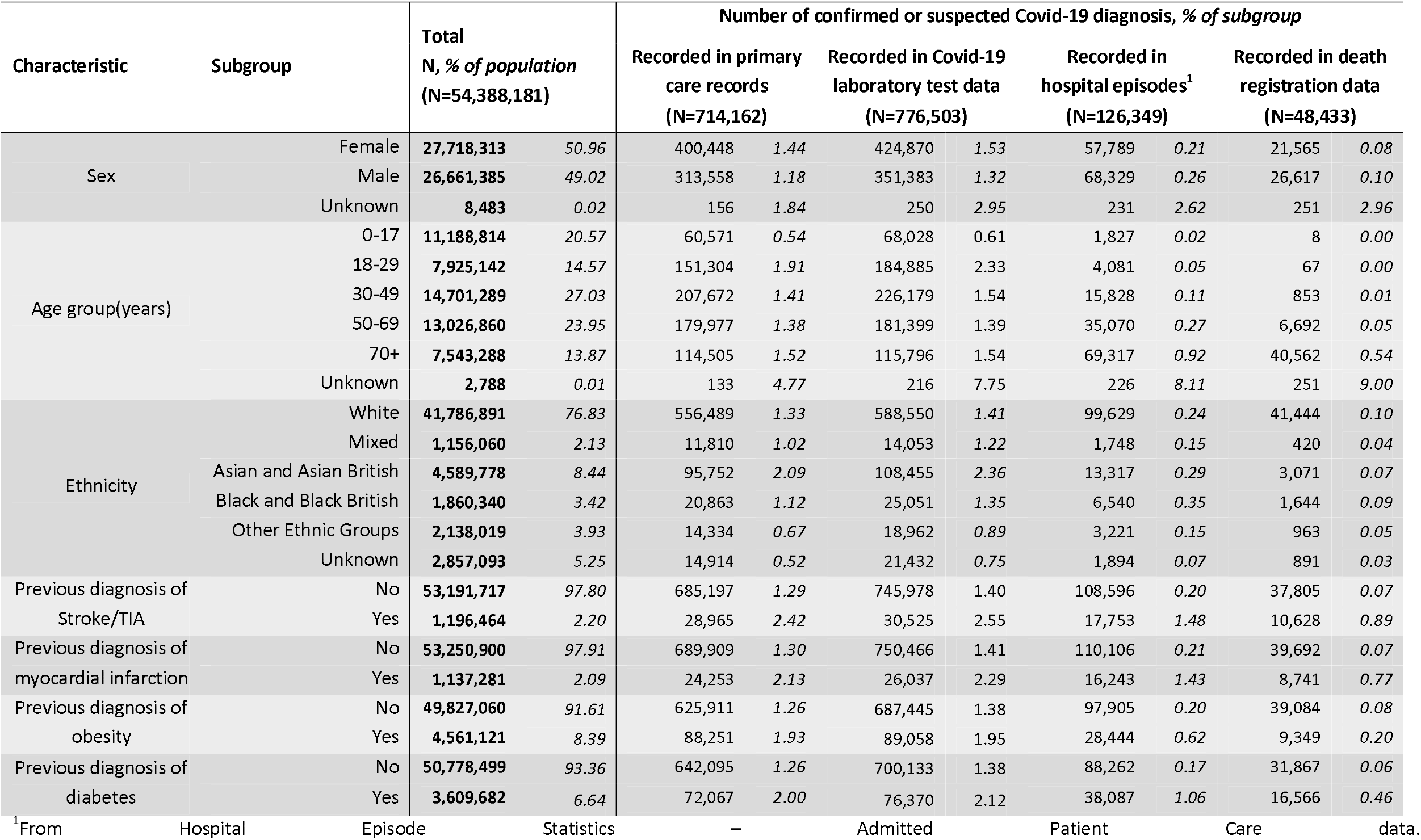
Characteristics of linked cohort and of people with a confirmed or suspected Covid-19 diagnosis, by data resource.

**Figure 2:**
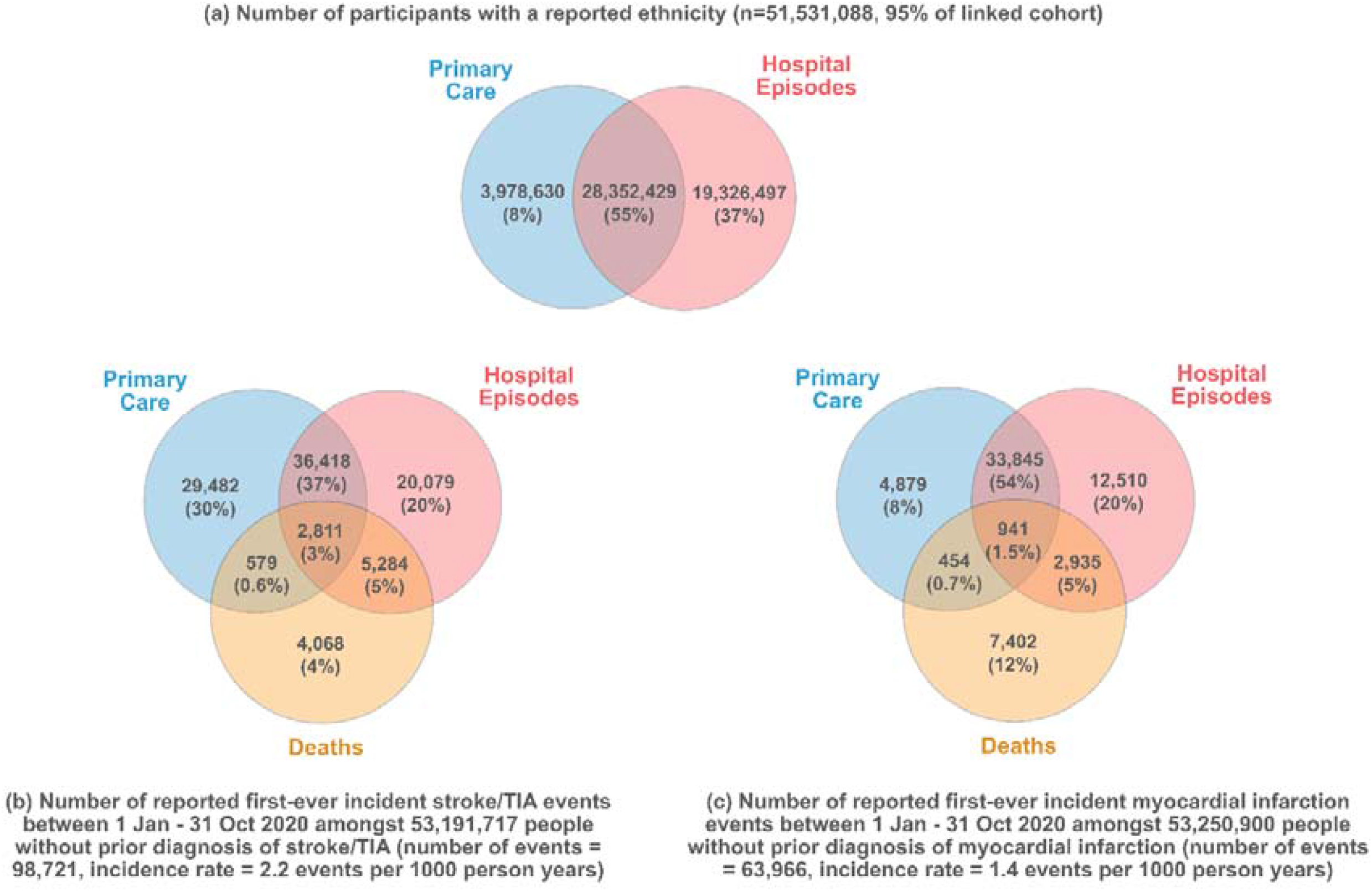
Data sources reporting person-level data on (a) ethnicity; (b) incident stroke/TIA (c) incident MI.

### Covid-19 diagnoses

Among people in the linked cohort, a total of 959,067 people had a confirmed or suspected Covid-19 diagnosis between 1^st^ January and 31^st^ October 2020 (714,162 in primary care data, 126,349 in hospital admission records, 776,503 in Covid-19 laboratory test data and 48,433 in death registry records). While 58% of these were recorded in both primary care and Covid-19 laboratory test data, 15% and 18% respectively were recorded in only one of these (**Figure 3**).

**Figure 3:**
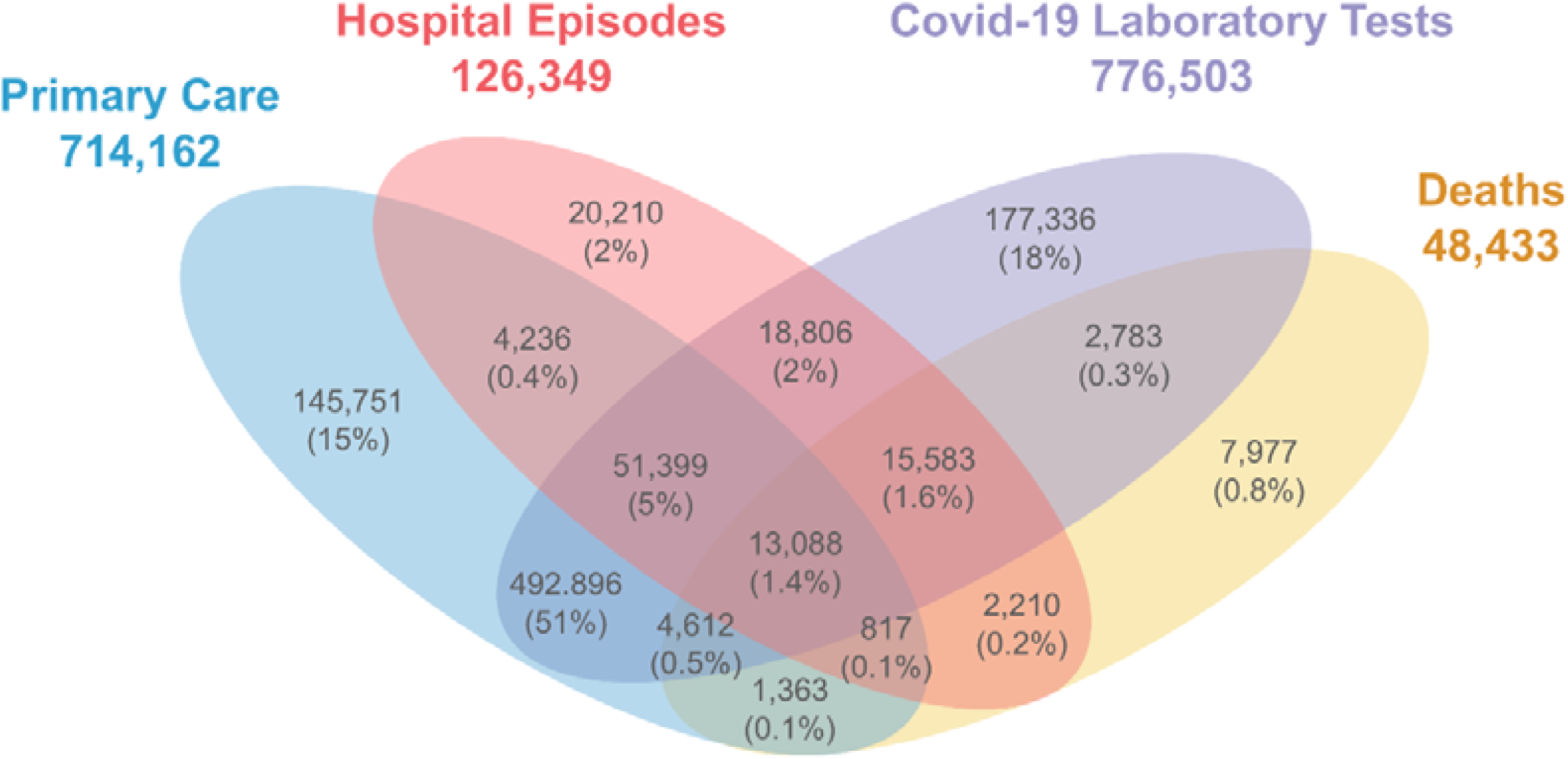
Data sources reporting person-level data on confirmed or suspected Covid-19 diagnoses between 1 Jan 2020 - 31 October 2020 (n=959,067). Numbers indicate distinct people with a confirmed or suspected Covid-19 diagnosis.

Whereas females are more likely to have a confirmed or suspected Covid-19 diagnosis in their primary care records (1.4% females versus 1.2% males) and in Covid-19 laboratory test data (1.5% females versus 1.3% males), they are less likely to have a Covid-19 diagnosis recorded in hospital episodes (0.21% females versus 0.26% males) or on death certificates (0.08% females versus 0.10% males). Older people are more likely to have a Covid-19 diagnosis from hospital episodes and death registrations, although young adults are more likely to have Covid-19 diagnoses recorded in Covid-19 laboratory test data and primary care. People with unknown age or sex are over 10 times more likely to have a Covid-19 diagnosis recorded in hospital episodes or on death certificates. A higher proportion of Asian and Asian British people have a Covid-19 diagnosis in primary care and in the Covid-19 laboratory tests in comparison with other ethnicities. However, such differences are not observed in information from hospital episodes or death certificates. People with a previous history of stroke/TIA, MI, obesity or diabetes are more likely to have a Covid-19 diagnosis recorded in all healthcare settings (**Table 3**).

When compared with the latest Public Health England reports of Covid-19 laboratory tests,^33^ Covid-19-related hospital admissions^34^ and deaths with Covid-19 on the death certificate,^35^ our linked cohort produces statistics which concord with relevant cumulative counts of Covid-19 cases (**Supplementary Table 6**).

## DISCUSSION

### Summary

We have described the development and key features of a novel linked EHR resource comprising a range of current and future planned linked datasets covering the entire population of England and forming part of wider UK-wide initiative to accelerate UK-wide research on Covid-19 and cardiovascular disease and beyond. We include descriptive analyses of a cohort of 54.4 million people alive at the start of 2020, including over 96% of the English population. The datasets described are already being accessed through the new NHS Digital TRE service for England to enable an expanding range of research projects via the BHF Data Science Centre’s CVD-COVID-UK consortium. Notably, combining person-level information across data sources delivers approximately 95% complete data on key characteristics including age, sex and ethnicity and is essential for identifying cardiovascular diseases of interest, such as stroke and myocardial infarction. Approximately 90% of people with a positive Covid-19 laboratory test have linkable primary care records, and enriching the Covid-19 laboratory test data with primary care, hospital episodes and death registry data enables ascertainment of approximately 20% additional confirmed or suspected Covid-19 cases.

Previously, research use of linked EHRs in England has been restricted to subsets of the population, according to the coverage of various data providers, including the individual primary care computer system suppliers (e.g., the Clinical Practice Research Datalink,^36^ The Health Improvement Network,^37^ QResearch^38^ and, more recently, OpenSafely^39^). As the national provider of information, data and IT systems for commissioners, analysts and clinicians in health and social care in England, NHS Digital handles larger volumes of health data than any other organisation globally and has extremely well developed and robust processes for maintaining data security and privacy. Alignment of the new Trusted Research Environment for England with NHS Digital’s systems therefore maximises security while minimising the need for transmission of large volumes of linked data to support population-scale research.

### Strengths and limitations

The currently available linked data assets comprise the world’s largest single population-based cohort available for research, which will be further enhanced as further datasets are added. The availability of primary care data linked to such a wide range of other data is unparalleled at this scale, while the resource is also making linked nationwide Covid-19 laboratory testing and community dispensing data available for research for the first time. Unsurprisingly, given the >96% coverage of the English population, the linked cohort represents the English population in terms of age, sex, ethnicity, and diabetes, when compared with UK Government England official statistics,^40–42^ includes the full distribution of general practices according to geographic location and size,^25^ and includes large enough numbers of people with different characteristics to support a diverse range of statistically well-powered research studies. For example, the cohort includes large numbers of: people in sub-groups typically under-represented in research (e.g., several tens of thousands in each of the ethnic minority subgroups); younger people for whom poor outcomes of Covid-19 are uncommon but nonetheless devastating (e.g., over 20 million under 30 years of age, among whom 75 deaths were recorded by 31^st^ October 2020); people experiencing the common exemplar cardiovascular outcomes of stroke/TIA and MI (many tens of thousands), suggesting substantial potential to support studies of the impact of Covid-19 on subtypes of stroke and MI as well as on a wide range of rare conditions.

The NHS Digital TRE for England ensures secure, privacy-protecting storage of and access to large volumes of data, while minimizing the expense and security risks of data travel. Provision of data in this way is enabling a broad programme of collaborative research, encompassing several projects, which would be challenging to justify under the data dissemination model but which meet the relevant ethics and data access requirements under the TRE model. Researchers from many different organisations have been able to gain rapid access to the linked datasets via the CVD-COVID-UK consortium and its data sharing agreement with NHS Digital, avoiding lengthy and costly processes for multiple separate organisational data-access approvals and agreements. The consortium is enabling collaboration amongst researchers from across the UK, with a wide range of expertise (including clinicians from many different specialist backgrounds, data managers, computer scientists, data wranglers, epidemiologists and biostatisticians). Further, it has encouraged productive interactions between researchers and NHS Digital staff (including project management, data management, data science and technical development teams), enabling joint approaches to developing the TRE service and to identifying and solving data provision and linkage challenges. The rich and diverse nature of this interdisciplinary collaboration supports clinically and methodologically informed data curation and analysis pipelines and will enhance the interpretation and clinical application of research outputs. Regular dataset updates ensure the contemporary relevance and dynamic nature of the data resource and will enable ongoing long-term follow-up of the whole population. The development of publicly shareable, validated phenotyping algorithms^19^ and analytic code^18^ will avoid duplication of effort by additional groups of researchers working with the same or similar datasets. Although developed with the initial intent of supporting the CVD-COVID-UK consortium research programme, establishing the NHS Digital TRE Service for England has wider benefits, given its clear potential to expand to support research more broadly beyond Covid-19 and beyond the cardiovascular domain. In addition, the work to establish the TRE has generated knowledge about linked EHR data and routes to their access across the UK health data science research community, benefiting other UK-wide initiatives, including: the UK-wide ISARIC study of the clinical characteristics of people hospitalized with Covid-19,^43^ collaborative efforts to address the determinants of Covid-19 susceptibility, severity and outcome through analyses of population-based cohorts with bio-samples linked to national EHRs^44,45^; the RECOVERY randomized trial of treatments for Covid-19^46^; the COG-UK Covid-19 viral sequencing study;^47^ and the UK Government Chief Scientific Adviser’s National Core Studies programme, established to coordinate the UK’s Covid-19 research response (in particular its underpinning Data and Connectivity theme led by Health Data Research UK).^48,49^

Nevertheless, there are some limitations: (i) it is not yet possible to bring external cohort studies or trials into the environment for linkage (although data can be linked to these through NHS Digital’s standard data dissemination route); (ii) the primary care data are currently restricted to a large subset of SNOMED codes and limited to people known to be alive from November 2019 onwards, although NHS Digital is currently enacting its plans to obtain a fully comprehensive primary care dataset, to be updated daily, which will become available during 2021 and will eventually replace the current primary care dataset; (iii) the TRE currently has a relatively limited range of services and analytical tools, although NHS Digital are committed to expanding these; (iv) the descriptive results presented here provide an overview of the available resources with illustrative examples but are not designed to inform reliable conclusions about the associations between patient characteristics and COVID-19 outcomes, as the analyses are unadjusted and so prone to confounding; (v) whilst some data quality checks have been performed before creating the linked cohort, future analyses may require additional checks to minimize influential errors, outliers and inconsistent records.

### Combining resources across the four nations of the UK

Similar, albeit not identical, EHR data resources are available in separate TREs provided by SAIL Databank for Wales, the National Data Safe Haven in Scotland and the Honest Broker Service in Northern Ireland. Due to differences in data structure and coding procedures between nations, we advocate the development of analysis plans which aim for maximum consistency but allow for nation-specific differences. Where appropriate, results of nation-specific analyses can be combined to produce results with UK-wide coverage. Such combined analyses will, increasingly, be able to take advantage of Health Data Research UK’s plans to provide the infrastructure, methods and tools to enable federation of analyses across TREs.^50^

### Conclusion

We describe the first-ever provision for research of linked nationwide EHR data for England and demonstrate the importance of linking person-level data from different health settings for defining exemplar cardiovascular disease outcomes, Covid-19 diagnoses and key characteristics. By covering almost the entire English population, the resource includes all age groups, ethnic geographic, and socioeconomic, health and personal characteristics and can enable statistically powerful population-scale research with very large numbers of outcomes. It is accessible by approved researchers through a secure TRE hosted by NHS Digital to support research on Covid-19 and cardiovascular disease and can expand to benefit other future research initiatives beyond Covid-19 and cardiovascular disease.

## Data Availability

Data are available for bona fide researchers accessible within the NHS Digital trusted Research Environment for England. Contact for information on how to join the CVD-COVID-UK consortium for access.

https://www.hdruk.ac.uk/wp-content/uploads/2021/02/210215-CVD-COVID-UK-TRE-Dataset-Dashboard_CLMS.pdf

https://web.www.healthdatagateway.org/dataset/7e5f0247-f033-4f98-aed3-3d7422b9dc6d

## ACKNOWLEDGEMENTS

The BHF Data Science Centre (BHF Grant no. SP/19/3/34678, awarded to Health Data Research UK) funded co-development (with NHS Digital) of the TRE, provision of linked datasets, data access, user software licences, computational usage and data management and wrangling support, with additional contributions from the HDR UK Data and Connectivity component of the UK Chief Scientific Adviser’s National Core Studies programme to coordinate national Covid-19 priority research. Consortium partner organisations funded the time of contributing data analysts, biostatisticians, epidemiologists and clinicians.

The results described are based on data from patients, collected by the NHS as part of their care and support. We would also like to acknowledge all data providers who make anonymised data available for research.

AA is supported by Health Data Research UK [HDR-9006] which receives its funding from the UK Medical Research Council, Engineering and Physical Sciences Research Council, Economic and Social Research Council, Department of Health and Social Care (England), Chief Scientist Office of the Scottish Government Health and Social Care Directorates, Health and Social Care Research and Development Division (Welsh Government), Public Health Agency (Northern Ireland), British Heart Foundation (BHF) and the Wellcome Trust; and Administrative Data Research UK which is funded by the Economic and Social Research Council [grant ES/S007393/1]. AB is supported by research funding from NIHR, British Medical Association, Astra-Zeneca and UK Research and Innovation. AB, AW and SD are part of the BigData@Heart Consortium, funded by the Innovative Medicines Initiative-2 Joint Undertaking under grant agreement No. 116074. AW and SI are supported by the BHF-Turing Cardiovascular Data Science Award (BCDSA\100005) and by core funding from: UK Medical Research Council (MR/L003120/1), British Heart Foundation (RG/13/13/30194; RG/18/13/33946) and NIHR Cambridge Biomedical Research Centre (BRC-1215-20014). JC, JS and RD are supported by the HDRUK South West Better Care Partnership and NIHR Bristol Biomedical Research Centre. SD is supported by Health Data Research UK London, which receives its funding from Health Data Research UK Ltd funded by the UK Medical Research Council, Engineering and Physical Sciences Research Council, Economic and Social Research Council, Department of Health and Social Care (England), Chief Scientist Office of the Scottish Government Health and Social Care Directorates, Health and Social Care Research and Development Division (Welsh Government), Public Health Agency (Northern Ireland), British Heart Foundation, and the Wellcome Trust; Alan Turing Fellowship (EP/N510129/1); National Institute for Health Research (NIHR) Biomedical Research Centre (BRC) at University College London Hospital NHS Trust (UCLH). VW is supported by the University of Bristol Medical Research Council Integrative Epidemiology Unit (MC_UU_00011/4). WW is supported by a Scottish Senior Clinical Fellowship, CSO (SCAF/17/01)

The views expressed are those of the author(s) and not necessarily those of the organisations listed.

---

**Annexe 1:**
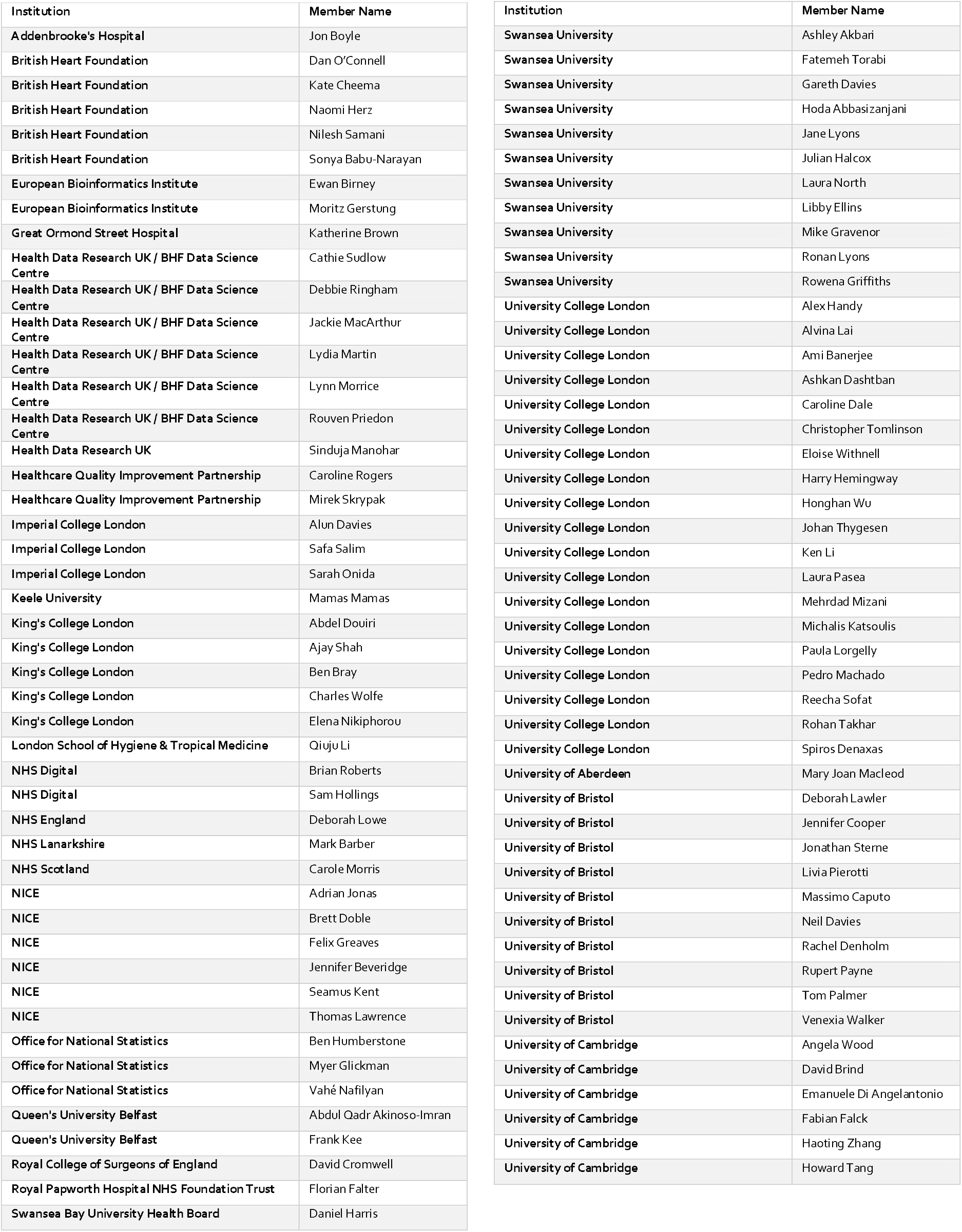
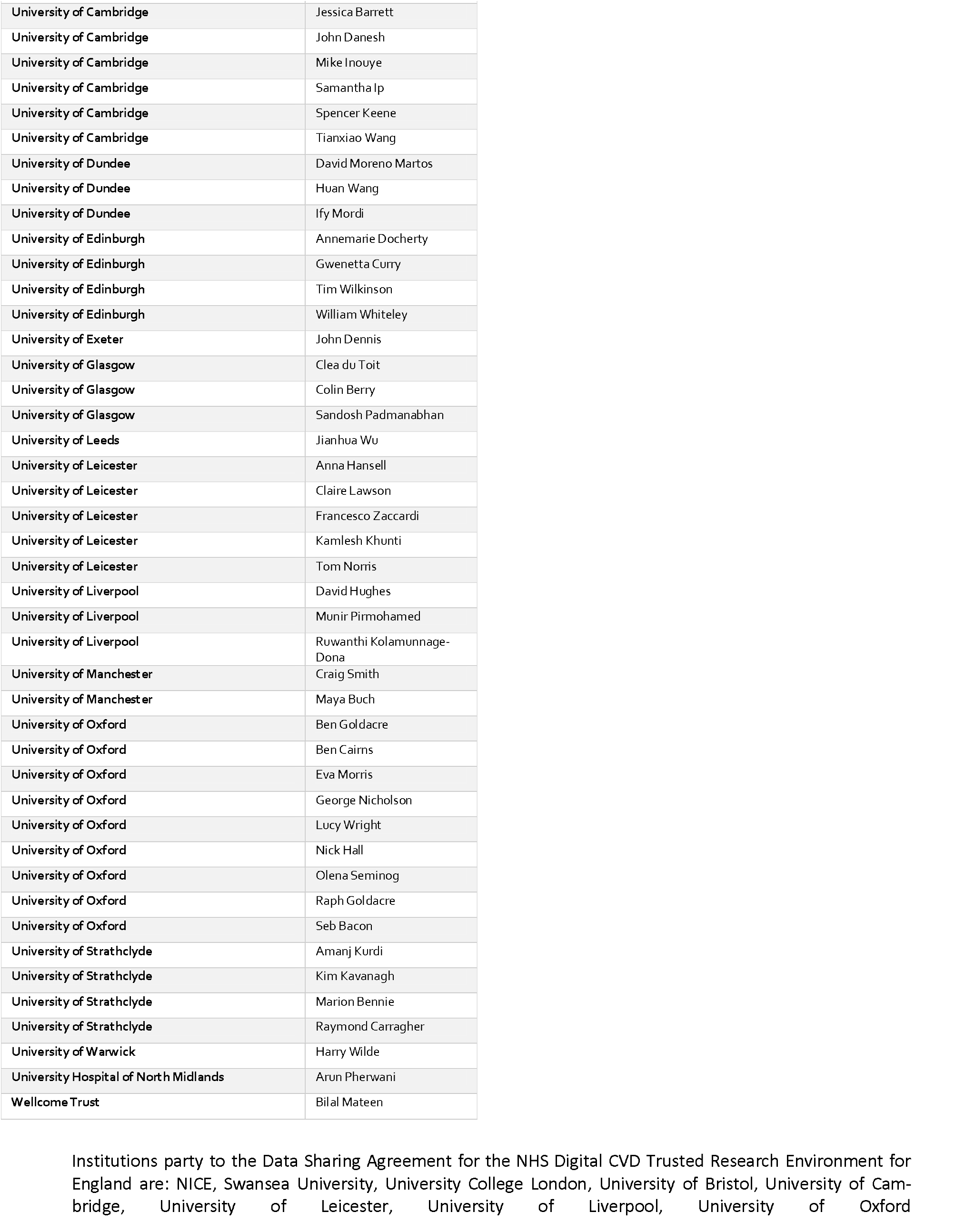
CVD-COVID-UK consortium members.

**Annexe 2:** CVD-COVID-UK RECORD statement - checklist of items, extended from the STROBE statement, that should be reported in observational studies using routinely collected health data.

|  | Item No. | STROBE items | Location in manuscript where items are reported | RECORD items | Location in manuscript where items are reported |
| --- | --- | --- | --- | --- | --- |
| **Title and abstract** |  |  |  |  |  |
|  | 1 | (a) Indicate the study's design with a commonly used term in the title or the abstract (b) Provide in the abstract an informative and balanced summary of what was done and what was found | Title and abstract | RECORD 1.1: The type of data used should be specified in the title or abstract. When possible, the name of the databases used should be included.<br>RECORD 1.2: If applicable, the geographic region and timeframe within which the study took place should be reported in the title or abstract.<br>RECORD 1.3: If linkage between databases was conducted for the study, this should be clearly stated in the title or abstract. | Title and abstract<br>Title and abstract<br>Title and abstract |
| **Introduction** |  |  |  |  |  |
| Background rationale | 2 | Explain the scientific background and rationale for the investigation being reported | Page 4 |  |  |
| Objectives | 3 | State specific objectives, including any prespecified hypotheses | Page 4 |  |  |
| **Methods** |  |  |  |  |  |
| Study Design | 4 | Present key elements of study design early in the paper | Pages 6 and 7<br>Figure 1, Table 1 |  |  |
| Setting | 5 | Describe the setting, locations, and relevant dates, including periods of recruitment, exposure, follow-up, and data collection | Pages 6 and 7<br>Table 2 |  |  |
| Participants | 6 | (a) *Cohort study* - Give the eligibility criteria, and the sources and methods of selection of participants. Describe methods of follow-up<br>*Case-control study* - Give the eligibility criteria, and the sources and methods of case ascertainment and control selection. Give the rationale for the choice of cases and controls<br>*Cross-sectional study* - Give the eligibility criteria, and the sources and methods of selection of participants<br>(b) *Cohort study* - For matched studies, give matching criteria and number of exposed and unexposed<br>*Case-control study* - For matched studies, give matching criteria and the number of controls per case | Pages 8 and 9, Supplementary Tables 1-5 | RECORD 6.1: The methods of study population selection (such as codes or algorithms used to identify subjects) should be listed in detail. If this is not possible, an explanation should be provided.<br>RECORD 6.2: Any validation studies of the codes or algorithms used to select the population should be referenced. If validation was conducted for this study and not published elsewhere, detailed methods and results should be provided.<br>RECORD 6.3: If the study involved linkage of databases, consider use of a flow diagram or other graphical display to demonstrate the data linkage process, including the number of individuals with linked data at each stage. | Page 9, Supplementary Tables 1-5<br>Page 9<br>Table 2 |
| Variables | 7 | Clearly define all outcomes, exposures, predictors, potential confounders, and effect modifiers. Give diagnostic criteria, if applicable. | Pages 8 and 9 | RECORD 7.1: A complete list of codes and algorithms used to classify exposures, outcomes, confounders, and effect modifiers should be provided. If these cannot be reported, an explanation should be provided. | Pages 8 and 9, Supplementary Tables 1-5 |
| Data sources/<br>measurement | 8 | For each variable of interest, give sources of data and details of methods of assessment (measurement).<br>Describe comparability of assessment methods if there is more than one group | Pages 8 and 9 |  |  |
| Bias | 9 | Describe any efforts to address potential sources of bias | Not applicable |  |  |
| Study size | 10 | Explain how the study size was arrived at | Page 10, Table 2 |  |  |
| Quantitative variables | 11 | Explain how quantitative variables were handled in the analyses. If applicable, describe which groupings were chosen, and why | Not applicable |  |  |
| Statistical methods | 12 | (a) Describe all statistical methods, including those used to control for confounding<br>(b) Describe any methods used to examine subgroups and interactions<br>(c) Explain how missing data were addressed<br>(d) *Cohort study* - If applicable, explain how loss to follow-up was addressed<br>*Case-control study* - If applicable, explain how matching of cases and controls was addressed<br>*Cross-sectional study* - If applicable, describe analytical methods taking account of sampling strategy<br>(e) Describe any sensitivity analyses | Not applicable |  |  |
| Data access and cleaning methods |  | .. |  | RECORD 12.1: Authors should describe the extent to which the investigators had access to the database population used to create the study population.<br>RECORD 12.2: Authors should provide information on the data cleaning methods used in the study. | Pages 7, 8 and 9<br>Page 8 |
| Linkage |  | .. |  | RECORD 12.3: State whether the study included person-level, institutional-level, or other data linkage across two or more databases. The methods of linkage and methods of linkage quality evaluation should be provided. | Page 6 |
| **Results** |  |  |  |  |  |
| Participants | 13 | (a) Report the numbers of individuals at each stage of the study (*e.g.*, numbers potentially eligible, examined for eligibility, confirmed eligible, included in the study, completing follow-up, and analysed)<br>(b) Give reasons for non-participation at each stage.<br>(c) Consider use of a flow diagram | Pages 10 and 11<br>Table 2<br>Pages 10 and 11<br>In Table 2 | RECORD 13.1: Describe in detail the selection of the persons included in the study (*i.e.*, study population selection) including filtering based on data quality, data availability and linkage. The selection of included persons can be described in the text and/or by means of the study flow diagram. | Pages 10 and 11<br>Table 2 |
| Descriptive data | 14 | (a) Give characteristics of study participants (*e.g.*, demographic, clinical, social) and information on exposures and potential confounders<br>(b) Indicate the number of participants with missing data for each variable of interest | Table 3<br>Table 3 |  |  |
|  |  | (c) *Cohort study* - summarise follow-up time (e.g., average and total amount) | Page 11, Figure 2 |  |  |
| Outcome data | 15 | *Cohort study* - Report numbers of outcome events or summary measures over time<br>*Case-control study* - Report numbers in each exposure category, or summary measures of exposure<br>*Cross-sectional study* - Report numbers of outcome events or summary measures | Table 3, Figure 2, Figure 3 |  |  |
| Main results | 16 | (a) Give unadjusted estimates and, if applicable, confounder-adjusted estimates and their precision (e.g., 95% confidence interval). Make clear which confounders were adjusted for and why they were included<br>(b) Report category boundaries when continuous variables were categorized<br>(c) If relevant, consider translating estimates of relative risk into absolute risk for a meaningful time period | Table 3<br>Table 3<br>NA |  |  |
| Other analyses | 17 | Report other analyses done—e.g., analyses of subgroups and interactions, and sensitivity analyses | Page 12<br>Table 3<br>Supplementary Table 6 |  |  |
| **Discussion** |  |  |  |  |  |
| Key results | 18 | Summarise key results with reference to study objectives | Page 13 |  |  |
| Limitations | 19 | Discuss limitations of the study, taking into account sources of potential bias or imprecision. Discuss both direction and magnitude of any potential bias | Page 15 | RECORD 19.1: Discuss the implications of using data that were not created or collected to answer the specific research question(s). Include discussion of misclassification bias, unmeasured confounding, missing data, and changing eligibility over time, as they pertain to the study being reported. | Page 15 |
| Interpretation | 20 | Give a cautious overall interpretation of results considering objectives, limitations, multiplicity of analyses, results from similar studies, and other relevant evidence | Page 16 |  |  |
| Generalisability | 21 | Discuss the generalisability (external validity) of the study results | Page 14, 15 |  |  |
| **Other Information** |  |  |  |  |  |
| Funding | 22 | Give the source of funding and the role of the funders for the present study and, if applicable, for the original study on which the present article is based | In Acknowledgements |  |  |
| Accessibility of protocol, raw data, and programming code |  | .. |  | RECORD 22.1: Authors should provide information on how to access any supplemental information such as the study protocol, raw data, or programming code. | Page 6 |

\*Reference: Benchimol EI, Smeeth L, Guttman A, Harron K, Moher D, Petersen I, Sørensen HT, von Elm E, Langan SM, the RECORD Working Committee. The REporting of studies Conducted using Observational Routinely-collected health Data (RECORD) Statement. *PLoS Medicine* 2015; 2:e1001885.
\*Checklist is protected under Creative Commons Attribution (CC BY) license.

## Notes

### Competing Interest Statement

The authors have declared no competing interest.

### Clinical Trial

Registered on the Health Data Research UK Innovation Gateway:
CVD-COVID-UK TRE asset in Health Data Research Innovation Gateway [Internet]. [cited 2021 Feb 18]. Available from: https://web.www.healthdatagateway.org/dataset/7e5f0247-f033-4f98-aed3-3d7422b9dc6d

### Clinical Protocols

https://github.com/BHFDSC

https://portal.caliberresearch.org/collections/bhf-data-science-centre

### Summary of Updates

Typo in discussion corrected: 200,000 people under 30 replaced with 20 million

