## Supplementary material for "Linked electronic health records for research on a nationwide cohort including over 54 million people in England"

**Supplementary Material**

**Supplementary Table 1: Codelists for Myocardial Infarction (see https://portal.caliberresearch.org/collections/bhf-data-science-centre)**

| **Code type** | **Code** | **Descriptions** |
| --- | --- | --- |
| ICD10 codes | I252 | Old myocardial infarction (History of MI) |
|  | I21 | Acute myocardial infarction (Acute MI not further specified) |
|  | I22 | Subsequent myocardial infarction |
|  | I23 | Certain current complications following acute myocardial infarction (Complications of MI) |
|  | I241 | Dressler's syndrome (complications of MI) |
| SNOMED CONCEPT IDS | 1755008 | Old myocardial infarction |
|  | 10273003 | Acute infarction of papillary muscle |
|  | 15990001 | Acute myocardial infarction of posterolateral wall |
|  | 22298006 | Myocardial infarction |
|  | 42531007 | Microinfarct of heart |
|  | 52035003 | Acute anteroapical myocardial infarction |
|  | 54329005 | Acute myocardial infarction of anterior wall |
|  | 57054005 | Acute myocardial infarction |
|  | 58612006 | Acute myocardial infarction of lateral wall |
|  | 62695002 | Acute anteroseptal myocardial infarction |
|  | 65547006 | Acute myocardial infarction of inferolateral wall |
|  | 66189004 | Postmyocardial infarction syndrome |
|  | 70211005 | Acute myocardial infarction of anterolateral wall |
|  | 70422006 | Acute subendocardial infarction |
|  | 71023004 | Pericarditis secondary to acute myocardial infarction |
|  | 73795002 | Acute myocardial infarction of inferior wall |
|  | 76593002 | Acute myocardial infarction of inferoposterior wall |
|  | 79009004 | Acute myocardial infarction of septum |
|  | 91335003 | Mural thrombus of heart |
|  | 113155009 | Myocardial infarction education |
|  | 129574000 | Postoperative myocardial infarction |
|  | 161502000 | H/O: myocardial infarct at less than 60 |
|  | 161503005 | H/O: myocardial infarct at greater than 60 |
|  | 164865005 | ECG: myocardial infarction |
|  | 194798004 | Acute anteroapical infarction |
|  | 194802003 | True posterior myocardial infarction |
|  | 194809007 | Acute atrial infarction |
|  | 194856005 | Subsequent myocardial infarction |
|  | 194857001 | Subsequent myocardial infarction of anterior wall |
|  | 194858006 | Subsequent myocardial infarction of inferior wall |
|  | 194861007 | Certain current complications following acute myocardial infarction |
|  | 194862000 | Haemopericardium as current complication following acute myocardial infarction |
|  | 194863005 | Atrial septal defect as current complication following acute myocardial infarction |
|  | 194865003 | Rupture of cardiac wall without hemopericardium as current complication following acute myocardial infarction |
|  | 194866002 | Rupture of chordae tendinae as current complication following acute myocardial infarction |
|  | 194867006 | Rupture of papillary muscle as current complication following acute myocardial infarction |
|  | 194868001 | Thrombosis of atrium, auricular appendage, and ventricle as current complications following acute myocardial infarction |
|  | 233838001 | Acute posterior myocardial infarction |
|  | 233843008 | Silent myocardial infarction |
|  | 233846000 | Ventricular septal defect as current complication following acute myocardial infarction |
|  | 233847009 | Cardiac rupture after acute myocardial infarction |
|  | 233860003 | Post-infarction mitral papillary muscle rupture |
|  | 233885007 | Post-infarction pericarditis |
|  | 233889001 | Post-infarction hemopericardium |
|  | 233929004 | Post-infarction mural thrombus |
|  | 266897007 | Family history of myocardial infarction |
|  | 267390005 | Post-infarction hypopituitarism |
|  | 304914007 | Acute Q wave myocardial infarction |
|  | 307140009 | Acute non-Q wave infarction |
|  | 308065005 | H/O: Myocardial infarction in last year |
|  | 311792005 | Postoperative transmural myocardial infarction of anterior wall |
|  | 311793000 | Postoperative transmural myocardial infarction of inferior wall |
|  | 311796008 | Postoperative subendocardial myocardial infarction |
|  | 314116003 | Post infarct angina |
|  | 315287002 | Diabetes mellitus insulin-glucose infusion in acute myocardial infarction |
|  | 371068009 | Myocardial infarction with complication (disorder) |
|  | 398274000 | Coronary artery thrombosis (disorder) |
|  | 399211009 | History of - myocardial infarction (context-dependent category) |
|  | 401303003 | Acute ST segment elevation myocardial infarction (disorder) |
|  | 401314000 | Acute non-ST segment elevation myocardial infarction (disorder) |
|  | 428752002 | Recent myocardial infarction (situation) |
|  | 461000119108 | History of MI less than 8 weeks |
|  | 698593009 | History of non-ST segment elevation myocardial infarction (situation) |
|  | 703330009 | Mitral valve regurgitation due to acute myocardial infarction with papillary muscle and chordal rupture |
|  | 703328007 | Mitral valve regurgitation due to acute myocardial infarction without papillary muscle and chordal rupture |
|  | 703326006 | Mitral regurgitation due to acute myocardial infarction |
|  | 103011000119106 | Coronary arteriosclerosis in patient with history of previous myocardial infarction (situation) |
|  | 285721000119104 | History of acute ST segment elevation myocardial infarction (situation) |
|  | 15960981000119105 | Mural thrombus of left ventricle following acute myocardial infarction (disorder) |
|  | 723858002 | Ventricular aneurysm as current complication following acute myocardial infarction (disorder) |
|  | 723859005 | Pulmonary embolism as current complication following acute myocardial infarction (disorder) |
|  | 723860000 | Arrhythmia as current complication following acute myocardial infarction (disorder) |
|  | 723861001 | Cardiogenic shock unrelated to mechanical complications as current complication following acute myocardial infarction (disorder) |
|  | 736978009 | Mural thrombus of right ventricle following acute myocardial infarction (disorder) |
|  | 1077002 | Septal infarction by electrocardiogram (finding) |
|  | 1089431000000107 | Postoperative nontransmural myocardial infarction (disorder) |
|  | 1089451000000100 | Acute nontransmural myocardial infarction (disorder) |
|  | 15712881000119105 | Acute ST segment elevation myocardial infarction of anterolateral wall (disorder) |
|  | 15712921000119103 | Acute ST segment elevation myocardial infarction of lateral wall (disorder) |
|  | 15712961000119108 | Acute ST segment elevation myocardial infarction of anteroseptal wall (disorder) |
|  | 15713121000119105 | Acute ST segment elevation myocardial infarction due to right coronary artery occlusion (disorder) |
|  | 15713161000119100 | Acute ST segment elevation myocardial infarction of septum (disorder) |
|  | 17531000119105 | Acute myocardial infarction due to left coronary artery occlusion (disorder) |
|  | 233825009 | Acute Q wave infarction - anteroseptal (disorder) |
|  | 233826005 | Acute non-Q wave infarction - anteroseptal (disorder) |
|  | 233832000 | Acute non-Q wave infarction - inferolateral (disorder) |
|  | 285981000119103 | Acute ST segment elevation myocardial infarction involving left anterior descending coronary artery (disorder) |
|  | 285991000119100 | Acute ST segment elevation myocardial infarction involving left main coronary artery (disorder) |
|  | 394659003 | Acute coronary syndrome (disorder) |
|  | 394710008 | First myocardial infarction (disorder) |
|  | 413444003 | Acute myocardial ischemia (disorder) |
|  | 428196007 | Mixed myocardial ischemia and infarction (disorder) |
|  | 429391004 | New myocardial infarction compared to prior study (finding) |
|  | 59063002 | Acute myocardial infarction of apical-lateral wall (disorder) |
|  | 63670007 | Myocardial imaging for infarct, planar technique (procedure) |
|  | 703165004 | Acute ST segment elevation myocardial infarction of anterior wall involving right ventricle (disorder) |
|  | 703210007 | Subsequent ST segment elevation myocardial infarction of anterior wall (disorder) |
|  | 703213009 | Acute ST segment elevation myocardial infarction of inferior wall (disorder) |
|  | 703251009 | Acute myocardial infarction of inferior wall involving right ventricle (disorder) |
|  | 703252002 | Acute myocardial infarction of anterior wall involving right ventricle (disorder) |
|  | 1089441000000103 | Postoperative transmural myocardial infarction (disorder) |
|  | 1089471000000109 | Acute transmural myocardial infarction (disorder) |
|  | 12238111000119106 | Acute ST segment elevation myocardial infarction of inferolateral wall (disorder) |
|  | 12238151000119107 | Acute ST segment elevation myocardial infarction of inferoposterior wall (disorder) |
|  | 15712841000119100 | Acute ST segment elevation myocardial infarction of posterolateral wall (disorder) |
|  | 15713041000119103 | Acute ST segment elevation myocardial infarction of posterior wall (disorder) |
|  | 15713081000119108 | Acute ST segment elevation myocardial infarction due to left coronary artery occlusion (disorder) |
|  | 15713201000119105 | Acute ST segment elevation myocardial infarction of posterobasal wall (disorder) |
|  | 23311000119105 | Acute myocardial infarction due to right coronary artery occlusion (disorder) |
|  | 233827001 | Acute Q wave infarction - anterolateral (disorder) |
|  | 233828006 | Acute non-Q wave infarction - anterolateral (disorder) |
|  | 233829003 | Acute Q wave infarction - inferior (disorder) |
|  | 233830008 | Acute non-Q wave infarction - inferior (disorder) |
|  | 233831007 | Acute Q wave infarction - inferolateral (disorder) |
|  | 233833005 | Acute Q wave infarction - lateral (disorder) |
|  | 233834004 | Acute non-Q wave infarction - lateral (disorder) |
|  | 233835003 | Acute widespread myocardial infarction (disorder) |
|  | 233836002 | Acute Q wave infarction - widespread (disorder) |
|  | 233837006 | Acute non-Q wave infarction - widespread (disorder) |
|  | 282006 | Acute myocardial infarction of basal-lateral wall (disorder) |
|  | 30277009 | Acute myocardial infarction with rupture of ventricle (disorder) |
|  | 314207007 | Non-Q wave myocardial infarction (disorder) |
|  | 41466009 | Myocardial imaging for infarct with ejection fraction, first pass technique (procedure) |
|  | 418044006 | Myocardial infarction in recovery phase (disorder) |
|  | 64627002 | Acute myocardial infarction of high lateral wall (disorder) |
|  | 703164000 | Acute ST segment elevation myocardial infarction of anterior wall (disorder) |
|  | 703209002 | Subsequent ST segment elevation myocardial infarction of inferior wall (disorder) |
|  | 703211006 | Subsequent ST segment elevation myocardial infarction (disorder) |
|  | 703212004 | Acute myocardial infarction during procedure (disorder) |
|  | 703253007 | Acute ST segment elevation myocardial infarction of inferior wall involving right ventricle (disorder) |
|  | 703360004 | Subsequent non-ST segment elevation myocardial infarction (disorder) |
|  | 70998009 | Acute myocardial infarction of posterobasal wall (disorder) |

**Supplementary Table 2a: Codelists for Ischaemic Stroke (see https://portal.caliberresearch.org/collections/bhf-data-science-centre)**

| **Code type** | **Code** | **Descriptions** |
| --- | --- | --- |
| **ICD10 codes** | I63.0 | Cerebral infarction due to thrombosis of precerebral arteries |
|  | I63.1 | Cerebral infarction due to embolism of precerebral arteries |
|  | I63.2 | Cerebral infarction due to unspecified occlusion or stenosis of precerebral arteries |
|  | I63.3 | Cerebral infarction due to thrombosis of cerebral arteries |
|  | I63.4 | Cerebral infarction due to embolism of cerebral arteries |
|  | I63.5 | Cerebral infarction due to unspecified occlusion or stenosis of cerebral arteries |
|  | I63.8 | Other cerebral infarction |
|  | I63.9 | Cerebral infarction, unspecified |
|  | I69.3 | Sequelae of cerebral infarction |
| SNOMED CONCEPT IDS | 71444005 | Cerebral thrombosis |
|  | 75543006 | Cerebral embolism |
|  | 78569004 | Posterior inferior cerebellar artery syndrome |
|  | 95457000 | Brain stem infarction |
|  | 95460007 | Cerebellar infarction |
|  | 195185009 | Cerebral infarct due to thrombosis of precerebral arteries |
|  | 195186005 | Cerebral infarction due to embolism of precerebral arteries |
|  | 195189003 | Cerebral infarction due to thrombosis of cerebral arteries |
|  | 195190007 | Cerebral infarction due to embolism of cerebral arteries |
|  | 195200006 | Carotid artery syndrome hemispheric |
|  | 195201005 | Multiple and bilateral precerebral artery syndromes |
|  | 195209007 | Middle cerebral artery syndrome |
|  | 195210002 | Anterior cerebral artery syndrome |
|  | 195211003 | Posterior cerebral artery syndrome |
|  | 195243003 | Sequelae of cerebral infarction |
|  | 230691006 | CVA - cerebrovascular accident due to cerebral artery occlusion |
|  | 230692004 | Infarction - precerebral |
|  | 230698000 | Lacunar infarction |
|  | 230699008 | Pure motor lacunar syndrome |
|  | 230700009 | Pure sensory lacunar infarction |
|  | 266253001 | Precerebral arterial occlusion |
|  | 307766002 | Left sided cerebral infarction |
|  | 307767006 | Right sided cerebral infarction |
|  | 373606000 | Occlusive stroke (disorder) |
|  | 413102000 | Infarction of basal ganglia (disorder) |
|  | 422504002 | Ischemic stroke (disorder) |
|  | 432504007 | Cerebral infarction (disorder) |
|  | 441526008 | Infarct of cerebrum due to iatrogenic cerebrovascular accident |
|  | 723083001 | Late effects of cerebral ischemic stroke (disorder) |
|  | 724424009 | Cerebral ischemic stroke due to small artery occlusion (disorder) |
|  | 724425005 | Cerebral ischemic stroke due to intracranial large artery atherosclerosis (disorder) |
|  | 724426006 | Cerebral ischemic stroke due to extracranial large artery atherosclerosis (disorder) |
|  | 724787004 | Epilepsy due to cerebrovascular accident (disorder) |
|  | 724993002 | Cerebral ischemic stroke due to occlusion of extracranial large artery (disorder) |
|  | 724994008 | Cerebral ischemic stroke due to stenosis of extracranial large artery (disorder) |
|  | 725132001 | Ischemic stroke without residual deficits (disorder) |
|  | 23671000119107 | Sequela of ischemic cerebral infarction (disorder) |
|  | 33301000119105 | Sequela of cardioembolic stroke |
|  | 33331000119103 | Sequela of lacunar stroke |
|  | 33331000119103 | Sequela of lacunar stroke |
|  | 46421000119102 | Behaviour disorder as sequela of cerebral infarction |
|  | 91601000119109 | Sequela of thrombotic stroke (disorder) |
|  | 118951000119103 | History of thrombotic stroke without residual deficits |
|  | 118961000119101 | History of hemorrhagic cerebrovascular accident without residual deficits (situation) |
|  | 125081000119106 | Cerebral infarction due to occlusion of precerebral artery |
|  | 140701000119108 | History of hemorrhagic stroke with hemiparesis (situation) |
|  | 140711000119106 | History of hemorrhagic stroke with hemiplegia |
|  | 140911000119109 | Ischaemic stroke with coma |
|  | 140921000119102 | Ischaemic stroke without coma |
|  | 141281000119101 | History of ischemic stroke without residual deficits (situation) |
|  | 141811000119106 | History of hemorrhagic cerebrovascular accident with residual deficit (situation) |
|  | 141821000119104 | History of ischemic cerebrovascular accident with residual deficit (situation) |
|  | 141831000119101 | History of embolic stroke with deficits |
|  | 145741000119101 | Apraxia as late effect of cerebrovascular disease |
|  | 293831000119105 | Cerebral infarction due to stenosis of precerebral artery |
|  | 672441000119103 | Hemiplegia of nondominant side due to and following ischemic cerebrovascular accident (disorder) |
|  | 672461000119104 | Hemiplegia of dominant side due to and following ischemic cerebrovascular accident (disorder) |
|  | 672501000119104 | Dysarthria due to and following ischemic cerebrovascular accident (disorder) |
|  | 672521000119108 | Dysphasia due to and following ischemic cerebrovascular accident (disorder) |
|  | 672541000119102 | Aphasia due to and following ischaemic cerebrovascular accident |
|  | 672561000119103 | Cognitive deficit due to and following ischemic cerebrovascular accident (disorder) |
|  | 674091000119108 | Vertigo due to and following ischemic cerebrovascular accident (disorder) |
|  | 674111000119100 | Ataxia due to and following ischemic cerebrovascular accident |
|  | 674121000119107 | Ataxia due to and following hemorrhagic cerebrovascular accident (disorder) |
|  | 674161000119102 | Monoplegia of upper limb due to and following ischemic cerebrovascular accident (disorder) |
|  | 674361000119104 | Apraxia due to and following ischemic cerebrovascular accident (disorder) |
|  | 674381000119108 | Weakness of facial muscle due to and following ischemic cerebrovascular accident (disorder) |
|  | 674391000119106 | Speech and language deficit due to and following hemorrhagic cerebrovascular accident (disorder) |
|  | 674401000119108 | Speech and language deficit due to and following ischemic cerebrovascular accident (disorder) |
|  | 16896891000119100 | History of cerebrovascular accident due to ischemia (situation) |
|  | 1089421000000100 | Cerebral infarction due to stenosis of cerebral artery (disorder) |
|  | 16000351000119100 | Cerebrovascular accident due to occlusion of left posterior cerebral artery (disorder) |
|  | 16000431000119100 | Cerebrovascular accident due to occlusion of right middle cerebral artery (disorder) |
|  | 16000511000119100 | Cerebrovascular accident due to occlusion of left middle cerebral artery (disorder) |
|  | 16002031000119100 | Cerebrovascular accident due to thrombus of right middle cerebral artery (disorder) |
|  | 16002111000119100 | Cerebrovascular accident due to thrombus of left middle cerebral artery (disorder) |
|  | 16218291000119100 | Acute cerebral ischemia (disorder) |
|  | 195230003 | Cerebral infarction due to cerebral venous thrombosis, non-pyogenic (disorder) |
|  | 20059004 | Occlusion of cerebral artery (disorder) |
|  | 230693009 | Anterior cerebral circulation infarction (disorder) |
|  | 230696001 | Posterior cerebral circulation infarction (disorder) |
|  | 230701008 | Pure sensorimotor lacunar infarction (disorder) |
|  | 292851000119109 | Lacunar ataxic hemiparesis of right dominant side (disorder) |
|  | 307363008 | Multiple lacunar infarcts (disorder) |
|  | 329361000119107 | Cerebrovascular accident due to occlusion of right middle cerebral artery by embolus (disorder) |
|  | 329421000119107 | Cerebrovascular accident due to occlusion of right posterior cerebral artery by embolus (disorder) |
|  | 329431000119105 | Cerebrovascular accident due to occlusion of left posterior cerebral artery by embolus (disorder) |
|  | 329461000119102 | Cerebrovascular accident due to occlusion of left cerebellar artery by embolus (disorder) |
|  | 329561000119101 | Occlusion of right posterior cerebral artery (disorder) |
|  | 329641000119104 | Cerebrovascular accident due to thrombus of basilar artery (disorder) |
|  | 329651000119102 | Cerebrovascular accident due to thrombus of right carotid artery (disorder) |
|  | 330411000119109 | Lacunar ataxic hemiparesis of left nondominant side (disorder) |
|  | 330421000119102 | Lacunar ataxic hemiparesis of right nondominant side (disorder) |
|  | 371040005 | Thrombotic stroke (disorder) |
|  | 371041009 | Embolic stroke (disorder) |
|  | 444657001 | Superior cerebellar artery syndrome (disorder) |
|  | 724429004 | Stroke co-occurrent with migraine (disorder) |
|  | 734383005 | Thrombosis of left middle cerebral artery (disorder) |
|  | 734961002 | Embolus of left posterior cerebral artery (disorder) |
|  | 734963004 | Embolus of right posterior cerebral artery (disorder) |
|  | 734964005 | Embolus of left middle cerebral artery (disorder) |
|  | 762629007 | Occlusion of right middle cerebral artery by embolus (disorder) |
|  | 762630002 | Occlusion of left middle cerebral artery by embolus (disorder) |
|  | 762651004 | Occlusion of right posterior cerebral artery by embolus (disorder) |
|  | 87555007 | Claude's syndrome (disorder) |
|  | 1089411000000100 | Cerebral infarction due to occlusion of cerebral artery (disorder) |
|  | 14309005 | Anterior choroidal artery syndrome (disorder) |
|  | 15978431000119100 | Thrombosis of right vertebral artery (disorder) |
|  | 16000391000119100 | Cerebrovascular accident due to occlusion of right posterior cerebral artery (disorder) |
|  | 195213000 | Cerebellar stroke syndrome (disorder) |
|  | 230694003 | Total anterior cerebral circulation infarction (disorder) |
|  | 230695002 | Partial anterior cerebral circulation infarction (disorder) |
|  | 230702001 | Lacunar ataxic hemiparesis (disorder) |
|  | 230703006 | Dysarthria-clumsy hand syndrome (disorder) |
|  | 230704000 | Multi-infarct state (disorder) |
|  | 24654003 | Weber-Gubler syndrome (disorder) |
|  | 25133001 | Completed stroke (disorder) |
|  | 276219001 | Occipital cerebral infarction (disorder) |
|  | 276220007 | Foville syndrome (disorder) |
|  | 276221006 | Millard-Gubler syndrome (disorder) |
|  | 276222004 | Top of basilar syndrome (disorder) |
|  | 292861000119106 | Lacunar ataxic hemiparesis of left dominant side (disorder) |
|  | 329371000119101 | Cerebrovascular accident due to occlusion of left middle cerebral artery by embolus (disorder) |
|  | 329481000119106 | Occlusion of right middle cerebral artery (disorder) |
|  | 329491000119109 | Occlusion of left middle cerebral artery (disorder) |
|  | 329571000119107 | Occlusion of left posterior cerebral artery (disorder) |
|  | 330791000119108 | Cerebrovascular accident due to thrombus of left carotid artery (disorder) |
|  | 413758000 | Cardioembolic stroke (disorder) |
|  | 426107000 | Acute lacunar infarction (disorder) |
|  | 426983002 | Infarction of medulla oblongata (disorder) |
|  | 734384004 | Thrombosis of right middle cerebral artery (disorder) |
|  | 734965006 | Embolus of right middle cerebral artery (disorder) |
|  | 762652006 | Occlusion of left posterior cerebral artery by embolus (disorder) |
|  | 90099008 | Subcortical leukoencephalopathy (disorder) |

**Supplementary Table 2b: Codelists for Haemorrhagic Stroke (see https://portal.caliberresearch.org/collections/bhf-data-science-centre)**

| **Code type** | **Code** | **Descriptions** |
| --- | --- | --- |
| **ICD10 codes** | I61 | Intracerebral haemorrhage |
|  | I69.1 | Sequelae of intracerebral haemorrhage |
| SNOMED CONCEPT IDS | 1508000 | Intracerebral hemorrhage |
|  | 7713009 | Intrapontine hemorrhage |
|  | 10458001 | Evacuation of intracerebral hematoma |
|  | 28837001 | Bulbar hemorrhage |
|  | 49422009 | Cortical hemorrhage |
|  | 52201006 | Internal capsule hemorrhage |
|  | 75038005 | Cerebellar hemorrhage |
|  | 195165005 | Basal ganglia hemorrhage |
|  | 195167002 | External capsule hemorrhage |
|  | 195168007 | Intracerebral haemorrhage, intraventricular |
|  | 195169004 | Intracerebral hemorrhage, multiple localized |
|  | 195241001 | Sequelae of intracerebral haemorrhage |
|  | 195242008 | Sequelae of other non-traumatic intracranial hemorrhage |
|  | 230710000 | Lobar cerebral hemorrhage |
|  | 274100004 | Cerebral hemorrhage |
|  | 308128006 | Right sided intracerebral hemorrhage, unspecified |
|  | 417506008 | Hemorrhagic stroke monitoring (regime/therapy) |
|  | 428267002 | History of cerebral hemorrhage (situation) |
|  | 732923001 | Hemorrhage of medulla oblongata (disorder) |
|  | 118961000119101 | History of hemorrhagic cerebrovascular accident without residual deficits (situation) |
|  | 140701000119108 | History of hemorrhagic stroke with hemiparesis (situation) |
|  | 140711000119106 | History of hemorrhagic stroke with hemiplegia |
|  | 141811000119106 | History of hemorrhagic cerebrovascular accident with residual deficit (situation) |
|  | 145741000119101 | Apraxia as late effect of cerebrovascular disease |
|  | 674121000119107 | Ataxia due to and following hemorrhagic cerebrovascular accident (disorder) |
|  | 674391000119106 | Speech and language deficit due to and following hemorrhagic cerebrovascular accident (disorder) |
|  | 230712008 | Lacunar hemorrhage (disorder) |
|  | 276722003 | Intracerebellar and posterior fossa hemorrhage (disorder) |
|  | 291531000119108 | Spontaneous hemorrhage of cerebral hemisphere (disorder) |
|  | 291541000119104 | Spontaneous hemorrhage of brain stem (disorder) |
|  | 95454007 | Brain stem hemorrhage (disorder) |
|  | 1078001000000100 | Haemorrhagic stroke (disorder) |
|  | 20908003 | Subcortical cerebral hemorrhage (disorder) |
|  | 230709005 | Massive supratentorial cerebral hemorrhage (disorder) |
|  | 230711001 | Thalamic hemorrhage (disorder) |
|  | 291511000119103 | Spontaneous hemorrhage of deep cerebral hemisphere (disorder) |
|  | 291521000119105 | Spontaneous hemorrhage of cortical intracerebral hemisphere (disorder) |

**Supplementary Table 2c: Codelists for Unspecified Stroke (see https://portal.caliberresearch.org/collections/bhf-data-science-centre)**

| **Code type** | **Code** | **Descriptions** |
| --- | --- | --- |
| **ICD10 codes** | G46.3 | Brain stem stroke syndrome |
|  | G46.4 | Cerebellar stroke syndrome |
|  | G46.5 | Pure motor lacunar syndrome |
|  | G46.6 | Pure sensory lacunar syndrome |
|  | G46.7 | Other lacunar syndromes |
|  | G46.8 | Other vascular syndromes of brain in cerebrovascular diseases |
|  | I64 | Stroke, not specified as haemorrhage or infarction |
|  | I69.4 | Sequelae of stroke, not specified as haemorrhage or infarction |
| SNOMED CONCEPT IDS | 6594005 | Cerebrovascular disorder in the puerperium |
|  | 170600009 | Stroke monitoring |
|  | 195212005 | Brainstem stroke syndrome |
|  | 195213000 | Cerebellar stroke syndrome |
|  | 195216008 | Left sided cerebral hemisphere cerebrovascular accident |
|  | 195217004 | Right sided CVA |
|  | 195239002 | Late effects of cerebrovascular disease |
|  | 195608005 | [X]Sequelae of stroke, not specified as haemorrhage or infarction |
|  | 230690007 | CVA - Cerebrovascular accident |
|  | 275434003 | Stroke in the puerperium |
|  | 275526006 | H/O: CVA |
|  | 275527002 | H/O: stroke |
|  | 277286006 | CPSP - Central post-stroke pain |
|  | 308067002 | H/O: Stroke in last year |
|  | 425642008 | Monoplegia of dominant lower limb as a late effect of cerebrovascular accident (disorder) |
|  | 425882004 | Paralytic syndrome as late effect of stroke (disorder) |
|  | 426033005 | Dysphagia as a late effect of cerebrovascular accident (disorder) |
|  | 426788002 | Vertigo as late effect of stroke (disorder) |
|  | 427065003 | Monoplegia of dominant upper limb as a late effect of cerebrovascular accident (disorder) |
|  | 427432001 | Paralytic syndrome as late effect of thalamic stroke (disorder) |
|  | 428668000 | Apraxia due to cerebrovascular accident (disorder) |
|  | 429993008 | History of cerebrovascular accident without residual deficits (situation) |
|  | 430947007 | Paralytic syndrome of nondominant side as late effect of stroke (disorder) |
|  | 430959006 | Paralytic syndrome of dominant side as late effect of stroke (disorder) |
|  | 431310008 | History of occlusion of cerebral artery (situation) |
|  | 433183000 | Neurogenic bladder as late effect of cerebrovascular accident (disorder) |
|  | 440140008 | History of cerebrovascular accident with residual deficit (situation) |
|  | 441529001 | Dysphasia as late effect of cerebrovascular disease |
|  | 441630004 | Aphasia as late effect of cerebrovascular disease |
|  | 441735003 | Sensory disorder as a late effect of cerebrovascular disease |
|  | 441759008 | Abnormal vision as a late effect of cerebrovascular disease |
|  | 441887006 | Monoplegia of lower limb as late effect of cerebrovascular disease |
|  | 441894009 | Monoplegia of nondominant lower limb as a late effect of cerebrovascular accident |
|  | 441960006 | Speech and language deficit as late effect of cerebrovascular accident |
|  | 441991000 | Hemiparesis as late effect of cerebrovascular accident |
|  | 442024001 | Hemiplegia as late effect of cerebrovascular disease |
|  | 442097001 | Monoplegia of upper limb as late effect of cerebrovascular disease |
|  | 441894009 | Monoplegia of nondominant lower limb as a late effect of cerebrovascular accident |
|  | 441960006 | Speech and language deficit as late effect of cerebrovascular accident |
|  | 441991000 | Hemiparesis as late effect of cerebrovascular accident |
|  | 442024001 | Hemiplegia as late effect of cerebrovascular disease |
|  | 442097001 | Monoplegia of upper limb as late effect of cerebrovascular disease |
|  | 442181008 | Monoplegia of nondominant upper limb as a late effect of cerebrovascular accident |
|  | 442212003 | Residual cognitive deficit as late effect of cerebrovascular accident |
|  | 442617003 | Aphasia as late effect of cerebrovascular accident |
|  | 442668000 | Hemiplegia of nondominant side as late effect of cerebrovascular disease |
|  | 442676003 | Hemiplegia of dominant side as late effect of cerebrovascular disease |
|  | 442733008 | Hemiplegia as late effect of cerebrovascular accident |
|  | 698767004 | Post-cerebrovascular accident epilepsy |
|  | 699270006 | Stroke annual review |
|  | 699429007 | History of cerebrovascular accident in last eight weeks (situation) |
|  | 713410003 | Pain following cerebrovascular accident |
|  | 720849008 | Education about stroke |
|  | 722929005 | Perinatal arterial ischemic stroke (disorder) |
|  | 40161000119102 | Weakness of face muscles as sequela of stroke |
|  | 48601000119107 | Hemiplegia and/or hemiparesis following stroke |
|  | 87551000119101 | Visual disturbance as sequela of cerebrovascular disease (disorder) |
|  | 92341000119107 | Weakness of extremities as sequela of stroke |
|  | 97531000119106 | History of parietal cerebrovascular accident |
|  | 99051000119101 | History of lacunar cerebrovascular accident |
|  | 102831000119104 | Paraplegia or paraparesis as sequela of stroke |
|  | 103761000119107 | Paralytic syndrome of all four limbs as sequela of stroke (disorder) |
|  | 133981000119106 | Dysarthria as late effects of cerebrovascular disease |
|  | 133991000119109 | Fluency disorder as sequela of cerebrovascular disease |
|  | 134771000119108 | Alteration of sensation as late effect of stroke (disorder) |
|  | 137991000119103 | Seizure disorder as sequela of stroke |
|  | 140281000119108 | Hemiparesis as late effect of cerebrovascular disease |
|  | 148871000119109 | Weakness as a late effect of stroke |
|  | 186831000119104 | Apraxia due to and following cerebrovascular accident (disorder) |
|  | 290581000119101 | Ataxia due to and following cerebrovascular accident (disorder) |
|  | 290621000119101 | Cognitive deficit due to and following cerebrovascular disease (disorder) |
|  | 290631000119103 | Dysarthria due to and following cerebrovascular accident (disorder) |
|  | 290791000119105 | Fluency disorder due to and following cerebrovascular accident (disorder) |
|  | 290931000119108 | Monoplegia of lower limb due to and following cerebrovascular accident (disorder) |
|  | 291091000119102 | Monoplegia of left nondominant upper limb due to and following cerebrovascular accident (disorder) |
|  | 291111000119105 | Monoplegia of right nondominant upper limb due to and following cerebrovascular accident (disorder) |
|  | 291121000119103 | Monoplegia of upper limb due to and following cerebrovascular accident (disorder) |
|  | 292851000119109 | Lacunar ataxic hemiparesis of right dominant side (disorder) |
|  | 292861000119106 | Lacunar ataxic hemiparesis of left dominant side (disorder) |
|  | 330411000119109 | Lacunar ataxic hemiparesis of left nondominant side (disorder) |
|  | 330421000119102 | Lacunar ataxic hemiparesis of right nondominant side (disorder) |
|  | 690051000119100 | History of stroke of cerebellum |
|  | 12242711000119100 | Weakness of left facial muscle due to and following cerebrovascular accident (disorder) |
|  | 12242751000119100 | Weakness of right facial muscle due to and following cerebrovascular accident (disorder) |
|  | 12367511000119100 | Paraplegia due to and following cerebrovascular accident (disorder) |
|  | 15982271000119100 | Weakness of right facial muscle due to and following cerebrovascular disease (disorder) |
|  | 15982311000119100 | Weakness of left facial muscle due to and following cerebrovascular disease (disorder) |
|  | 16260551000119100 | Dysphasia due to and following cerebrovascular accident (disorder) |
|  | 111297002 | Nonparalytic stroke (disorder) |
|  | 116288000 | Paralytic stroke (disorder) |
|  | 16371781000119100 | Cerebellar stroke (disorder) |
|  | 195209007 | Middle cerebral artery syndrome (disorder) |
|  | 195210002 | Anterior cerebral artery syndrome (disorder) |
|  | 195211003 | Posterior cerebral artery syndrome (disorder) |
|  | 195212005 | Brainstem stroke syndrome (disorder) |
|  | 195216008 | Left sided cerebral hemisphere cerebrovascular accident (disorder) |
|  | 195217004 | Right sided cerebral hemisphere cerebrovascular accident (disorder) |
|  | 230706003 | Hemorrhagic cerebral infarction (disorder) |
|  | 230708002 | Posterior cerebral circulation hemorrhagic infarction (disorder) |
|  | 281240008 | Extension of cerebrovascular accident (disorder) |
|  | 297138001 | Embolus of circle of Willis (disorder) |
|  | 444172003 | Recurrent transient cerebral ischemic attack (disorder) |
|  | 230690007 | Cerebrovascular accident (disorder) |
|  | 230707007 | Anterior cerebral circulation hemorrhagic infarction (disorder) |
|  | 230713003 | Stroke of uncertain pathology (disorder) |
|  | 230714009 | Anterior circulation stroke of uncertain pathology (disorder) |
|  | 230715005 | Posterior circulation stroke of uncertain pathology (disorder) |
|  | 275434003 | Stroke in the puerperium (disorder) |
|  | 426814001 | Transient cerebral ischemia due to atrial fibrillation (disorder) |
|  | 57981008 | Progressing stroke (disorder) |

**Supplementary Table 2d: Codelists for Transient Ischaemic Attack (see https://portal.caliberresearch.org/collections/bhf-data-science-centre)**

| **Code type** | **Code** | **Descriptions** |
| --- | --- | --- |
| **ICD10 codes** | G45.0 | Vertebro-basilar artery syndrome |
|  | G45.1 | Carotid artery syndrome (hemispheric) |
|  | G45.2 | Multiple and bilateral precerebral artery syndromes |
|  | G45.3 | Amaurosis fugax |
|  | G45.4 | Transient global amnesia |
|  | G45.8 | Other transient cerebral ischaemic attacks and related syndromes |
|  | G45.9 | Transient cerebral ischaemic attack, unspecified |
|  | G46.0 | Middle cerebral artery syndrome |
|  | G46.1 | Anterior cerebral artery syndrome |
|  | G46.2 | Posterior cerebral artery syndrome |
|  | I65 | Occlusion and stenosis of precerebral arteries, not resulting in cerebral infarction |
|  | I66 | Occlusion and stenosis of cerebral arteries, not resulting in cerebral infarction |
| SNOMED CONCEPT IDS | 88032003 | Amaurosis fugax |
|  | 195206000 | Intermittent cerebral ischaemia |
|  | 230716006 | Anterior circulation transient ischaemic attack |
|  | 266257000 | TIA |
|  | 710575003 | Transient ischemic attack due to embolism |
|  | 140221000119109 | History of transient ischemic attack due to embolism (situation) |
|  | 13016361000119100 | History of amaurosis fugax (situation) |
|  | 15258001 | Subclavian steal syndrome (disorder) |
|  | 195199008 | Vertebrobasilar artery syndrome (disorder) |
|  | 195205001 | Impending cerebral ischemia (disorder) |
|  | 34781003 | Vertebral artery syndrome (disorder) |
|  | 64009001 | Basilar artery syndrome (disorder) |
|  | 751371000000107 | Personal history of transient ischaemic attack (situation) |
|  | 230717002 | Vertebrobasilar territory transient ischemic attack (disorder) |

**Supplementary Table 3: Codelists for Obesity (see https://portal.caliberresearch.org/collections/bhf-data-science-centre)**

| **Code type** | **Code** | **Descriptions** |
| --- | --- | --- |
| **ICD10 codes** | E66 | Obesity |
| SNOMED CONCEPT IDS | 162864005 | Body mass index 30+ - obesity (finding) |
|  | 408512008 | Body mass index 40+ - severely obese (finding) |
|  | 914721000000105 | Obese class I (body mass index 30.0 - 34.9) (finding) |
|  | 914731000000107 | Obese class II (body mass index 35.0 - 39.9) (finding) |
|  | 914741000000103 | Obese class III (body mass index equal to or greater than 40.0) (finding) |
|  | 921031000000102 | Child body mass index 98.1st-99.6th centile (finding) |
|  | 443371000124107 | Obese class I (finding) |
|  | 443381000124105 | Obese class II (finding) |
|  | 921051000000109 | Child body mass index greater than 99.6th centile (finding) |

**Supplementary Table 4: Codelists for Diabetes (see https://portal.caliberresearch.org/collections/bhf-data-science-centre)**

| **ICD10 codes** | E10 | Insulin-dependent diabetes mellitus |
| --- | --- | --- |
|  | E11 | Non-insulin-dependent diabetes mellitus |
|  | E12 | Malnutrition-related diabetes mellitus |
|  | O242 | Diabetes mellitus in pregnancy: Pre-existing malnutrition-related diabetes mellitus |
|  | E13 | Other specified diabetes mellitus |
|  | E14 | Unspecified diabetes mellitus |
|  | G590 | Diabetic mononeuropathy |
|  | G632 | Diabetic polyneuropathy |
|  | H280 | Diabetic cataract |
|  | H360 | Diabetic retinopathy |
|  | M142 | Diabetic arthropathy |
|  | N083 | Glomerular disorders in diabetes mellitus |
|  | O240 | Diabetes mellitus in pregnancy: Pre-existing diabetes mellitus, insulin-dependent |
|  | O241 | Diabetes mellitus in pregnancy: Pre-existing diabetes mellitus, non-insulin-dependent |
|  | O243 | Diabetes mellitus in pregnancy: Pre-existing diabetes mellitus, unspecified |
| SNOMED CONCEPT IDS | 4855003 | Diabetic retinopathy |
|  | 6143009 | Diabetic patient education |
|  | 8801005 | Secondary diabetes mellitus |
|  | 9859006 | Insulin-resistant diabetes mellitus AND acanthosis nigricans |
|  | 11530004 | Brittle diabetes |
|  | 19378003 | Diabetic pseudotabes |
|  | 19429009 | Chronic ulcer of skin |
|  | 21858001 | Diabetes with renal manifestations |
|  | 24927004 | Diabetes with ketoacidosis |
|  | 25093002 | Diabetic oculopathy |
|  | 25412000 | Diabetic retinal microaneurysm |
|  | 25907005 | Diabetic gangrene |
|  | 26298008 | Diabetic coma with ketoacidosis |
|  | 28453007 | Maturity onset diabetes mellitus in young |
|  | 31411005 | Background diabetic retinopathy |
|  | 33248009 | Diabetes with non-ketotic non-hyperosmolar coma |
|  | 34140002 | Diabetic gastroparesis |
|  | 35777006 | Diabetic mononeuropathy multiplex |
|  | 38046004 | Diffuse type diabetic glomerulosclerosis |
|  | 38205001 | Diarrhea in diabetes |
|  | 39058009 | Diabetic amyotrophy |
|  | 39127005 | Symmetric diabetic proximal motor neuropathy |
|  | 39181008 | Diabetic radiculopathy |
|  | 43959009 | Diabetic cataract |
|  | 44054006 | Diabetes mellitus type II |
|  | 46635009 | Insulin dependent diabetes mellitus |
|  | 48951005 | Bullosis diabeticorum |
|  | 49455004 | Diabetic polyneuropathy |
|  | 50620007 | Diabetic autonomic neuropathy |
|  | 51002006 | Diabetes mellitus associated with pancreatic disease |
|  | 54181000 | Diabetes-nephrosis syndrome |
|  | 55692006 | Diabetes with hyperosmolar coma |
|  | 59276001 | Proliferative diabetic retinopathy |
|  | 62260007 | Pretibial pigmental patches in diabetes |
|  | 63510008 | Nodular type diabetic glomerulosclerosis |
|  | 73211009 | Diabetes mellitus |
|  | 74627003 | Diabetic complication |
|  | 75524006 | Malnutrition related diabetes mellitus |
|  | 79554005 | Asymmetric diabetic proximal motor neuropathy |
|  | 81830002 | Diabetic mononeuropathy simplex |
|  | 82980005 | Anemia of diabetes |
|  | 110996009 | Armanni-Ebstein kidney |
|  | 111552007 | Diabetes mellitus without complication |
|  | 111556005 | Diabetic ketoacidosis without coma |
|  | 111557001 | Diabetes with coma |
|  | 111558006 | Insulin coma |
|  | 126534007 | Diabetic mixed sensory-motor polyneuropathy |
|  | 126535008 | Diabetic motor polyneuropathy |
|  | 127011001 | Diabetic sensory polyneuropathy |
|  | 127012008 | Lipoatrophic diabetes |
|  | 127013003 | Diabetic renal disease |
|  | 127014009 | Diabetic peripheral angiopathy |
|  | 134395001 | Diabetic retinopathy screening |
|  | 161649006 | H/O: insulin therapy |
|  | 170745003 | Diabetic on diet only |
|  | 170746002 | Diabetic on oral treatment |
|  | 170747006 | Diabetic on insulin |
|  | 170763003 | Diabetic - good control |
|  | 170766006 | Loss of hypoglycemic warning |
|  | 170769004 | Diabetic - cooperative patient |
|  | 170770003 | Diabetic-uncooperative patient |
|  | 171183004 | Diabetes mellitus screening |
|  | 183056000 | Patient advised about diabetic diet |
|  | 190325001 | Diabetes mellitus, juvenile type, with ketoacidosis |
|  | 190326000 | Diabetes mellitus, adult onset, with ketoacidosis |
|  | 190327009 | Other specified diabetes mellitus with ketoacidosis |
|  | 190328004 | Diabetes mellitus NOS with ketoacidosis |
|  | 190329007 | Diabetes mellitus with hyperosmolar coma |
|  | 190330002 | Diabetes mellitus, juvenile type, with hyperosmolar coma |
|  | 190331003 | Diabetes mellitus, adult onset, with hyperosmolar coma |
|  | 190332005 | Diabetes mellitus NOS with hyperosmolar coma |
|  | 190333000 | Diabetes mellitus with ketoacidotic coma |
|  | 190334006 | Diabetes mellitus, juvenile type, with ketoacidotic coma |
|  | 190335007 | Diabetes mellitus, adult onset, with ketoacidotic coma |
|  | 190337004 | Diabetes mellitus NOS with ketoacidotic coma |
|  | 190339001 | Diabetes mellitus, juvenile type, with renal manifestation |
|  | 190340004 | Diabetes mellitus, adult onset, with renal manifestation |
|  | 190341000 | Other specified diabetes mellitus with renal complications |
|  | 190342007 | Diabetes mellitis with nephropathy NOS |
|  | 190343002 | Diabetes mellitus with ophthalmic manifestation |
|  | 190345009 | Diabetes mellitus, juvenile type, with ophthalmic manifestation |
|  | 190346005 | Diabetes mellitus, adult onset, with ophthalmic manifestation |
|  | 190347001 | Other specified diabetes mellitus with ophthalmic complications |
|  | 190348006 | Diabetes mellitus NOS with ophthalmic manifestation |
|  | 190350003 | Diabetes mellitus, juvenile type, with neurological manifestation |
|  | 190351004 | Diabetes mellitus, adult onset, with neurological manifestation |
|  | 190352006 | Other specified diabetes mellitus with neurological complications |
|  | 190353001 | Diabetes mellitus NOS with neurological manifestation |
|  | 190354007 | Diabetes mellitus with: [gangrene] or [peripheral circulatory disorder] |
|  | 190355008 | Diabetes mellitus, juvenile type, with peripheral circulatory disorder |
|  | 190356009 | Diabetes mellitus, adult onset, with peripheral circulatory disorder |
|  | 190357000 | Diabetes mellitus, adult with gangrene |
|  | 190358005 | IDDM with peripheral circulatory disorder |
|  | 190359002 | NIDDM with peripheral circulatory disorder |
|  | 190360007 | Other specified diabetes mellitus with peripheral circulatory complications |
|  | 190361006 | Diabetes mellitus NOS with peripheral circulatory disorder |
|  | 190363009 | Type I diabetes mellitus with renal complications |
|  | 190364003 | Type 1 diabetes mellitus with ophthalmic complications |
|  | 190365002 | Type 1 diabetes mellitus with neurological complications |
|  | 190366001 | Type I diabetes mellitus with multiple complications |
|  | 190368000 | Type I diabetes mellitus with ulcer |
|  | 190369008 | Type 1 diabetes mellitus with gangrene |
|  | 190370009 | Type I diabetes mellitus with retinopathy |
|  | 190372001 | Type I diabetes mellitus maturity onset |
|  | 190385003 | Type 2 diabetes mellitus with renal complications |
|  | 190386002 | Type II diabetes mellitus with ophthalmic complications |
|  | 190387006 | Type 2 diabetes mellitus with neurological complications |
|  | 190388001 | Type II diabetes mellitus with multiple complications |
|  | 190389009 | Type 2 diabetes mellitus with ulcer |
|  | 190390000 | Type II diabetes mellitus with gangrene |
|  | 190391001 | Type 2 diabetes mellitus with retinopathy |
|  | 190405001 | Malnutrition-related diabetes mellitus with coma |
|  | 190406000 | Malnutrition-related diabetes mellitus with ketoacidosis |
|  | 190408004 | Malnutrition-related diabetes mellitus with ophthalmic complications |
|  | 190409007 | Malnutrition-related diabetes mellitus with neurological complications |
|  | 190410002 | Malnutrition-related diabetes mellitus with peripheral circulatory complications |
|  | 190411003 | Malnutrition-related diabetes mellitus with multiple complications |
|  | 190422004 | Diabetes mellitus with unspecified complication |
|  | 190423009 | Diabetes mellitus, juvenile type, with unspecified complication |
|  | 190424003 | Diabetes mellitus, adult onset, with unspecified complication |
|  | 190425002 | Other specified diabetes mellitus with unspecified complications |
|  | 190426001 | Diabetes mellitus NOS with unspecified complication |
|  | 190430003 | Hypoglycemic coma NOS |
|  | 193141005 | Diabetic mononeuritis multiplex |
|  | 193156001 | Diabetic mononeuritis NOS |
|  | 193183000 | Acute painful diabetic neuropathy |
|  | 193184006 | Chronic painful diabetic neuropathy |
|  | 193185007 | Asymptomatic diabetic neuropathy |
|  | 193349004 | Preproliferative diabetic retinopathy |
|  | 193350004 | Advanced diabetic maculopathy |
|  | 193353002 | Diabetic retinopathy NOS |
|  | 193489006 | Diabetic iritis |
|  | 197605007 | Nephrotic syndrome in diabetes mellitus |
|  | 199229001 | Pre-existing diabetes mellitus, insulin-dependent |
|  | 199230006 | Pre-existing diabetes mellitus, non-insulin-dependent |
|  | 199231005 | Pre-existing malnutrition-related diabetes mellitus |
|  | 200687002 | Cellulitis in diabetic foot |
|  | 201250006 | Ischaemic ulcer diabetic foot |
|  | 201251005 | Neuropathic diabetic ulcer - foot |
|  | 201252003 | Mixed diabetic ulcer - foot |
|  | 201723002 | Diabetic hand syndrome |
|  | 201724008 | Diabetic Charcot's arthropathy |
|  | 230572002 | Diabetic neuropathy |
|  | 230573007 | Diabetic distal sensorimotor polyneuropathy |
|  | 230574001 | Diabetic acute painful polyneuropathy |
|  | 230575000 | Diabetic chronic painful polyneuropathy |
|  | 230576004 | Diabetic asymmetric polyneuropathy |
|  | 230577008 | Diabetic mononeuropathy |
|  | 230578003 | Diabetic truncal radiculopathy |
|  | 230579006 | Diabetic thoracic radiculopathy |
|  | 232019003 | Visually threatening diabetic retinopathy |
|  | 232020009 | Diabetic maculopathy |
|  | 232021008 | Proliferative diabetic retinopathy new vessels on disc |
|  | 232022001 | Proliferative diabetic retinopathy with new vessels elsewhere than on disc |
|  | 232023006 | Diabetic traction retinal detachment |
|  | 236499007 | Microalbuminuric diabetic nephropathy |
|  | 236500003 | Clinical diabetic nephropathy |
|  | 237599002 | Insulin-treated non-insulin-dependent diabetes mellitus |
|  | 237604008 | Diabetes mellitus autosomal dominant type II |
|  | 237620003 | Abnormal metabolic state in diabetes mellitus |
|  | 237621004 | Diabetic severe hyperglycemia |
|  | 237633009 | Hypoglycemic state in diabetes |
|  | 237635002 | Nocturnal hypoglycemia |
|  | 238981002 | Soft tissue complication of diabetes mellitus |
|  | 238982009 | Diabetic dermopathy |
|  | 238983004 | Diabetic thick skin syndrome |
|  | 238984005 | Diabetic rubeosis |
|  | 248542005 | Diabetic relative |
|  | 267381003 | Diabetes mellitus with renal manifestation |
|  | 267382005 | Diabetes mellitus with neurological manifestation |
|  | 267383000 | Diabetes mellitus with peripheral circulatory disorder |
|  | 267384006 | Hypoglycaemic coma |
|  | 267474009 | Diabetes with other complications |
|  | 267604001 | Myasthenic syndrome due to diabetic amyotrophy |
|  | 268519009 | Diabetic - poor control |
|  | 274589008 | [EDTA] Diabetes Type I (insulin dependent) associated with renal failure |
|  | 274590004 | [EDTA] Diabetes Type II (non-insulin-dependent) associated with renal failure |
|  | 275522008 | Diabetes mellitus with gangrene |
|  | 280137006 | Diabetic foot |
|  | 281023007 | Diabetic child |
|  | 284350006 | Dietary advice for diabetes mellitus |
|  | 286912007 | Diabetes with ketoacidosis - no coma |
|  | 290002008 | Unstable type I diabetes mellitus |
|  | 308105005 | O/E - Right diabetic foot at risk |
|  | 308106006 | O/E - Left diabetic foot at risk |
|  | 309426007 | Diabetic glomerulopathy |
|  | 309595004 | Retinal abnormality - diabetes-related |
|  | 309635005 | H/O: Admission in last year for diabetes foot problem |
|  | 310387003 | Diabetic intracapillary glomerulosclerosis |
|  | 310505005 | Diabetic hyperosmolar non-ketotic state |
|  | 311366001 | Kimmelstiel-Wilson syndrome |
|  | 311782002 | Advanced diabetic retinal disease |
|  | 312903003 | Mild non proliferative diabetic retinopathy |
|  | 312904009 | Moderate non proliferative diabetic retinopathy |
|  | 312905005 | Severe non proliferative diabetic retinopathy |
|  | 312906006 | Proliferative diabetic retinopathy - non high risk |
|  | 312907002 | Proliferative diabetic retinopathy - high risk |
|  | 312908007 | Proliferative diabetic retinopathy - quiescent |
|  | 312909004 | Proliferative diabetic retinopathy - iris neovascularisation |
|  | 312910009 | Diabetic vitreous hemorrhage |
|  | 312912001 | Diabetic macular edema |
|  | 313435000 | Type I diabetes mellitus without complication |
|  | 313436004 | Non-insulin-dependent diabetes mellitus without complication |
|  | 314010006 | Diffuse diabetic maculopathy |
|  | 314011005 | Focal diabetic maculopathy |
|  | 314014002 | Ischaemic diabetic maculopathy |
|  | 314015001 | Mixed diabetic maculopathy |
|  | 314194001 | Diabetic on insulin and oral treatment |
|  | 314368001 | Type I diabetes mellitus with mononeuropathy |
|  | 314369009 | Type 1 diabetes mellitus with polyneuropathy |
|  | 314370005 | Type II diabetes mellitus with mononeuropathy |
|  | 314371009 | Type II diabetes mellitus with polyneuropathy |
|  | 314377008 | Type I diabetes mellitus with nephropathy |
|  | 314378003 | Type 2 diabetes mellitus with nephropathy |
|  | 314537004 | Diabetic optic papillopathy |
|  | 314771006 | Type 1 diabetes mellitus with hypoglycaemic coma |
|  | 314772004 | Type 2 diabetes mellitus with hypoglycaemic coma |
|  | 314887002 | Insulin dependent diabetes mellitus with diabetic cataract |
|  | 314888007 | Type II diabetes mellitus with diabetic cataract |
|  | 314892000 | Type I diabetes mellitus with peripheral angiopathy |
|  | 314893005 | Type 1 diabetes mellitus with arthropathy |
|  | 314894004 | Type I diabetes mellitus with neuropathic arthropathy |
|  | 314902007 | Type II diabetes mellitus with peripheral angiopathy |
|  | 314903002 | Non-insulin dependent diabetes mellitus with arthropathy |
|  | 314904008 | Type II diabetes mellitus with neuropathic arthropathy |
|  | 359611005 | Diabetic neuropathy with neurologic complication |
|  | 361216007 | Diabetic femoral mononeuropathy |
|  | 371054002 | Type I diabetes mellitus with complication (disorder) |
|  | 371055001 | Type I diabetes mellitus with ketoacidosis (disorder) |
|  | 371056000 | Type II diabetes mellitus with complication (disorder) |
|  | 371086007 | Diabetes mellitus with skin ulcer (disorder) |
|  | 371087003 | Diabetic foot ulcer (disorder) |
|  | 372069003 | Diabetes mellitus with complication (disorder) |
|  | 390834004 | Non proliferative diabetic retinopathy (disorder) |
|  | 390850007 | O/E - no right diabetic retinopathy (context-dependent category) |
|  | 390853009 | O/E - no left diabetic retinopathy (context-dependent category) |
|  | 390854003 | O/E - diabetic maculopathy present both eyes (context-dependent category) |
|  | 390855002 | O/E - diabetic maculopathy absent both eyes (context-dependent category) |
|  | 391178000 | High risk non proliferative diabetic retinopathy (disorder) |
|  | 395204000 | Hyperosmolar non-ketotic state in type 2 diabetes mellitus (disorder) |
|  | 398140007 | Somogyi phenomenon (disorder) |
|  | 398819009 | Diabetic foot at risk (context-dependent category) |
|  | 399862001 | Proliferative diabetic retinopathy - high risk with no macular edema (disorder) |
|  | 399863006 | Very severe nonproliferative diabetic retinopathy with no macular edema (disorder) |
|  | 399864000 | Diabetic macular edema not clinically significant (disorder) |
|  | 399865004 | Very severe proliferative diabetic retinopathy (disorder) |
|  | 399866003 | Diabetic retinal venous beading (disorder) |
|  | 399868002 | Diabetic intraretinal microvascular anomalies (disorder) |
|  | 399869005 | High risk proliferative diabetic retinopathy not amenable to photocoagulation (disorder) |
|  | 399870006 | Non-high-risk proliferative diabetic retinopathy with no macular edema (disorder) |
|  | 399871005 | Visually threatening diabetic retinopathy (disorder) |
|  | 399872003 | Severe nonproliferative diabetic retinopathy with clinically significant macular edema (disorder) |
|  | 399873008 | Severe nonproliferative diabetic retinopathy with no macular edema (disorder) |
|  | 399874002 | Proliferative diabetic retinopathy - high risk with clinically significant macular edema (disorder) |
|  | 399875001 | Non-high-risk proliferative diabetic retinopathy with clinically significant macular edema (disorder) |
|  | 399876000 | Very severe nonproliferative diabetic retinopathy (disorder) |
|  | 399877009 | Very severe nonproliferative diabetic retinopathy with clinically significant macular edema (disorder) |
|  | 401087005 | Diabetes mellitus with persistent microalbuminuria (disorder) |
|  | 401088000 | Diabetes mellitus with persistent proteinuria (disorder) |
|  | 401109007 | Type 1 diabetes mellitus with persistent proteinuria (disorder) |
|  | 401110002 | Type 1 diabetes mellitus with persistent microalbuminuria (disorder) |
|  | 401111003 | Type 2 diabetes mellitus with persistent proteinuria (disorder) |
|  | 401112005 | Type 2 diabetes mellitus with persistent microalbuminuria (disorder) |
|  | 401191002 | Diabetic foot examination (regime/therapy) |
|  | 403423006 | Congenital total lipoatrophy (disorder) |
|  | 408287009 | Type 1 diabetes mellitus with exudative maculopathy (disorder) |
|  | 408396006 | Diabetic retinopathy screening not indicated (context-dependent category) |
|  | 408397002 | Diabetic foot examination not indicated (context-dependent category) |
|  | 408409007 | O/E - right eye background diabetic retinopathy (context-dependent category) |
|  | 408410002 | O/E - left eye background diabetic retinopathy (context-dependent category) |
|  | 408411003 | O/E - right eye preproliferative diabetic retinopathy (context-dependent category) |
|  | 408412005 | O/E - left eye preproliferative diabetic retinopathy (context-dependent category) |
|  | 408413000 | O/E - right eye proliferative diabetic retinopathy (context-dependent category) |
|  | 408414006 | O/E - left eye proliferative diabetic retinopathy (context-dependent category) |
|  | 408415007 | O/E - right eye diabetic maculopathy (context-dependent category) |
|  | 408416008 | O/E - left eye diabetic maculopathy (context-dependent category) |
|  | 408417004 | Type 2 diabetes mellitus with exudative maculopathy (disorder) |
|  | 408660003 | Type II diabetes mellitus with ketoacidosis (disorder) |
|  | 412752009 | Diabetic foot examination declined (context-dependent category) |
|  | 413122001 | Diabetic retinopathy screening refused (context-dependent category) |
|  | 413180006 | Pan retinal photocoagulation for diabetes (procedure) |
|  | 414894003 | O/E - left eye stable treated proliferative diabetic retinopathy (context-dependent category) |
|  | 414910007 | O/E - right eye stable treated proliferative diabetic retinopathy (context-dependent category) |
|  | 417677008 | O/E - sight threatening diabetic retinopathy (context-dependent category) |
|  | 419100001 | Infection of foot associated with diabetes (disorder) |
|  | 420270002 | Ketoacidosis in type I diabetes mellitus (disorder) |
|  | 420279001 | Renal disorder associated with type II diabetes mellitus (disorder) |
|  | 420414003 | Multiple complications of type II diabetes mellitus (disorder) |
|  | 420422005 | Ketoacidosis in diabetes mellitus (disorder) |
|  | 420436000 | Mononeuropathy associated with type II diabetes mellitus (disorder) |
|  | 420486006 | Exudative maculopathy associated with type I diabetes mellitus (disorder) |
|  | 420514000 | Persistent proteinuria associated with type I diabetes mellitus (disorder) |
|  | 420662003 | Coma associated with diabetes mellitus (disorder) |
|  | 420683009 | Neurological disorder associated with malnutrition-related diabetes mellitus (disorder) |
|  | 420715001 | Persistent microalbuminuria associated with type II diabetes mellitus (disorder) |
|  | 420756003 | Diabetic cataract associated with type II diabetes mellitus (disorder) |
|  | 420789003 | Diabetic retinopathy associated with type I diabetes mellitus (disorder) |
|  | 420825003 | Gangrene associated with type I diabetes mellitus (disorder) |
|  | 420868002 | Disorder associated with type I diabetes mellitus (disorder) |
|  | 420918009 | Mononeuropathy associated with type I diabetes mellitus (disorder) |
|  | 420996007 | Coma associated with malnutrition-related diabetes mellitus (disorder) |
|  | 421075007 | Ketoacidotic coma in type I diabetes mellitus (disorder) |
|  | 421164006 | Hypoglycemic coma in type II diabetes mellitus (disorder) |
|  | 421165007 | Diabetic oculopathy associated with type I diabetes mellitus (disorder) |
|  | 421256007 | Ophthalmic complication of malnutrition-related diabetes mellitus (disorder) |
|  | 421305000 | Persistent microalbuminuria associated with type I diabetes mellitus (disorder) |
|  | 421326000 | Neurologic disorder associated with type II diabetes mellitus (disorder) |
|  | 421365002 | Peripheral circulatory disorder associated with type I diabetes mellitus (disorder) |
|  | 421437000 | Hypoglycemic coma in type I diabetes mellitus (disorder) |
|  | 421468001 | Neurological disorder associated with type I diabetes mellitus (disorder) |
|  | 421631007 | Gangrene associated with type II diabetes mellitus (disorder) |
|  | 421707005 | Polyneuropathy associated with type II diabetes mellitus (disorder) |
|  | 421725003 | Hypoglycemic coma in diabetes mellitus (disorder) |
|  | 421750000 | Ketoacidosis in type II diabetes mellitus (disorder) |
|  | 421779007 | Exudative maculopathy associated with type II diabetes mellitus (disorder) |
|  | 421847006 | Ketoacidotic coma in type II diabetes mellitus (disorder) |
|  | 421893009 | Renal disorder associated with type I diabetes mellitus (disorder) |
|  | 421895002 | Peripheral circulatory disorder associated with diabetes mellitus (disorder) |
|  | 421920002 | Diabetic cataract associated with type I diabetes mellitus (disorder) |
|  | 421966007 | Non-ketotic non-hyperosmolar coma associated with diabetes mellitus (disorder) |
|  | 421986006 | Persistent proteinuria associated with type II diabetes mellitus (disorder) |
|  | 422014003 | Disorder associated with type II diabetes melliltus (disorder) |
|  | 422034002 | Diabetic retinopathy associated with type II diabetes mellitus (disorder) |
|  | 422088007 | Neurologic disorder associated with diabetes mellitus (disorder) |
|  | 422099009 | Diabetic oculopathy associated with type II diabetes mellitus (disorder) |
|  | 422126006 | Hyperosmolar coma associated with diabetes mellitus (disorder) |
|  | 422166005 | Peripheral circulatory disorder associated with type II diabetes mellitus (disorder) |
|  | 422183001 | Skin ulcer associated with diabetes mellitus (disorder) |
|  | 422228004 | Multiple complications of type I diabetes mellitus (disorder) |
|  | 422275004 | Gangrene associated with diabetes mellitus (disorder) |
|  | 422297002 | Polyneuropathy associated with type I diabetes mellitus (disorder) |
|  | 423263001 | Diabetic autonomic neuropathy associated with type 2 diabetes mellitus (disorder) |
|  | 424736006 | Diabetic peripheral neuropathy (disorder) |
|  | 424989000 | Diabetic gastroparesis associated with type 2 diabetes mellitus (disorder) |
|  | 425159004 | Diabetic gastroparesis associated with type 1 diabetes mellitus (disorder) |
|  | 425442003 | Diabetic autonomic neuropathy associated with type 1 diabetes mellitus (disorder) |
|  | 425455002 | Diabetic glomerulonephritis (disorder) |
|  | 426705001 | Diabetes mellitus associated with cystic fibrosis (disorder) |
|  | 426875007 | Latent autoimmune diabetes mellitus in adult (disorder) |
|  | 426907004 | Small vessel disease due to type 1 diabetes mellitus (disorder) |
|  | 427027005 | Amyotrophy due to type 2 diabetes mellitus (disorder) |
|  | 427134009 | Small vessel disease due to type 2 diabetes mellitus (disorder) |
|  | 427571000 | Amyotrophy due to type 1 diabetes mellitus (disorder) |
|  | 427943001 | Diabetic ophthalmoplegia (disorder) |
|  | 428007007 | Erectile dysfunction associated with type 2 diabetes mellitus (disorder) |
|  | 428274007 | Dietary education for type II diabetes mellitus (procedure) |
|  | 428896009 | Hyperosmolality due to uncontrolled type 1 diabetes mellitus (disorder) |
|  | 429094000 | Dietary education for type I diabetes mellitus (procedure) |
|  | 429729007 | Diabetic education completed (situation) |
|  | 433874008 | History of diabetic peripheral angiopathy (situation) |
|  | 441628001 | Multiple complications due to diabetes mellitus |
|  | 441656006 | Hyperglycaemic crisis in diabetes mellitus |
|  | 443694000 | Type II diabetes mellitus uncontrolled (finding) |
|  | 444073006 | Type I diabetes mellitus uncontrolled (finding) |
|  | 445170001 | Macroalbuminuric diabetic nephropathy (disorder) |
|  | 472969004 | History of diabetes mellitus type 2 (situation) |
|  | 472970003 | History of diabetes mellitus type 1 (situation) |
|  | 472972006 | History of autosomal dominant diabetes mellitus |
|  | 473134007 | Symptomatic diabetic peripheral neuropathy absent |
|  | 473180009 | Discussion about diabetic ketoacidosis in pregnancy (procedure) |
|  | 609561005 | Maturity-onset diabetes of the young (disorder) |
|  | 609562003 | Maturity onset diabetes of the young, type 1 (disorder) |
|  | 700414001 | Education about diabetes and driving |
|  | 706891008 | Neuropathy due to brittle type I diabetes mellitus |
|  | 706894000 | Retinopathy due to unstable diabetes mellitus type 1 (disorder) |
|  | 707221002 | Diabetic glomerulosclerosis |
|  | 712882000 | Diabetic autonomic neuropathy due to type 1 diabetes mellitus |
|  | 712883005 | Diabetic autonomic neuropathy due to type 2 diabetes mellitus |
|  | 713130008 | Assessment of diabetic foot ulcer |
|  | 713150007 | Diabetic foot ulcer prevention (regime/therapy) |
|  | 713457002 | Neovascular glaucoma due to diabetes mellitus (disorder) |
|  | 713702000 | Gastroparesis due to type 1 diabetes mellitus |
|  | 713703005 | Gastroparesis due to type 2 diabetes mellitus (disorder) |
|  | 713704004 | Gastroparesis due to diabetes mellitus (disorder) |
|  | 713705003 | Polyneuropathy due to type 1 diabetes mellitus |
|  | 713706002 | Polyneuropathy due to type 2 diabetes mellitus |
|  | 715759002 | Provision of written information about diabetes and hypertension (procedure) |
|  | 715879000 | Provision of written information about diabetes and high cholesterol (procedure) |
|  | 715894001 | Provision of written information about diabetes mellitus (procedure) |
|  | 719216001 | Hypoglycemic coma co-occurrent and due to diabetes mellitus type II (disorder) |
|  | 719943000 | Provision of written information about diabetes and high hemoglobin A1c level (procedure) |
|  | 721283000 | Acidosis due to type 1 diabetes mellitus (disorder) |
|  | 721284006 | Acidosis due to type 2 diabetes mellitus (disorder) |
|  | 722161008 | Diabetic retinal eye exam (procedure) |
|  | 723074006 | Renal papillary necrosis due to diabetes mellitus (disorder) |
|  | 724136006 | Diabetic mastopathy |
|  | 724810001 | Radiculoplexoneuropathy due to diabetes mellitus (disorder) |
|  | 724876003 | Lesion of skin co-occurrent and due to diabetes mellitus (disorder) |
|  | 724997001 | Lumbosacral plexopathy co-occurrent and due to diabetes mellitus (disorder) |
|  | 735200002 | Absence of lower limb due to diabetes mellitus (disorder) |
|  | 735537007 | Hyperosmolar hyperglycemic coma due to diabetes mellitus without ketoacidosis (disorder) |
|  | 735538002 | Lactic acidosis co-occurrent and due to diabetes mellitus (disorder) |
|  | 735539005 | Metabolic acidosis co-occurrent and due to diabetes mellitus (disorder) |
|  | 739681000 | Diabetic oculopathy due to type I diabetes mellitus |
|  | 762489000 | Acute complication with diabetes mellitus |
|  | 768792007 | Cataract of right eye co-occurrent and due to diabetes mellitus |
|  | 768793002 | Diabetic cataract of left eye |
|  | 768794008 | Bilateral diabetic cataracts |
|  | 768797001 | Iritis of right eye co-occurrent and due to diabetes mellitus |
|  | 768798006 | Iritis of left eye co-occurrent and due to diabetes mellitus |
|  | 768799003 | Iritis of bilateral eyes co-occurrent and due to diabetes mellitus (disorder) |
|  | 769181007 | Preproliferative retinopathy of right eye co-occurrent and due to diabetes mellitus (disorder) |
|  | 769182000 | Preproliferative diabetic retinopathy of left eye |
|  | 769183005 | Mild nonproliferative diabetic retinopathy of right eye |
|  | 769184004 | Mild nonproliferative retinopathy of left eye |
|  | 769185003 | Moderate non-proliferative diabetic retinopathy of right eye |
|  | 769186002 | Moderate nonproliferative diabetic retinopathy of left eye |
|  | 769187006 | Severe nonproliferative diabetic retinopathy of right eye |
|  | 769188001 | Severe nonproliferative retinopathy of left eye co-occurrent and due to diabetes mellitus (disorder) |
|  | 769190000 | Very severe nonproliferative diabetic retinopathy of right eye |
|  | 769191001 | Very severe nonproliferative diabetic retinopathy of left eye |
|  | 769217008 | Diabetic macular edema of right eye |
|  | 769218003 | Macular oedema of left eye co-occurrent and due to diabetes mellitus |
|  | 769219006 | Macular edema due to type 1 diabetes mellitus |
|  | 769220000 | Macular edema co-occurrent and due to type 2 diabetes mellitus |
|  | 769221001 | Clinically significant macular edema of right eye co-occurrent and due to diabetes mellitus (disorder) |
|  | 769222008 | Clinically significant macular oedema of left eye co-occurrent and due to diabetes mellitus |
|  | 769244003 | Diabetic maculopathy of right eye |
|  | 769245002 | Disorder of left macula co-occurrent and due to diabetes mellitus (disorder) |
|  | 770094004 | Cervical radiculoplexus neuropathy co-occurrent and due to diabetes mellitus (disorder) |
|  | 770095003 | Cranial nerve palsy due to diabetes mellitus |
|  | 770096002 | Erectile dysfunction co-occurrent and due to diabetes mellitus (disorder) |
|  | 770097006 | Clinically significant macular edema co-occurrent and due to diabetes mellitus (disorder) |
|  | 770098001 | Cranial nerve palsy with type 1 diabetes mellitus |
|  | 770323005 | Retinal edema due to diabetes mellitus |
|  | 770324004 | Retinal ischemia due to diabetes mellitus |
|  | 770361008 | Vitreous hemorrhage of right eye co-occurrent and due to diabetes mellitus |
|  | 770362001 | Vitreous haemorrhage of left eye with diabetes mellitus |
|  | 770581008 | Diabetic retinal microaneurysm of right eye |
|  | 770582001 | Retinal microaneurysm of left eye co-occurrent and due to diabetes mellitus |
|  | 770599000 | Retinal venous beading of right eye co-occurrent and due to diabetes mellitus |
|  | 770600002 | Retinal venous beading of left eye with diabetes mellitus |
|  | 770765001 | Proliferative diabetic retinopathy of right eye |
|  | 770766000 | Proliferative diabetic retinopathy of left eye |
|  | 201000119106 | Disorder associated with well controlled type 2 diabetes mellitus |
|  | 691000119103 | Erectile dysfunction associated with type I diabetes mellitus |
|  | 701000119103 | Mixed hyperlipidaemia associated with type II diabetes mellitus |
|  | 711000119100 | Diabetic stage 5 chronic renal impairment associated with type 2 diabetes mellitus |
|  | 721000119107 | Diabetic stage 4 chronic renal impairment associated with type 2 diabetes mellitus |
|  | 731000119105 | Chronic kidney disease stage 3 associated with type 2 diabetes mellitus |
|  | 741000119101 | Chronic kidney disease stage 2 associated with type 2 diabetes mellitus |
|  | 751000119104 | Chronic kidney disease stage 1 associated with type 2 diabetes mellitus |
|  | 761000119102 | Diabetic dyslipidemia associated with type 2 diabetes mellitus |
|  | 771000119108 | Chronic renal impairment associated with type II diabetes mellitus |
|  | 781000119106 | Diabetic neuropathic arthropathy associated with type 2 diabetes mellitus |
|  | 791000119109 | Angina associated with type II diabetes mellitus |
|  | 1491000119102 | Diabetic vitreous haemorrhage associated with type II diabetes mellitus |
|  | 1501000119109 | Proliferative diabetic retinopathy associated with type II diabetes mellitus |
|  | 1511000119107 | Diabetic peripheral neuropathy associated with type II diabetes mellitus |
|  | 1521000119100 | Diabetic foot ulcer associated with type 2 diabetes mellitus |
|  | 1531000119102 | Diabetic dermopathy associated with diabetes mellitus type 2 |
|  | 1541000119106 | Diabetic skin ulcer associated with type 2 diabetes mellitus |
|  | 1551000119108 | Nonproliferative diabetic retinopathy associated with type II diabetes mellitus |
|  | 1561000119105 | Diabetic peripheral neuropathy associated with type I diabetes mellitus |
|  | 1571000119104 | Mixed hyperlipidemia associated with type 1 diabetes mellitus (disorder) |
|  | 4551000175107 | Diabetic foot exam not done |
|  | 4581000175103 | Diabetic retinal eye exam not done (situation) |
|  | 12811000119100 | Complication due to diabetes mellitus type 2 |
|  | 18521000119106 | Microalbuminuria due to type 1 diabetes mellitus |
|  | 28331000119107 | Retinal oedema due to type 2 diabetes mellitus |
|  | 31211000119101 | Peripheral vascular disease due to type I diabetes |
|  | 41911000119107 | Glaucoma due to type 2 diabetes mellitus (disorder) |
|  | 60951000119105 | Blindness due to type 2 diabetes mellitus |
|  | 60961000119107 | Nonproliferative diabetic retinopathy due to type 1 diabetes mellitus |
|  | 60971000119101 | Proliferative retinopathy due to type 1 diabetes mellitus |
|  | 60991000119100 | Blindness due to type 1 diabetes mellitus (disorder) |
|  | 71421000119105 | Hypertension in chronic kidney disease due to type 2 diabetes mellitus (disorder) |
|  | 71441000119104 | Nephrotic syndrome due to type 2 diabetes mellitus |
|  | 71701000119105 | Hypertension in chronic kidney disease due to type 1 diabetes mellitus |
|  | 71721000119101 | Nephrotic syndrome due to type 1 diabetes mellitus |
|  | 71771000119100 | Type 1 diabetes mellitus with neuropathic arthropathy |
|  | 71791000119104 | Peripheral neuropathy due to type 1 diabetes mellitus |
|  | 72021000119109 | Diabetic dermopathy due to type 1 diabetes mellitus |
|  | 72031000119107 | Severe malnutrition due to type 1 diabetes mellitus (disorder) |
|  | 72041000119103 | Osteomyelitis due to type 1 diabetes mellitus (disorder) |
|  | 72051000119101 | Severe malnutrition due to type 2 diabetes mellitus |
|  | 72061000119104 | Osteomyelitis due to type 2 diabetes mellitus (disorder) |
|  | 72141000119104 | Chronic ulcer of skin due to type 1 diabetes mellitus |
|  | 82541000119100 | Traction retinal detachment due to type 2 diabetes mellitus |
|  | 82551000119103 | Rubeosis iridis due to type 2 diabetes mellitus |
|  | 82571000119107 | Traction retinal detachment due to type 1 diabetes mellitus |
|  | 82581000119105 | Rubeosis iridis due to type 1 diabetes mellitus |
|  | 84361000119102 | Insulin reactive hypoglycemia in type 2 diabetes mellitus |
|  | 84371000119108 | Hypoglycemia due to type 1 diabetes mellitus |
|  | 87441000119104 | Ankle ulcer due to type 2 diabetes mellitus (disorder) |
|  | 87451000119102 | Heel AND/OR midfoot ulcer due to type 2 diabetes mellitus |
|  | 87461000119100 | Forefoot ulcer due to type 2 diabetes mellitus |
|  | 87471000119106 | Ankle ulcer due to type 1 diabetes mellitus (disorder) |
|  | 87481000119109 | Heel AND/OR midfoot ulcer due to type 1 diabetes mellitus |
|  | 87491000119107 | Forefoot ulcer due to type 1 diabetes mellitus |
|  | 87921000119104 | Cranial nerve palsy due to type 2 diabetes mellitus |
|  | 90721000119101 | Chronic kidney disease stage 1 due to type 1 diabetes mellitus (disorder) |
|  | 90731000119103 | Chronic kidney disease stage 2 due to type 1 diabetes mellitus |
|  | 90741000119107 | Chronic kidney disease stage 3 due to type 1 diabetes mellitus |
|  | 90751000119109 | Chronic kidney disease stage 4 due to type 1 diabetes mellitus |
|  | 90761000119106 | Chronic kidney disease stage 5 due to type 1 diabetes mellitus (disorder) |
|  | 90771000119100 | End stage renal disease on dialysis due to type 1 diabetes mellitus (disorder) |
|  | 90781000119102 | Microalbuminuria due to type 2 diabetes mellitus (disorder) |
|  | 90791000119104 | End stage renal disease on dialysis due to type 2 diabetes mellitus (disorder) |
|  | 96441000119101 | Chronic kidney disease due to type 1 diabetes mellitus |
|  | 97331000119101 | Macular edema and retinopathy due to type 2 diabetes mellitus (disorder) |
|  | 97341000119105 | Proliferative retinopathy with retinal oedema due to type 2 diabetes mellitus |
|  | 97621000119107 | Stasis ulcer due to type 2 diabetes mellitus (disorder) |
|  | 102621000119101 | Skin ulcer due to type 2 diabetes mellitus |
|  | 102781000119107 | Sensory neuropathy due to type 1 diabetes mellitus (disorder) |
|  | 103981000119101 | Proliferative diabetic retinopathy following surgery |
|  | 104941000119109 | Retinal ischemia due to type 1 diabetes mellitus |
|  | 104951000119106 | Vitreous haemorrhage due to type 1 diabetes mellitus |
|  | 104961000119108 | Retinal ischemia due to type 2 diabetes mellitus |
|  | 108781000119105 | Neuropathic ulcer of midfoot AND/OR heel due to type 2 diabetes mellitus (disorder) |
|  | 108791000119108 | Suspected glaucoma due to type 2 diabetes mellitus (situation) |
|  | 109171000119104 | Retinal oedema due to type 1 diabetes mellitus |
|  | 110141000119100 | Ulcer of lower limb due to type 1 diabetes mellitus (disorder) |
|  | 110171000119107 | Leg ulcer due to type 2 diabetes mellitus |
|  | 110181000119105 | Peripheral sensory neuropathy due to type 2 diabetes mellitus |
|  | 111231000119109 | Dyslipidemia with high density lipoprotein below reference range and triglyceride above reference range due to type 2 diabetes mellitus |
|  | 119831000119106 | Hypoglycemia unawareness in type 2 diabetes mellitus (disorder) |
|  | 120711000119108 | Hypoglycaemic unawareness in type 1 diabetes mellitus |
|  | 120731000119103 | Hypoglycemia due to type 2 diabetes mellitus (disorder) |
|  | 127991000119101 | Hypertension concurrent and due to end stage renal disease on dialysis due to type 2 diabetes mellitus |
|  | 128001000119105 | Hypertension concurrent and due to end stage renal disease on dialysis due to type 1 diabetes mellitus (disorder) |
|  | 137931000119102 | Hyperlipidemia due to type 2 diabetes mellitus |
|  | 137941000119106 | Hyperlipidemia due to type 1 diabetes mellitus |
|  | 138881000119106 | Mild nonproliferative retinopathy due to type 1 diabetes mellitus |
|  | 138891000119109 | Moderate nonproliferative retinopathy due to type 1 diabetes mellitus (disorder) |
|  | 138911000119106 | Mild nonproliferative retinopathy due to type 2 diabetes mellitus |
|  | 138921000119104 | Moderate nonproliferative retinopathy due to type 2 diabetes mellitus (disorder) |
|  | 140101000119109 | Hypertension in chronic kidney disease stage 5 due to type 2 diabetes mellitus |
|  | 140111000119107 | Hypertension in chronic kidney disease stage 4 due to type 2 diabetes mellitus (disorder) |
|  | 140121000119100 | Hypertension in chronic kidney disease stage 3 due to type 2 diabetes mellitus |
|  | 140131000119102 | Hypertension in chronic kidney disease stage 2 due to type 2 diabetes mellitus (disorder) |
|  | 140381000119104 | Neuropathic toe ulcer due to type 2 diabetes mellitus (disorder) |
|  | 140391000119101 | Ulcer of toe due to type 2 diabetes mellitus (disorder) |
|  | 140521000119107 | Ischemic foot ulcer due to type 2 diabetes mellitus (disorder) |
|  | 140531000119105 | Neuropathic foot ulcer due to type 2 diabetes mellitus (disorder) |
|  | 142881000119105 | History of nocturnal hypoglycaemia |
|  | 157141000119108 | Proteinuria due to type 2 diabetes mellitus |
|  | 164881000119109 | Foot ulcer due to type 1 diabetes mellitus (disorder) |
|  | 243421000119104 | Proteinuria due to type 1 diabetes mellitus (disorder) |
|  | 324301000119107 | Arthropathy due to metabolic disorder (disorder) |
|  | 367991000119101 | Hyperglycemia due to type 1 diabetes mellitus |
|  | 368051000119109 | Hyperglycemia due to type 2 diabetes mellitus (disorder) |
|  | 368521000119107 | Disorder of nerve co-occurrent and due to type 1 diabetes mellitus (disorder) |
|  | 368551000119104 | Dyslipidemia due to type 1 diabetes mellitus (disorder) |
|  | 368561000119102 | Hyperosmolarity due to type 1 diabetes mellitus (disorder) |
|  | 368581000119106 | Neuropathy due to type 2 diabetes mellitus (disorder) |
|  | 368591000119109 | Cheiropathy due to type 2 diabetes mellitus (disorder) |
|  | 368601000119102 | Hyperosmolar coma due to secondary diabetes mellitus |
|  | 368711000119106 | Mild nonproliferative retinopathy due to secondary diabetes mellitus |
|  | 368721000119104 | Non-proliferative retinopathy due to secondary diabetes mellitus (disorder) |
|  | 368741000119105 | Moderate non-proliferative retinopathy due to secondary diabetes mellitus (disorder) |
|  | 430801000124103 | Proliferative retinopathy (disorder) |
|  | 10656231000119100 | Skin ulcer of toe due to diabetes mellitis type 1 (disorder) |
|  | 10656271000119100 | Skin ulcer of toe due to diabetes mellitis type 2 |
|  | 10660471000119100 | Ulcer of left foot co-occurrent and due to diabetes mellitus type 2 (disorder) |
|  | 10661671000119100 | Ulcer of right foot co-occurrent and due to diabetes mellitus type 2 (disorder) |
|  | 10995761000119100 | History of diabetic foot ulcer (situation) |
|  | 1481000119100 | Diabetes mellitus type 2 without retinopathy (disorder) |
|  | 164971000119101 | Type 2 diabetes mellitus controlled by diet (finding) |
|  | 24481000000101 | Type 2 diabetic on diet only (finding) |
|  | 335621000000101 | Maternally inherited diabetes mellitus (disorder) |
|  | 359642000 | Diabetes mellitus type 2 in nonobese (disorder) |
|  | 445353002 | Brittle type II diabetes mellitus (finding) |
|  | 703138006 | Type II diabetes mellitus in remission (disorder) |
|  | 81531005 | Diabetes mellitus type 2 in obese (disorder) |
|  | 237627000 | Pregnancy and type 2 diabetes mellitus (disorder) |
|  | 24471000000103 | Type 2 diabetic on insulin (finding) |
|  | 279321000000104 | Diabetes type 2 review (regime/therapy) |
|  | 444110003 | Type II diabetes mellitus well controlled (finding) |
|  | 609567009 | Pre-existing type 2 diabetes mellitus in pregnancy (disorder) |
|  | 754121000000107 | Type II diabetic dietary review (regime/therapy) |
|  | 724205009 | Laminopathy type Decaudain Vigouroux (disorder) |
|  | 1067201000000100 | Eating disorder co-occurrent with diabetes mellitus type 1 (disorder) |
|  | 111307005 | Leprechaunism syndrome (disorder) |
|  | 123763000 | Houssay's syndrome (disorder) |
|  | 190416008 | Steroid-induced diabetes mellitus without complication (disorder) |
|  | 198131000000101 | Hypoglycaemic warning good (disorder) |
|  | 20678000 | Extreme insulin resistance with acanthosis nigricans, hirsutism AND autoantibodies to the insulin receptors (disorder) |
|  | 237600004 | Malnutrition-related diabetes mellitus - fibrocalculous (disorder) |
|  | 237601000 | Secondary endocrine diabetes mellitus (disorder) |
|  | 237610008 | Acrorenal field defect, ectodermal dysplasia, and lipoatrophic diabetes (disorder) |
|  | 237612000 | Photomyoclonus, diabetes mellitus, deafness, nephropathy and cerebral dysfunction (disorder) |
|  | 237618001 | Insulin-dependent diabetes mellitus secretory diarrhea syndrome (disorder) |
|  | 237632004 | Hypoglycemic event in diabetes (disorder) |
|  | 267467004 | Diabetes mellitus (& [ketoacidosis]) (disorder) |
|  | 2751001 | Fibrocalculous pancreatic diabetes (disorder) |
|  | 279291000000109 | Diabetes type 1 review (regime/therapy) |
|  | 33559001 | Pineal hyperplasia AND diabetes mellitus syndrome (disorder) |
|  | 368171000119104 | Dermatitis due to drug induced diabetes mellitus (disorder) |
|  | 385041000000108 | Diabetes mellitus with multiple complications (disorder) |
|  | 385051000000106 | Pre-existing diabetes mellitus (disorder) |
|  | 408539000 | Insulin autoimmune syndrome (disorder) |
|  | 413183008 | Diabetes mellitus caused by non-steroid drugs without complication (disorder) |
|  | 413184002 | Fibrocalculous pancreatopathy without complication (disorder) |
|  | 609564002 | Pre-existing type 1 diabetes mellitus in pregnancy (disorder) |
|  | 609565001 | Permanent neonatal diabetes mellitus (disorder) |
|  | 609568004 | Diabetes mellitus due to genetic defect in beta cell function (disorder) |
|  | 609571007 | Maturity-onset diabetes of the young, type 4 (disorder) |
|  | 609572000 | Maturity-onset diabetes of the young, type 5 (disorder) |
|  | 609575003 | Maturity-onset diabetes of the young, type 8 (disorder) |
|  | 703136005 | Diabetes mellitus in remission (disorder) |
|  | 721973006 | Lipodystrophy, intellectual disability, deafness syndrome (disorder) |
|  | 722206009 | Pancreatic hypoplasia, diabetes mellitus, congenital heart disease syndrome (disorder) |
|  | 722454003 | Intellectual disability, craniofacial dysmorphism, hypogonadism, diabetes mellitus syndrome (disorder) |
|  | 734022008 | Wolfram-like syndrome (disorder) |
|  | 737212004 | Diabetes mellitus caused by chemical (disorder) |
|  | 763325000 | Insulin resistance (disorder) |
|  | 771571000000102 | History of secondary diabetes mellitus (situation) |
|  | 894741000000107 | Hypoglycaemic warning absent (disorder) |
|  | 105401000119101 | Diabetes mellitus due to pancreatic injury (disorder) |
|  | 106281000119103 | Pre-existing diabetes mellitus in mother complicating childbirth (disorder) |
|  | 10754881000119100 | Diabetes mellitus in mother complicating childbirth (disorder) |
|  | 1102351000000100 | Ketosis-prone diabetes mellitus (disorder) |
|  | 112991000000101 | Lipoatrophic diabetes mellitus without complication (disorder) |
|  | 190407009 | Malnutrition-related diabetes mellitus with renal complications (disorder) |
|  | 190412005 | Malnutrition-related diabetes mellitus without complications (disorder) |
|  | 190447002 | Steroid-induced diabetes (disorder) |
|  | 198121000000103 | Hypoglycaemic warning impaired (disorder) |
|  | 23045005 | Insulin dependent diabetes mellitus type IA (disorder) |
|  | 237608006 | Lipodystrophy, partial, with Rieger anomaly, short stature, and insulinopenic diabetes mellitus (disorder) |
|  | 237613005 | Hyperproinsulinemia (disorder) |
|  | 237616002 | Hypogonadism, diabetes mellitus, alopecia, mental retardation and electrocardiographic abnormalities (disorder) |

|  | 237617006 | Megaloblastic anemia, thiamine-responsive, with diabetes mellitus and sensorineural deafness (disorder) |
| --- | --- | --- |
|  | 237619009 | Diabetes-deafness syndrome maternally transmitted (disorder) |
|  | 237622006 | Poor glycemic control (disorder) |
|  | 237650006 | Insulin resistance in diabetes (disorder) |
|  | 237651005 | Insulin resistance - type A (disorder) |
|  | 237652003 | Insulin resistance - type B (disorder) |
|  | 24203005 | Extreme insulin resistance with acanthosis nigricans, hirsutism AND abnormal insulin receptors (disorder) |
|  | 267471001 | Diabetes + eye manifestation (& [cataract] or [retinopathy]) (disorder) |
|  | 28032008 | Insulin dependent diabetes mellitus type IB (disorder) |
|  | 284449005 | Congenital total lipodystrophy (disorder) |
|  | 303059007 | Postpancreatectomy hypoinsulinemia (disorder) |
|  | 31321000119102 | Diabetes mellitus type 1 without retinopathy (disorder) |
|  | 367261000119100 | Hyperosmolarity co-occurrent and due to drug induced diabetes mellitus (disorder) |
|  | 408540003 | Diabetes mellitus caused by non-steroid drugs (disorder) |
|  | 427089005 | Diabetes mellitus due to cystic fibrosis (disorder) |
|  | 42954008 | Diabetes mellitus associated with receptor abnormality (disorder) |
|  | 444074000 | Type I diabetes mellitus well controlled (finding) |
|  | 445260006 | Posttransplant diabetes mellitus (disorder) |
|  | 530558861000132000 | Atypical diabetes mellitus (disorder) |
|  | 532411000000102 | Diabetes mellitus, adult onset, with no mention of complication (disorder) |
|  | 5368009 | Drug-induced diabetes mellitus (disorder) |
|  | 57886004 | Protein-deficient diabetes mellitus (disorder) |
|  | 59079001 | Diabetes mellitus associated with hormonal etiology (disorder) |
|  | 5969009 | Diabetes mellitus associated with genetic syndrome (disorder) |
|  | 609563008 | Pre-existing diabetes mellitus in pregnancy (disorder) |
|  | 609566000 | Pregnancy and type 1 diabetes mellitus (disorder) |
|  | 609569007 | Diabetes mellitus due to genetic defect in insulin action (disorder) |
|  | 609570008 | Maturity-onset diabetes of the young, type 3 (disorder) |
|  | 609573005 | Maturity-onset diabetes of the young, type 6 (disorder) |
|  | 609574004 | Maturity-onset diabetes of the young, type 7 (disorder) |
|  | 609576002 | Maturity-onset diabetes of the young, type 9 (disorder) |
|  | 609577006 | Maturity-onset diabetes of the young, type 10 (disorder) |
|  | 609578001 | Maturity-onset diabetes of the young, type 11 (disorder) |
|  | 658011000000104 | Diabetes mellitus with other specified manifestation (disorder) |
|  | 703137001 | Type I diabetes mellitus in remission (disorder) |
|  | 70694009 | Diabetes mellitus AND insipidus with optic atrophy AND deafness (disorder) |
|  | 709147009 | Gingivitis co-occurrent with diabetes mellitus (disorder) |
|  | 716362006 | Gingival disease co-occurrent with diabetes mellitus (disorder) |
|  | 720519003 | Atherosclerosis, deafness, diabetes, epilepsy, nephropathy syndrome (disorder) |
|  | 724067006 | Permanent neonatal diabetes mellitus with cerebellar agenesis syndrome (disorder) |
|  | 733072002 | Alaninuria, microcephaly, dwarfism, enamel hypoplasia, diabetes mellitus syndrome (disorder) |
|  | 735199000 | History of diabetes related lower limb amputation (situation) |
|  | 754101000000103 | Type I diabetic dietary review (regime/therapy) |
|  | 754461000000105 | Referral to type I diabetes structured education programme (procedure) |
|  | 75682002 | Diabetes mellitus caused by insulin receptor antibodies (disorder) |
|  | 773001000000103 | Symptomatic diabetic peripheral neuropathy (disorder) |
|  | 775841000000109 | Diabetic retinopathy detected by national screening programme (disorder) |
|  | 80660001 | Mauriac's syndrome (disorder) |
|  | 83728000 | Polyglandular autoimmune syndrome, type 2 (disorder) |
|  | 91352004 | Diabetes mellitus due to structurally abnormal insulin (disorder) |
|  | 10154911000001100 | Glimepiride 1mg tablets (Alpharma Ltd) (product) |
|  | 10155111000001100 | Glimepiride 2mg tablets (Alpharma Ltd) (product) |
|  | 10155311000001100 | Glimepiride 3mg tablets (Alpharma Ltd) (product) |
|  | 10155511000001100 | Glimepiride 4mg tablets (Alpharma Ltd) (product) |
|  | 10272611000001100 | Glimepiride 1mg tablets (Unichem Plc) (product) |
|  | 10273811000001100 | Glimepiride 1mg tablets (A A H Pharmaceuticals Ltd) (product) |
|  | 10274311000001100 | Glimepiride 3mg tablets (A A H Pharmaceuticals Ltd) (product) |
|  | 10274511000001100 | Glimepiride 4mg tablets (A A H Pharmaceuticals Ltd) (product) |
|  | 10293511000001100 | Metformin 850mg tablets (Relonchem Ltd) (product) |
|  | 10377311000001100 | Glibenclamide 2.5mg tablets (Arrow Generics Ltd) (product) |
|  | 10377711000001100 | Glibenclamide 5mg tablets (Arrow Generics Ltd) (product) |
|  | 10378111000001100 | Gliclazide 80mg tablets (Arrow Generics Ltd) (product) |
|  | 10390211000001100 | Metformin 500mg tablets (Arrow Generics Ltd) (product) |
|  | 10390811000001100 | Metformin 850mg tablets (Arrow Generics Ltd) (product) |
|  | 10437311000001100 | Glimepiride 4mg tablets (Niche Generics Ltd) (product) |
|  | 10458811000001100 | Actos 15mg tablets (PI) (Dowelhurst Ltd) (product) |
|  | 10465111000001100 | Avandia 8mg tablets (PI) (Dowelhurst Ltd) (product) |
|  | 10486511000001100 | NovoNorm 500microgram tablets (PI) (Dowelhurst Ltd) (product) |
|  | 10487311000001100 | NovoNorm 2mg tablets (PI) (Dowelhurst Ltd) (product) |
|  | 10488111000001100 | Avandamet 4mg/1000mg tablets (PI) (Waymade Ltd) (product) |
|  | 10517711000001100 | Glimepiride 1mg tablets (Winthrop Pharmaceuticals UK Ltd) (product) |
|  | 10518111000001100 | Glimepiride 3mg tablets (Winthrop Pharmaceuticals UK Ltd) (product) |
|  | 10518311000001100 | Glimepiride 4mg tablets (Winthrop Pharmaceuticals UK Ltd) (product) |
|  | 10539211000001100 | Glimepiride 1mg tablets (PI) (Waymade Ltd) (product) |
|  | 10539511000001100 | Glimepiride 2mg tablets (PI) (Waymade Ltd) (product) |
|  | 10539711000001100 | Glimepiride 3mg tablets (PI) (Waymade Ltd) (product) |
|  | 10540011000001100 | Glimepiride 4mg tablets (PI) (Waymade Ltd) (product) |
|  | 10542811000001100 | Humalog 100units/ml solution for injection 10ml vials (PI) (Waymade Ltd) (product) |
|  | 10543211000001100 | NovoRapid FlexPen 100units/ml solution for injection 3ml pre-filled pens (Waymade Healthcare Plc) (product) |
|  | 10548211000001100 | Lantus 100units/ml solution for injection 10ml vials (PI) (Waymade Ltd) (product) |
|  | 10548611000001100 | Lantus 100units/ml solution for injection 3ml pre-filled OptiSet pens (Waymade Healthcare Plc) (product) |
|  | 10549011000001100 | Lantus 100units/ml solution for injection 3ml cartridges (PI) (Waymade Ltd) (product) |
|  | 10617511000001100 | Glimepiride 2mg tablets (Somex Pharma) (product) |
|  | 10689011000001100 | Glimepiride 2mg tablets (Pliva Pharma Ltd) (product) |
|  | 10689211000001100 | Glimepiride 3mg tablets (Pliva Pharma Ltd) (product) |
|  | 10741611000001100 | Metsol 500mg/5ml oral solution (Orbis Consumer Products Ltd) (product) |
|  | 10750111000001100 | Metformin 500mg/5ml oral solution sugar free (product) |
|  | 10832711000001100 | Metformin 500mg/5ml oral solution sugar free (Unichem Plc) (product) |
|  | 10898411000001100 | Apidra 100units/ml solution for injection 3ml OptiClik cartridges (sanofi-aventis) (product) |
|  | 10922311000001100 | Competact 15mg/850mg tablets (Takeda UK Ltd) (product) |
|  | 10956911000001100 | Prandin 1mg tablets (Novo Nordisk Ltd) (product) |
|  | 10963611000001100 | Metformin 500mg/5ml oral solution sugar free (A A H Pharmaceuticals Ltd) (product) |
|  | 11018411000001100 | Glimepiride 4mg tablets (Dr Reddy's Labs) (product) |
|  | 11018911000001100 | Glimepiride 2mg tablets (Dr Reddy's Labs) (product) |
|  | 11019211000001100 | Glimepiride 1mg tablets (Dr Reddy's Labs) (product) |
|  | 11114611000001100 | Levemir Penfill 100units/ml solution for injection 3ml cartridges (Waymade Ltd) (product) |
|  | 11116211000001100 | Pork Mixtard 30 100units/ml suspension for injection 10ml vials (Waymade Ltd) (product) |
|  | 11118911000001100 | Hypurin Bovine Neutral 100units/ml solution for injection 10ml vials (Waymade Ltd) (product) |
|  | 11121911000001100 | Velosulin 100units/ml solution for injection 10ml vials (Waymade Ltd) (product) |
|  | 11124011000001100 | Mixtard 30 100units/ml suspension for injection 10ml vials (Waymade Ltd) (product) |
|  | 11124411000001100 | Mixtard 30 InnoLet 100units/ml suspension for injection 3ml pre-filled pens (Waymade Healthcare Plc) (product) |
|  | 11134711000001100 | Insulatard 100units/ml suspension for injection 10ml vials (Waymade Ltd) (product) |
|  | 11135611000001100 | Mixtard 10 Penfill 100units/ml suspension for injection 3ml cartridges (Waymade Ltd) (product) |
|  | 11148111000001100 | Levemir InnoLet 100units/ml solution for injection 3ml pre-filled pens (Novo Nordisk Ltd) (product) |
|  | 11149211000001100 | Glimepiride 1mg tablets (Teva UK Ltd) (product) |
|  | 11149411000001100 | Glimepiride 2mg tablets (Teva UK Ltd) (product) |
|  | 11149611000001100 | Glimepiride 3mg tablets (Teva UK Ltd) (product) |
|  | 11342811000001100 | Glimepiride 1mg tablets (Sandoz Ltd) (product) |
|  | 11343511000001100 | Glimepiride 3mg tablets (Sandoz Ltd) (product) |
|  | 11406411000001100 | Gliclazide 80mg tablets (Pliva Pharma Ltd) (product) |
|  | 11473711000001100 | Januvia 100mg tablets (Merck Sharp & Dohme Ltd) (product) |
|  | 11492211000001100 | Gliclazide 80mg tablets (Winthrop Pharmaceuticals UK Ltd) (product) |
|  | 11494611000001100 | Byetta 10micrograms/0.04ml solution for injection 2.4ml pre-filled pens (AstraZeneca UK Ltd) (product) |
|  | 11494811000001100 | Byetta 5micrograms/0.02ml solution for injection 1.2ml pre-filled pens (AstraZeneca UK Ltd) (product) |
|  | 11766011000001100 | Prandin 1mg tablets (Waymade Healthcare Plc) (product) |
|  | 11766211000001100 | Prandin 2mg tablets (Waymade Healthcare Plc) (product) |
|  | 11933011000001100 | Lantus 100units/ml solution for injection 3ml pre-filled SoloStar pens (Sanofi) (product) |
|  | 12107511000001100 | Glibenclamide 625micrograms/5ml oral solution (Special Order) (product) |
|  | 12108411000001100 | Glibenclamide 2.5mg/5ml oral suspension (Special Order) (product) |
|  | 12108711000001100 | Glibenclamide 2.5mg/5ml oral solution (Special Order) (product) |
|  | 12109911000001100 | Gliclazide 50mg/5ml oral suspension (Special Order) (product) |
|  | 12110211000001100 | Gliclazide 60mg/5ml oral suspension (Special Order) (product) |
|  | 12135411000001100 | Glibenclamide 15mg/5ml oral suspension (product) |
|  | 12135511000001100 | Glibenclamide 2.5mg/5ml oral solution (product) |
|  | 12135611000001100 | Glibenclamide 2.5mg/5ml oral suspension (product) |
|  | 12135811000001100 | Glibenclamide 625micrograms/5ml oral suspension (product) |
|  | 12135911000001100 | Gliclazide 100mg/5ml oral suspension (product) |
|  | 12136111000001100 | Gliclazide 20mg/5ml oral suspension (product) |
|  | 12136211000001100 | Gliclazide 50mg/5ml oral suspension (product) |
|  | 12144611000001100 | Apidra 100units/ml solution for injection 3ml pre-filled SoloStar pens (Sanofi) (product) |
|  | 12147311000001100 | Glimepiride 2mg tablets (Kent Pharmaceuticals Ltd) (product) |
|  | 12200111000001100 | Repaglinide 500microgram tablets (PI) (Waymade Healthcare Plc) (product) |
|  | 12200311000001100 | Repaglinide 1mg tablets (PI) (Waymade Healthcare Plc) (product) |
|  | 12200511000001100 | Repaglinide 2mg tablets (PI) (Waymade Healthcare Plc) (product) |
|  | 12798011000001100 | Metformin 1g/5ml oral solution (product) |
|  | 13100611000001100 | Avandamet 4mg/1000mg tablets (Dowelhurst Ltd) (product) |
|  | 13180611000001100 | NovoMix 30 Penfill 100units/ml suspension for injection 3ml cartridges (Dowelhurst Ltd) (product) |
|  | 13186611000001100 | Prandin 0.5mg tablets (Dowelhurst Ltd) (product) |
|  | 13318011000001100 | Tolbutamide 250mg/5ml oral suspension (Special Order) (product) |
|  | 13411611000001100 | Galvus 50mg tablets (Novartis Pharmaceuticals UK Ltd) (product) |
|  | 13413111000001100 | Metformin 850mg / Vildagliptin 50mg tablets (product) |
|  | 13456711000001100 | Nazdol MR 30mg tablets (Consilient Health Ltd) (product) |
|  | 134587001 | Gliclazide 30mg m/r tablet (product) |
|  | 134608004 | Product containing precisely nateglinide 180 milligram/1 each conventional release oral tablet (clinical drug) |
|  | 13462811000001100 | Gliclazide 30mg/5ml oral suspension (Special Order) (product) |
|  | 13466811000001100 | Gliclazide 30mg/5ml oral suspension (product) |
|  | 13511000001103 | Diabetamide 2.5mg tablets (Ashbourne Pharmaceuticals Ltd) (product) |
|  | 13549911000001100 | Gliclazide 30mg modified-release tablets (Kent Pharmaceuticals Ltd) (product) |
|  | 13613911000001100 | Gliclazide 30mg modified-release tablets (A A H Pharmaceuticals Ltd) (product) |
|  | 13626011000001100 | Niddaryl 1mg tablets (Dee Pharmaceuticals Ltd) (product) |
|  | 13627211000001100 | Niddaryl 4mg tablets (Dee Pharmaceuticals Ltd) (product) |
|  | 13720911000001100 | Glimepiride 3mg tablets (Ranbaxy (UK) Ltd) (product) |
|  | 13748611000001100 | Glucophage SR 750mg tablets (Merck Serono Ltd) (product) |
|  | 13756411000001100 | Metformin 500mg tablets (Tillomed Laboratories Ltd) (product) |
|  | 13772611000001100 | Glucophage SR 500mg tablets (Waymade Healthcare Plc) (product) |
|  | 13823711000001100 | Actos 15mg tablets (Doncaster Pharmaceuticals Ltd) (product) |
|  | 13823911000001100 | Actos 30mg tablets (Doncaster Pharmaceuticals Ltd) (product) |
|  | 13833311000001100 | Avandamet 2mg/1000mg tablets (Doncaster Pharmaceuticals Ltd) (product) |
|  | 13833511000001100 | Avandamet 4mg/1000mg tablets (Doncaster Pharmaceuticals Ltd) (product) |
|  | 13859411000001100 | Insulin human 100units/ml solution for injection 10ml vials (product) |
|  | 13864511000001100 | Diamicron 30mg MR tablets (Doncaster Pharmaceuticals Ltd) (product) |
|  | 13877811000001100 | Humalog KwikPen 100units/ml solution for injection 3ml pre-filled pens (Eli Lilly and Company Ltd) (product) |
|  | 13884911000001100 | Humalog Mix50 KwikPen 100units/ml suspension for injection 3ml pre-filled pens (Eli Lilly and Company Ltd) (product) |
|  | 14040211000001100 | Actos 15mg tablets (Sigma Pharmaceuticals Plc) (product) |
|  | 14055711000001100 | Amaryl 4mg tablets (Sigma Pharmaceuticals Plc) (product) |
|  | 14056111000001100 | Amaryl 1mg tablets (Sigma Pharmaceuticals Plc) (product) |
|  | 14061411000001100 | Amaryl 3mg tablets (Sigma Pharmaceuticals Plc) (product) |
|  | 14122111000001100 | Glimepiride 1mg tablets (Consilient Health Ltd) (product) |
|  | 14122511000001100 | Glimepiride 3mg tablets (Consilient Health Ltd) (product) |
|  | 14190111000001100 | Humulin S 100units/ml solution for injection 10ml vials (Waymade Healthcare Plc) (product) |
|  | 14190311000001100 | Humulin M3 100units/ml suspension for injection 3ml cartridges (Waymade Healthcare Plc) (product) |
|  | 14200411000001100 | Avandamet 2mg/500mg tablets (Sigma Pharmaceuticals Plc) (product) |
|  | 14200811000001100 | Avandia 4mg tablets (Sigma Pharmaceuticals Plc) (product) |
|  | 14201211000001100 | Avandia 8mg tablets (Sigma Pharmaceuticals Plc) (product) |
|  | 14247811000001100 | Humalog 100units/ml solution for injection 3ml cartridges (Sigma Pharmaceuticals Plc) (product) |
|  | 14248011000001100 | Humalog Mix25 100units/ml suspension for injection 3ml cartridges (Sigma Pharmaceuticals Plc) (product) |
|  | 14248811000001100 | Humalog Mix50 Pen 100units/ml suspension for injection 3ml pre-filled pens (Sigma Pharmaceuticals Plc) (product) |
|  | 14380511000001100 | NovoMix 30 FlexPen 100units/ml suspension for injection 3ml pre-filled pens (Sigma Pharmaceuticals Plc) (product) |
|  | 14386011000001100 | NovoMix 30 Penfill 100units/ml suspension for injection 3ml cartridges (Sigma Pharmaceuticals Plc) (product) |
|  | 14389911000001100 | NovoRapid FlexPen 100units/ml solution for injection 3ml pre-filled pens (Sigma Pharmaceuticals Plc) (product) |
|  | 14664711000001100 | NovoRapid Penfill 100units/ml solution for injection 3ml cartridges (Sigma Pharmaceuticals Plc) (product) |
|  | 14712111000001100 | Mixtard 30 Penfill 100units/ml suspension for injection 3ml cartridges (Sigma Pharmaceuticals Plc) (product) |
|  | 14723311000001100 | Prandin 1mg tablets (Sigma Pharmaceuticals Plc) (product) |
|  | 14723511000001100 | Prandin 2mg tablets (Sigma Pharmaceuticals Plc) (product) |
|  | 14753811000001100 | Prandin 0.5mg tablets (Sigma Pharmaceuticals Plc) (product) |
|  | 14760811000001100 | Lantus 100units/ml solution for injection 3ml OptiClik cartridges (Sigma Pharmaceuticals Plc) (product) |
|  | 14762411000001100 | Lantus 100units/ml solution for injection 10ml vials (Sigma Pharmaceuticals Plc) (product) |
|  | 14763111000001100 | Lantus 100units/ml solution for injection 3ml pre-filled OptiSet pens (Sigma Pharmaceuticals Plc) (product) |
|  | 14774311000001100 | Metformin 850mg tablets (LPC Medical (UK) Ltd) (product) |
|  | 147811000001105 | Tolbutamide 500mg tablets (Kent Pharmaceuticals Ltd) (product) |
|  | 15019811000001100 | Metformin 500mg modified-release tablets (Kent Pharmaceuticals Ltd) (product) |
|  | 15091011000001100 | Glimepiride 1mg tablets (Sigma Pharmaceuticals Plc) (product) |
|  | 15091411000001100 | Glimepiride 3mg tablets (Sigma Pharmaceuticals Plc) (product) |
|  | 15100211000001100 | Nazdol MR 30mg tablets (Teva UK Ltd) (product) |
|  | 15100511000001100 | Nazdol MR 30mg tablets (Generics (UK) Ltd) (product) |
|  | 152311000001101 | NovoNorm 500microgram tablets (Novo Nordisk Pharmaceuticals Ltd) (product) |
|  | 15374311000001100 | Glucophage 1000mg oral powder sachets (Merck Serono Ltd) (product) |
|  | 15411211000001100 | Metformin 1g oral powder sachets sugar free (product) |
|  | 15603111000001100 | Humulin R 500units/ml solution for injection 20ml vials (Imported (United States)) (product) |
|  | 15858611000001100 | Victoza 6mg/ml solution for injection 3ml pre-filled pens (Novo Nordisk Ltd) (product) |
|  | 15871011000001100 | Insulin human 100units/ml solution for injection 10ml vials (Special Order) (product) |
|  | 15973711000001100 | Metformin 850mg tablets (Zanza Laboratories Ltd) (product) |
|  | 16053711000001100 | Glimepiride 2mg tablets (Bristol Laboratories Ltd) (product) |
|  | 16054111000001100 | Glimepiride 4mg tablets (Bristol Laboratories Ltd) (product) |
|  | 16061411000001100 | Metformin 500mg tablets (Bristol Laboratories Ltd) (product) |
|  | 16125411000001100 | Glimepiride 2mg/5ml oral solution (Special Order) (product) |
|  | 16130611000001100 | Glimepiride 2mg/5ml oral solution (product) |
|  | 16130711000001100 | Glimepiride 2mg/5ml oral suspension (product) |
|  | 16133711000001100 | Actos 30mg tablets (Lexon (UK) Ltd) (product) |
|  | 16137211000001100 | Amaryl 1mg tablets (Lexon (UK) Ltd) (product) |
|  | 16137411000001100 | Amaryl 2mg tablets (Lexon (UK) Ltd) (product) |
|  | 16152911000001100 | Competact 15mg/850mg tablets (Lexon (UK) Ltd) (product) |
|  | 16187411000001100 | Glucophage SR 500mg tablets (Lexon (UK) Ltd) (product) |
|  | 16225011000001100 | Prandin 0.5mg tablets (Lexon (UK) Ltd) (product) |
|  | 16256311000001100 | Actos 30mg tablets (Mawdsley-Brooks & Company Ltd) (product) |
|  | 16259811000001100 | Amaryl 1mg tablets (Mawdsley-Brooks & Company Ltd) (product) |
|  | 16271611000001100 | Avandamet 2mg/1000mg tablets (Mawdsley-Brooks & Company Ltd) (product) |
|  | 16272811000001100 | Avandia 4mg tablets (Mawdsley-Brooks & Company Ltd) (product) |
|  | 16517211000001100 | Humalog Pen 100units/ml solution for injection 3ml pre-filled pens (Mawdsley-Brooks & Company Ltd) (product) |
|  | 16517411000001100 | Competact 15mg/850mg tablets (Mawdsley-Brooks & Company Ltd) (product) |
|  | 16517711000001100 | Humalog Mix25 100units/ml suspension for injection 3ml cartridges (Mawdsley-Brooks & Company Ltd) (product) |
|  | 16518911000001100 | Humalog Mix50 100units/ml suspension for injection 3ml cartridges (Mawdsley-Brooks & Company Ltd) (product) |
|  | 16519611000001100 | Humalog Mix50 Pen 100units/ml suspension for injection 3ml pre-filled pens (Mawdsley-Brooks & Company Ltd) (product) |
|  | 16520011000001100 | Humulin I 100units/ml suspension for injection 3ml cartridges (Mawdsley-Brooks & Company Ltd) (product) |
|  | 16520211000001100 | Humulin I 100units/ml suspension for injection 10ml vials (Mawdsley-Brooks & Company Ltd) (product) |
|  | 16521011000001100 | Humulin M3 100units/ml suspension for injection 3ml cartridges (Mawdsley-Brooks & Company Ltd) (product) |
|  | 16521311000001100 | Humulin S 100units/ml solution for injection 3ml cartridges (Mawdsley-Brooks & Company Ltd) (product) |
|  | 16523211000001100 | NovoMix 30 FlexPen 100units/ml suspension for injection 3ml pre-filled pens (Mawdsley-Brooks & Company Ltd) (product) |
|  | 16523811000001100 | NovoMix 30 Penfill 100units/ml suspension for injection 3ml cartridges (Mawdsley-Brooks & Company Ltd) (product) |
|  | 16530311000001100 | Humalog Mix25 100units/ml suspension for injection 10ml vials (Eli Lilly and Company Ltd) (product) |
|  | 16533211000001100 | Januvia 100mg tablets (Mawdsley-Brooks & Company Ltd) (product) |
|  | 16536211000001100 | Dacadis MR 30mg tablets (Generics (UK) Ltd) (product) |
|  | 16548811000001100 | Insulin lispro biphasic 25/75 100units/ml suspension for injection 10ml vials (product) |
|  | 16677511000001100 | Zicron 40mg tablets (Bristol Laboratories Ltd) (product) |
|  | 16701111000001100 | Metformin 500mg tablets (Pfizer Ltd) (product) |
|  | 17060511000001100 | Janumet 50mg/1000mg tablets (Merck Sharp & Dohme Ltd) (product) |
|  | 17071811000001100 | Metformin 1g / Sitagliptin 50mg tablets (product) |
|  | 17420011000001100 | Prandin 0.5mg tablets (Mawdsley-Brooks & Company Ltd) (product) |
|  | 17455811000001100 | Starlix 60mg tablets (Mawdsley-Brooks & Company Ltd) (product) |
|  | 17510111000001100 | Humalog 100units/ml solution for injection 10ml vials (Mawdsley-Brooks & Company Ltd) (product) |
|  | 17510311000001100 | Insulatard InnoLet 100units/ml suspension for injection 3ml pre-filled pens (Mawdsley-Brooks & Company Ltd) (product) |
|  | 17510511000001100 | Lantus 100units/ml solution for injection 3ml cartridges (Mawdsley-Brooks & Company Ltd) (product) |
|  | 17510711000001100 | Lantus 100units/ml solution for injection 3ml OptiClik cartridges (Mawdsley-Brooks & Company Ltd) (product) |
|  | 17536111000001100 | Competact 15mg/850mg tablets (Sigma Pharmaceuticals Plc) (product) |
|  | 17559311000001100 | Humulin I 100units/ml suspension for injection 10ml vials (Sigma Pharmaceuticals Plc) (product) |
|  | 17572311000001100 | Actos 15mg tablets (Necessity Supplies Ltd) (product) |
|  | 17572711000001100 | Actos 30mg tablets (Necessity Supplies Ltd) (product) |
|  | 17582511000001100 | Amaryl 3mg tablets (Necessity Supplies Ltd) (product) |
|  | 17595011000001100 | Avandia 8mg tablets (Necessity Supplies Ltd) (product) |
|  | 17602211000001100 | NovoRapid Penfill 100units/ml solution for injection 3ml cartridges (Necessity Supplies Ltd) (product) |
|  | 17608511000001100 | Humulin I KwikPen 100units/ml suspension for injection 3ml pre-filled pens (Eli Lilly and Company Ltd) (product) |
|  | 17622211000001100 | Prandin 1mg tablets (Necessity Supplies Ltd) (product) |
|  | 17622711000001100 | Prandin 2mg tablets (Necessity Supplies Ltd) (product) |
|  | 17895211000001100 | Actos 45mg tablets (Sigma Pharmaceuticals Plc) (product) |
|  | 17915111000001100 | Glibenclamide 5mg tablets (Phoenix Healthcare Distribution Ltd) (product) |
|  | 17916311000001100 | Glimepiride 1mg tablets (Phoenix Healthcare Distribution Ltd) (product) |
|  | 17916711000001100 | Glimepiride 2mg tablets (Phoenix Healthcare Distribution Ltd) (product) |
|  | 17917111000001100 | Glimepiride 3mg tablets (Phoenix Healthcare Distribution Ltd) (product) |
|  | 17917511000001100 | Glimepiride 4mg tablets (Phoenix Healthcare Distribution Ltd) (product) |
|  | 17940411000001100 | Metformin 850mg tablets (Phoenix Healthcare Distribution Ltd) (product) |
|  | 18048711000001100 | Humalog Mix25 KwikPen 100units/ml suspension for injection 3ml pre-filled pens (Sigma Pharmaceuticals Plc) (product) |
|  | 18048911000001100 | Humulin M3 100units/ml suspension for injection 10ml vials (Sigma Pharmaceuticals Plc) (product) |
|  | 18049911000001100 | Actos 15mg tablets (Lexon (UK) Ltd) (product) |
|  | 18061311000001100 | Januvia 100mg tablets (Lexon (UK) Ltd) (product) |
|  | 18083911000001100 | Insulin human 100units/ml solution for injection 3.15ml cartridges (product) |
|  | 18141911000001100 | Metabet SR 500mg tablets (Morningside Healthcare Ltd) (product) |
|  | 18251011000001100 | Diamicron 30mg MR tablets (Necessity Supplies Ltd) (product) |
|  | 18265411000001100 | Glucobay 50mg tablets (Necessity Supplies Ltd) (product) |
|  | 18266611000001100 | Humalog 100units/ml solution for injection 3ml cartridges (Necessity Supplies Ltd) (product) |
|  | 18266811000001100 | Humalog 100units/ml solution for injection 10ml vials (Necessity Supplies Ltd) (product) |
|  | 18286111000001100 | Humalog Mix50 100units/ml suspension for injection 3ml cartridges (Waymade Healthcare Plc) (product) |
|  | 18309611000001100 | Acarbose 50mg tablets (A A H Pharmaceuticals Ltd) (product) |
|  | 18462411000001100 | Glimepiride 1mg tablets (Accord Healthcare Ltd) (product) |
|  | 18490511000001100 | Lantus 100units/ml solution for injection 3ml cartridges (Necessity Supplies Ltd) (product) |
|  | 18490711000001100 | Lantus 100units/ml solution for injection 3ml OptiClik cartridges (Necessity Supplies Ltd) (product) |
|  | 18491611000001100 | Lantus 100units/ml solution for injection 3ml pre-filled SoloStar pens (Necessity Supplies Ltd) (product) |
|  | 18491811000001100 | Lantus 100units/ml solution for injection 10ml vials (Necessity Supplies Ltd) (product) |
|  | 18544011000001100 | Actos 45mg tablets (Doncaster Pharmaceuticals Ltd) (product) |
|  | 18553711000001100 | Humalog KwikPen 100units/ml solution for injection 3ml pre-filled pens (Waymade Healthcare Plc) (product) |
|  | 18553911000001100 | Humalog Mix25 KwikPen 100units/ml suspension for injection 3ml pre-filled pens (Waymade Healthcare Plc) (product) |
|  | 18554111000001100 | Humalog Mix50 KwikPen 100units/ml suspension for injection 3ml pre-filled pens (Waymade Healthcare Plc) (product) |
|  | 18570911000001100 | Competact 15mg/850mg tablets (Doncaster Pharmaceuticals Ltd) (product) |
|  | 18586911000001100 | Januvia 100mg tablets (Doncaster Pharmaceuticals Ltd) (product) |
|  | 18596311000001100 | Onglyza 2.5mg tablets (Bristol-Myers Squibb Pharmaceuticals Ltd) (product) |
|  | 18624411000001100 | Acarbose 100mg tablets (Phoenix Healthcare Distribution Ltd) (product) |
|  | 18630911000001100 | Lantus 100units/ml solution for injection 3ml pre-filled OptiSet pens (Mawdsley-Brooks & Company Ltd) (product) |
|  | 18885611000001100 | Metabet SR 1000mg tablets (Morningside Healthcare Ltd) (product) |
|  | 19179211000001100 | Repaglinide 500microgram tablets (Teva UK Ltd) (product) |
|  | 19179511000001100 | Repaglinide 1mg tablets (Teva UK Ltd) (product) |
|  | 19179811000001100 | Repaglinide 2mg tablets (Teva UK Ltd) (product) |
|  | 19276711000001100 | Repaglinide 2mg tablets (Aspire Pharma Ltd) (product) |
|  | 19299911000001100 | Repaglinide 500microgram tablets (Actavis UK Ltd) (product) |
|  | 19300811000001100 | Repaglinide 2mg tablets (Actavis UK Ltd) (product) |
|  | 19306711000001100 | Enyglid 0.5mg tablets (Consilient Health Ltd) (product) |
|  | 19308111000001100 | Glucient SR 500mg tablets (Consilient Health Ltd) (product) |
|  | 19360311000001100 | Repaglinide 500microgram tablets (Alliance Healthcare (Distribution) Ltd) (product) |
|  | 19360811000001100 | Repaglinide 1mg tablets (Alliance Healthcare (Distribution) Ltd) (product) |
|  | 19361511000001100 | Pioglitazone 30mg tablets (A A H Pharmaceuticals Ltd) (product) |
|  | 19469211000001100 | Pioglitazone 15mg tablets (Alliance Healthcare (Distribution) Ltd) (product) |
|  | 19469411000001100 | Pioglitazone 30mg tablets (Alliance Healthcare (Distribution) Ltd) (product) |
|  | 19476211000001100 | Pioglitazone 15mg tablets (Consilient Health Ltd) (product) |
|  | 19489311000001100 | Acarbose 50mg tablets (Alliance Healthcare (Distribution) Ltd) (product) |
|  | 19492811000001100 | Trajenta 5mg tablets (Boehringer Ingelheim Ltd) (product) |
|  | 19525211000001100 | Linagliptin 5mg tablets (product) |
|  | 19573011000001100 | Metformin 1g modified-release tablets (A A H Pharmaceuticals Ltd) (product) |
|  | 19592911000001100 | Pioglitazone 15mg tablets (Dr Reddy's Laboratories (UK) Ltd) (product) |
|  | 19593111000001100 | Pioglitazone 30mg tablets (Dr Reddy's Laboratories (UK) Ltd) (product) |
|  | 19612811000001100 | Galvus 50mg tablets (Doncaster Pharmaceuticals Ltd) (product) |
|  | 19613111000001100 | Humalog KwikPen 100units/ml solution for injection 3ml pre-filled pens (DE Pharmaceuticals) (product) |
|  | 19613311000001100 | Humalog Mix25 KwikPen 100units/ml suspension for injection 3ml pre-filled pens (DE Pharmaceuticals) (product) |
|  | 19613711000001100 | Humalog Mix25 100units/ml suspension for injection 3ml cartridges (Doncaster Pharmaceuticals Ltd) (product) |
|  | 19613911000001100 | Humalog Mix50 100units/ml suspension for injection 3ml cartridges (Doncaster Pharmaceuticals Ltd) (product) |
|  | 19614311000001100 | Humulin I 100units/ml suspension for injection 10ml vials (Doncaster Pharmaceuticals Ltd) (product) |
|  | 19700011000001100 | Acarbose 100mg tablets (Doncaster Pharmaceuticals Ltd) (product) |
|  | 19700511000001100 | Acarbose 50mg tablets (Doncaster Pharmaceuticals Ltd) (product) |
|  | 19873911000001100 | Eucreas 50mg/1000mg tablets (Doncaster Pharmaceuticals Ltd) (product) |
|  | 19958011000001100 | Glizofar 30mg tablets (Teva UK Ltd) (product) |
|  | 19958211000001100 | Glizofar 45mg tablets (Teva UK Ltd) (product) |
|  | 20115111000001100 | Januvia 25mg Tablets (Merck Sharp & Dohme Ltd) (product) |
|  | 20133211000001100 | Januvia 100mg tablets (Waymade Healthcare Plc) (product) |
|  | 20163311000001100 | NovoMix 30 Penfill 100units/ml suspension for injection 3ml cartridges (Doncaster Pharmaceuticals Ltd) (product) |
|  | 20166811000001100 | Glimepiride 2mg tablets (Accord Healthcare Ltd) (product) |
|  | 202611000001102 | Metformin 500mg tablets (A A H Pharmaceuticals Ltd) (product) |
|  | 20357411000001100 | Pioglitazone 15mg tablets (Actavis UK Ltd) (product) |
|  | 20357811000001100 | Pioglitazone 45mg tablets (Actavis UK Ltd) (product) |
|  | 20552511000001100 | Diagemet XL 500mg tablets (Genus Pharmaceuticals Ltd) (product) |
|  | 20566511000001100 | Repaglinide 1mg tablets (Creo Pharma Ltd) (product) |
|  | 20777611000001100 | Repaglinide 1mg tablets (Accord Healthcare Ltd) (product) |
|  | 20916911000001100 | Pioglitazone 15mg tablets (Zentiva) (product) |
|  | 20917111000001100 | Pioglitazone 30mg tablets (Zentiva) (product) |
|  | 20917311000001100 | Pioglitazone 45mg tablets (Zentiva) (product) |
|  | 20946611000001100 | Gliclazide 30mg modified-release tablets (Actavis UK Ltd) (product) |
|  | 21022811000001100 | Janumet 50mg/1000mg tablets (Waymade Healthcare Plc) (product) |
|  | 21027911000001100 | Acarbose 50mg tablets (Arrow Generics Ltd) (product) |
|  | 21112511000001100 | Acarbose 100mg tablets (Actavis UK Ltd) (product) |
|  | 21203911000001100 | Lantus 100units/ml solution for injection 3ml pre-filled SoloStar pens (Waymade Healthcare Plc) (product) |
|  | 21324011000001100 | Glimepiride 6mg/5ml oral suspension (Special Order) (product) |
|  | 21609511000001100 | Forxiga 5mg tablets (Bristol-Myers Squibb Pharmaceuticals Ltd) (product) |
|  | 21609811000001100 | Forxiga 10mg tablets (Bristol-Myers Squibb Pharmaceuticals Ltd) (product) |
|  | 21734711000001100 | Acarbose 50mg tablets (Waymade Healthcare Plc) (product) |
|  | 21792811000001100 | Metformin 500mg modified-release tablets (Waymade Healthcare Plc) (product) |
|  | 21880311000001100 | Repaglinide 1mg tablets (Waymade Healthcare Plc) (product) |
|  | 21884111000001100 | Tolbutamide 500mg tablets (Waymade Healthcare Plc) (product) |
|  | 21928511000001100 | Tresiba Penfill 100units/ml solution for injection 3ml cartridges (Novo Nordisk Ltd) (product) |
|  | 21939511000001100 | Insulin degludec 100units/ml solution for injection 3ml cartridges (product) |
|  | 21941011000001100 | Lyxumia 20micrograms/0.2ml solution for injection 3ml pre-filled pens (Sanofi) (product) |
|  | 21953711000001100 | Lyxumia 10micrograms/0.2ml solution for injection 3ml pre-filled pen and Lyxumia 20micrograms/0.2ml solution for injection 3ml pre-filled pen (Sanofi) (product) |
|  | 21994611000001100 | Lixisenatide 10micrograms/0.2ml solution for injection 3ml pre-filled disposable devices (product) |
|  | 21994711000001100 | Lixisenatide 10micrograms/0.2ml solution for injection 3ml pre-filled disposable devices and Lixisenatide 20micrograms/0.2ml solution for injection 3ml pre-filled disposable devices (product) |
|  | 22026311000001100 | Glibenclamide 5mg tablets (Waymade Healthcare Plc) (product) |
|  | 22027111000001100 | Glimepiride 1mg tablets (Waymade Healthcare Plc) (product) |
|  | 22027411000001100 | Glimepiride 2mg tablets (Waymade Healthcare Plc) (product) |
|  | 22028211000001100 | Glimepiride 4mg tablets (Waymade Healthcare Plc) (product) |
|  | 22028411000001100 | Glipizide 5mg tablets (Waymade Healthcare Plc) (product) |
|  | 22105211000001100 | Metformin 500mg/5ml oral solution sugar free (Zentiva) (product) |
|  | 22225011000001100 | Laaglyda MR 60 mg tablets (Consilient Health Ltd) (product) |
|  | 22226111000001100 | Gliclazide 60mg modified-release tablets (product) |
|  | 22349811000001100 | Metformin 500mg tablets (Aurobindo Pharma Ltd) (product) |
|  | 22393411000001100 | Metformin 500mg/5ml oral solution sugar free (Waymade Healthcare Plc) (product) |
|  | 22879311000001100 | Januvia 50mg tablets (Waymade Healthcare Plc) (product) |
|  | 22949011000001100 | Pioglitazone 15mg tablets (Ranbaxy (UK) Ltd) (product) |
|  | 22949311000001100 | Pioglitazone 30mg tablets (Ranbaxy (UK) Ltd) (product) |
|  | 22949511000001100 | Pioglitazone 45mg tablets (Ranbaxy (UK) Ltd) (product) |
|  | 233411000001102 | Euglucon 2.5mg tablets (Aventis Pharma) (product) |
|  | 23372211000001100 | Glidipion 45mg tablets (Actavis UK Ltd) (product) |
|  | 234011000001108 | Daonil 5mg tablets (Aventis Pharma) (product) |
|  | 23487611000001100 | Pioglitazone 30mg tablets (Accord Healthcare Ltd) (product) |
|  | 23632611000001100 | Vipdomet 12.5mg/1000mg tablets (Takeda UK Ltd) (product) |
|  | 23634111000001100 | Vipidia 6.25mg tablets (Takeda UK Ltd) (product) |
|  | 23636011000001100 | Vipidia 12.5mg tablets (Takeda UK Ltd) (product) |
|  | 23636311000001100 | Vipidia 25mg tablets (Takeda UK Ltd) (product) |
|  | 23637211000001100 | Alogliptin 12.5mg / Metformin 1g tablets (product) |
|  | 23637411000001100 | Alogliptin 25mg tablets (product) |
|  | 23637511000001100 | Alogliptin 6.25mg tablets (product) |
|  | 23677111000001100 | Januvia 25mg tablets (Waymade Healthcare Plc) (product) |
|  | 23920511000001100 | Gliclazide 30mg modified-release tablets (Phoenix Healthcare Distribution Ltd) (product) |
|  | 23943411000001100 | Glibenclamide 5mg tablets (Doncaster Pharmaceuticals Ltd) (product) |
|  | 24054611000001100 | Dapagliflozin 5mg / Metformin 1g tablets (product) |
|  | 24104511000001100 | Canagliflozin 300mg tablets (product) |
|  | 24106311000001100 | Glimepiride 1mg tablets (DE Pharmaceuticals) (product) |
|  | 24106511000001100 | Glimepiride 2mg tablets (DE Pharmaceuticals) (product) |
|  | 24130611000001100 | Acarbose 50mg tablets (Morningside Healthcare Ltd) (product) |
|  | 24130811000001100 | Acarbose 100mg tablets (Morningside Healthcare Ltd) (product) |
|  | 24135111000001100 | Pioglitazone 45mg tablets (Morningside Healthcare Ltd) (product) |
|  | 24380011000001100 | Metformin 500mg tablets (DE Pharmaceuticals) (product) |
|  | 24380511000001100 | Metformin 850mg tablets (DE Pharmaceuticals) (product) |
|  | 24554311000001100 | Metformin 500mg modified-release tablets (Actavis UK Ltd) (product) |
|  | 24568211000001100 | Sukkarto SR 1000mg tablets (Morningside Healthcare Ltd) (product) |
|  | 25290611000001100 | Empagliflozin 25mg tablets (product) |
|  | 257011000001104 | Metformin 500mg tablets (Generics (UK) Ltd) (product) |
|  | 259611000001101 | Amaryl 4mg tablets (Aventis Pharma) (product) |
|  | 26209611000001100 | NovoRapid PumpCart 100units/ml solution for injection 1.6ml cartridges (Novo Nordisk Ltd) (product) |
|  | 26655811000001100 | Insulin aspart 100units/ml solution for injection 1.6ml cartridges (product) |
|  | 270611000001105 | Avandia 8mg tablets (GlaxoSmithKline) (product) |
|  | 27990411000001100 | Forxiga 10mg tablets (Waymade Healthcare Plc) (product) |
|  | 27990611000001100 | Galvus 50mg tablets (Waymade Healthcare Plc) (product) |
|  | 28044911000001100 | Insulin human 500units/ml solution for injection 20ml vials (Special Order) (product) |
|  | 28054311000001100 | Xultophy 100units/ml / 3.6mg/ml solution for injection 3ml pre-filled pens (Novo Nordisk Ltd) (product) |
|  | 28277811000001100 | Victoza 6mg/ml solution for injection 3ml pre-filled pens (Waymade Healthcare Plc) (product) |
|  | 28415811000001100 | Metformin 500mg/5ml oral solution sugar free (Focus Pharmaceuticals Ltd) (product) |
|  | 28420711000001100 | Vamju 30mg modified-release tablets (AMCo) (product) |
|  | 28426011000001100 | Bydureon 2mg powder and solvent for prolonged-release suspension for injection pre-filled pens (AstraZeneca UK Ltd) (product) |
|  | 28461011000001100 | Trulicity 0.75mg/0.5ml solution for injection pre-filled pens (Eli Lilly and Company Ltd) (product) |
|  | 28775211000001100 | Canagliflozin 100mg tablets (Colorama Pharmaceuticals Ltd) (product) |
|  | 28776011000001100 | Liraglutide 6mg/ml solution for injection 3ml pre-filled disposable devices (Colorama Pharmaceuticals Ltd) (product) |
|  | 28785111000001100 | Metformin 1g oral powder sachets sugar free (J M McGill Ltd) (product) |
|  | 28789611000001100 | Dulaglutide 0.75mg/0.5ml solution for injection pre-filled disposable devices (product) |
|  | 28989711000001100 | Insulin lispro 200units/ml solution for injection 3ml pre-filled disposable devices (product) |
|  | 29699911000001100 | Canagliflozin 100mg tablets (J M McGill Ltd) (product) |
|  | 29742811000001100 | Diabiom 15mg tablets (Tillomed Laboratories Ltd) (product) |
|  | 29754211000001100 | Acarbose 100mg tablets (Sigma Pharmaceuticals Plc) (product) |
|  | 29854711000001100 | Liraglutide 6mg/ml solution for injection 3ml pre-filled disposable devices (Niche Pharma Ltd) (product) |
|  | 29869311000001100 | Glibenclamide 2.5mg tablets (Sigma Pharmaceuticals Plc) (product) |
|  | 29869511000001100 | Glibenclamide 5mg tablets (Sigma Pharmaceuticals Plc) (product) |
|  | 29906411000001100 | Metformin 850mg tablets (Sigma Pharmaceuticals Plc) (product) |
|  | 29918411000001100 | Metformin 500mg/5ml oral solution sugar free (Sigma Pharmaceuticals Plc) (product) |
|  | 29960011000001100 | Apidra 100units/ml solution for injection 3ml pre-filled SoloStar pens (Lexon (UK) Ltd) (product) |
|  | 29985811000001100 | Pioglitazone 15mg tablets (Sigma Pharmaceuticals Plc) (product) |
|  | 29986011000001100 | Pioglitazone 30mg tablets (Sigma Pharmaceuticals Plc) (product) |
|  | 30012111000001100 | Glucient SR 750mg tablets (Consilient Health Ltd) (product) |
|  | 30012311000001100 | Glucient SR 1000mg tablets (Consilient Health Ltd) (product) |
|  | 30077211000001100 | Pioglitazone 15mg tablets (DE Pharmaceuticals) (product) |
|  | 30100711000001100 | Tolbutamide 500mg tablets (DE Pharmaceuticals) (product) |
|  | 30112111000001100 | Pioglitazone 30mg tablets (DE Pharmaceuticals) (product) |
|  | 30134511000001100 | Glibenclamide 2.5mg tablets (Mawdsley-Brooks & Company Ltd) (product) |
|  | 30134711000001100 | Glibenclamide 5mg tablets (Mawdsley-Brooks & Company Ltd) (product) |
|  | 30137811000001100 | Glimepiride 2mg tablets (Mawdsley-Brooks & Company Ltd) (product) |
|  | 30138311000001100 | Glimepiride 3mg tablets (Mawdsley-Brooks & Company Ltd) (product) |
|  | 30138511000001100 | Glimepiride 4mg tablets (Mawdsley-Brooks & Company Ltd) (product) |
|  | 30173411000001100 | Synjardy 5mg/850mg tablets (Boehringer Ingelheim Ltd) (product) |
|  | 30208411000001100 | Eucreas 50mg/850mg tablets (Waymade Healthcare Plc) (product) |
|  | 30214811000001100 | Vipidia 25mg tablets (Waymade Healthcare Plc) (product) |
|  | 30268411000001100 | Apidra 100units/ml solution for injection 3ml pre-filled SoloStar pens (Waymade Healthcare Plc) (product) |
|  | 30318111000001100 | Empagliflozin 12.5mg / Metformin 1g tablets (product) |
|  | 30829111000001100 | Repaglinide 500microgram tablets (DE Pharmaceuticals) (product) |
|  | 30857011000001100 | Pioglitazone 15mg tablets (Mawdsley-Brooks & Company Ltd) (product) |
|  | 30875411000001100 | Repaglinide 500microgram tablets (Mawdsley-Brooks & Company Ltd) (product) |
|  | 30875711000001100 | Repaglinide 1mg tablets (Mawdsley-Brooks & Company Ltd) (product) |
|  | 30876011000001100 | Repaglinide 2mg tablets (Mawdsley-Brooks & Company Ltd) (product) |
|  | 30928411000001100 | Liraglutide 6mg/ml solution for injection 3ml pre-filled disposable devices (DE Pharmaceuticals) (product) |
|  | 30983711000001100 | Bilxona 60mg modified-release tablets (Actavis UK Ltd) (product) |
|  | 31351911000001100 | Alogliptin 12.5mg tablets (J M McGill Ltd) (product) |
|  | 31352311000001100 | Alogliptin 6.25mg tablets (J M McGill Ltd) (product) |
|  | 322511000001103 | Euglucon 5mg tablets (Aventis Pharma) (product) |
|  | 32413911000001100 | Gliclazide 80mg tablets (Genesis Pharmaceuticals Ltd) (product) |
|  | 32431711000001100 | Alogliptin 12.5mg / Metformin 1g tablets (Colorama Pharmaceuticals Ltd) (product) |
|  | 32431911000001100 | Alogliptin 12.5mg tablets (Colorama Pharmaceuticals Ltd) (product) |
|  | 32492811000001100 | Pioglitazone 15mg tablets (Brown & Burk UK Ltd) (product) |
|  | 32493011000001100 | Pioglitazone 30mg tablets (Brown & Burk UK Ltd) (product) |
|  | 32493211000001100 | Pioglitazone 45mg tablets (Brown & Burk UK Ltd) (product) |
|  | 32493811000001100 | Glimepiride 3mg tablets (Brown & Burk UK Ltd) (product) |
|  | 325218001 | Product containing precisely glibenclamide 2.5 milligram/1 each conventional release oral tablet (clinical drug) |
|  | 325242002 | Product containing precisely gliclazide 80 milligram/1 each conventional release oral tablet (clinical drug) |
|  | 325243007 | Product containing precisely glipizide 5 milligram/1 each conventional release oral tablet (clinical drug) |
|  | 325248003 | Product containing precisely glipizide 2.5 milligram/1 each conventional release oral tablet (clinical drug) |
|  | 325262005 | Product containing precisely glimepiride 3 milligram/1 each conventional release oral tablet (clinical drug) |
|  | 325263000 | Product containing precisely glimepiride 4 milligram/1 each conventional release oral tablet (clinical drug) |
|  | 325279004 | Product containing precisely metformin hydrochloride 850 milligram/1 each conventional release oral tablet (clinical drug) |
|  | 3255911000001100 | Humulin Isophane 100units/ml suspension for injection 10ml vials (Eli Lilly & Co Ltd) (product) |
|  | 3258411000001100 | Insuman Basal 100units/ml suspension for injection 5ml vials (Aventis Pharma) (product) |
|  | 326037007 | Product containing precisely acarbose 50 milligram/1 each conventional release oral tablet (clinical drug) |
|  | 326048000 | Product containing precisely repaglinide 1 milligram/1 each conventional release oral tablet (clinical drug) |
|  | 326049008 | Product containing precisely repaglinide 2 milligram/1 each conventional release oral tablet (clinical drug) |
|  | 326056002 | Product containing precisely rosiglitazone (as rosiglitazone maleate) 8 milligram/1 each conventional release oral tablet (clinical drug) |
|  | 326057006 | Product containing precisely rosiglitazone (as rosiglitazone maleate) 4 milligram/1 each conventional release oral tablet (clinical drug) |
|  | 3260611000001100 | Insuman Basal 100units/ml suspension for injection 3ml pre-filled OptiSet pens (Sanofi) (product) |
|  | 326062007 | Product containing precisely pioglitazone (as pioglitazone hydrochloride) 15 milligram/1 each conventional release oral tablet (clinical drug) |
|  | 3262011000001100 | Insulatard FlexPen 100units/ml suspension for injection (Novo Nordisk Pharmaceuticals Ltd) (product) |
|  | 3262511000001100 | Insulatard NovoLet 100units/ml suspension for injection (Novo Nordisk Pharmaceuticals Ltd) (product) |
|  | 32625911000001100 | Sitagliptin 50mg/5ml oral solution (Special Order) (product) |
|  | 3263711000001100 | Insulatard Penfill 100units/ml suspension for injection 3ml cartridges (Novo Nordisk Pharmaceuticals Ltd) (product) |
|  | 3264111000001100 | Insuman Comb 25 100units/ml suspension for injection 3ml cartridges (Aventis Pharma) (product) |
|  | 3264711000001100 | Mixtard 10 Penfill 100units/ml suspension for injection 3ml cartridges (Novo Nordisk Pharmaceuticals Ltd) (product) |
|  | 3265011000001100 | Insuman Basal 100units/ml suspension for injection 3ml cartridges (Aventis Pharma) (product) |
|  | 3266111000001100 | Mixtard 20 NovoLet 100units/ml suspension for injection (Novo Nordisk Pharmaceuticals Ltd) (product) |
|  | 3266811000001100 | Hypurin Porcine 30/70 Mix 100units/ml suspension for injection 10ml vials (C P Pharmaceuticals Ltd) (product) |
|  | 3267811000001100 | Human Mixtard 20 Penfill 100units/ml suspension for injection 1.5ml cartridges (Novo Nordisk Pharmaceuticals Ltd) (product) |
|  | 3267911000001100 | Pork Mixtard 30 100units/ml suspension for injection 10ml vials (Novo Nordisk Pharmaceuticals Ltd) (product) |
|  | 3270511000001100 | Mixtard 30 NovoLet 100units/ml suspension for injection (Novo Nordisk Pharmaceuticals Ltd) (product) |
|  | 3271011000001100 | Humulin M5 100units/ml suspension for injection 10ml vials (Eli Lilly & Co Ltd) (product) |
|  | 3272011000001100 | Mixtard 40 NovoLet 100units/ml suspension for injection (Novo Nordisk Pharmaceuticals Ltd) (product) |
|  | 3272811000001100 | Mixtard 50 Penfill 100units/ml suspension for injection 3ml cartridges (Novo Nordisk Pharmaceuticals Ltd) (product) |
|  | 3273611000001100 | Humulin M3 100units/ml suspension for injection 3ml cartridges (Eli Lilly & Co Ltd) (product) |
|  | 3274511000001100 | Insuman Comb 50 100units/ml suspension for injection 5ml vials (Aventis Pharma) (product) |
|  | 3275011000001100 | Mixtard 30 ge 100units/ml suspension for injection 10ml vials (Novo Nordisk Pharmaceuticals Ltd) (product) |
|  | 3276911000001100 | Humalog Mix50 Pen 100units/ml suspension for injection 3ml pre-filled pens (Eli Lilly and Company Ltd) (product) |
|  | 3277211000001100 | NovoMix 30 FlexPen 100units/ml suspension for injection 3ml pre-filled pens (Novo Nordisk Ltd) (product) |
|  | 3277711000001100 | Hypurin Porcine 30/70 Mix 100units/ml suspension for injection 1.5ml cartridges (C P Pharmaceuticals Ltd) (product) |
|  | 3278311000001100 | Mixtard 50 NovoLet 100units/ml suspension for injection (Novo Nordisk Pharmaceuticals Ltd) (product) |
|  | 3278511000001100 | Humalog Pen 100units/ml solution for injection 3ml pre-filled pens (Eli Lilly and Company Ltd) (product) |
|  | 3280111000001100 | Humalog 100units/ml solution for injection 10ml vials (Eli Lilly & Co Ltd) (product) |
|  | 3281211000001100 | Mixtard 40 Penfill 100units/ml suspension for injection 3ml cartridges (Novo Nordisk Pharmaceuticals Ltd) (product) |
|  | 3281611000001100 | NovoRapid Novolet 100units/ml solution for injection (Novo Nordisk Pharmaceuticals Ltd) (product) |
|  | 3282711000001100 | Humalog 100units/ml solution for injection 1.5ml cartridges (Eli Lilly & Co Ltd) (product) |
|  | 3283211000001100 | Lantus 100units/ml solution for injection 3ml pre-filled OptiSet pens (Sanofi) (product) |
|  | 3283411000001100 | Hypurin Porcine Neutral 100units/ml suspension for injection 1.5ml cartridges (C P Pharmaceuticals Ltd) (product) |
|  | 3284911000001100 | Humulin Lente 100units/ml suspension for injection 10ml vials (Eli Lilly & Co Ltd) (product) |
|  | 3285011000001100 | Hypurin Porcine Neutral 100units/ml solution for injection 10ml vials (C P Pharmaceuticals Ltd) (product) |
|  | 32857511000001100 | Sitagliptin 50mg/5ml oral suspension (Special Order) (product) |
|  | 32879911000001100 | Sitagliptin 50mg/5ml oral suspension (product) |
|  | 3288511000001100 | Actrapid NovoLet 100units/ml solution for injection (Novo Nordisk Pharmaceuticals Ltd) (product) |
|  | 3290411000001100 | HumaJect S Pen 100units/ml solution for injection (Eli Lilly & Co Ltd) (product) |
|  | 3294911000001100 | Insuman Rapid 100units/ml solution for injection 5ml vials (Aventis Pharma) (product) |
|  | 3309511000001100 | Human Actrapid Penfill 100units/ml solution for injection 1.5ml cartridges (Novo Nordisk Pharmaceuticals Ltd) (product) |
|  | 3310711000001100 | Actrapid Penfill 100units/ml solution for injection 3ml cartridges (Novo Nordisk Pharmaceuticals Ltd) (product) |
|  | 3312611000001100 | Humulin S 100units/ml solution for injection 10ml vials (Eli Lilly & Co Ltd) (product) |
|  | 33547711000001100 | Metformin 850mg/5ml oral solution sugar free (Colonis Pharma Ltd) (product) |
|  | 33548911000001100 | Metformin 500mg/5ml oral solution sugar free (Colonis Pharma Ltd) (product) |
|  | 33549511000001100 | Acarbose 50mg tablets (Mylan Ltd) (product) |
|  | 33549711000001100 | Acarbose 100mg tablets (Mylan Ltd) (product) |
|  | 33550811000001100 | Metformin 1g/5ml oral solution sugar free (product) |
|  | 33599111000001100 | Pioglitazone 15mg tablets (Mylan Ltd) (product) |
|  | 33619611000001100 | Repaglinide 2mg tablets (Mylan Ltd) (product) |
|  | 3468611000001100 | Insulin aspart 100units/ml solution for injection 3ml cartridges (product) |
|  | 3469811000001100 | Insulin isophane biphasic human 15/85 100units/ml suspension for injection 5ml vials (product) |
|  | 3470111000001100 | Insulin isophane biphasic human 20/80 100units/ml suspension for injection 3ml pre-filled disposable devices (product) |
|  | 3470211000001100 | Insulin isophane biphasic human 25/75 100units/ml suspension for injection 3ml cartridges (product) |
|  | 3470711000001100 | Insulin isophane biphasic human 30/70 100units/ml suspension for injection 10ml vials (product) |
|  | 3471011000001100 | Insulin isophane biphasic human 30/70 100units/ml suspension for injection 3ml pre-filled disposable devices (product) |
|  | 3471311000001100 | Insulin isophane biphasic human 40/60 100units/ml suspension for injection 3ml pre-filled disposable devices (product) |
|  | 3471711000001100 | Insulin isophane biphasic human 50/50 100units/ml suspension for injection 3ml pre-filled disposable devices (product) |
|  | 3472011000001100 | Insulin isophane biphasic porcine 30/70 100units/ml suspension for injection 10ml vials (product) |
|  | 3472411000001100 | Insulin isophane human 100units/ml suspension for injection 1.5ml cartridges (product) |
|  | 3472511000001100 | Insulin isophane human 100units/ml suspension for injection 10ml vials (product) |
|  | 3472811000001100 | Insulin isophane human 100units/ml suspension for injection 5ml vials (product) |
|  | 3473011000001100 | Insulin isophane porcine 100units/ml suspension for injection 10ml vials (product) |
|  | 3474611000001100 | Insulin soluble porcine 100units/ml suspension for injection 1.5ml cartridges (product) |
|  | 363211000001102 | Glucophage 500mg tablets (Merck Pharmaceuticals) (product) |
|  | 365111000001109 | Glucophage 850mg tablets (Merck Pharmaceuticals) (product) |
|  | 3651211000001100 | Starlix 120mg tablets (Novartis Pharmaceuticals UK Ltd) (product) |
|  | 3763311000001100 | Glurenorm 30mg tablets (Sanofi-Synthelabo Ltd) (product) |
|  | 378911000001109 | Gliclazide 80mg tablets (A A H Pharmaceuticals Ltd) (product) |
|  | 386011000001102 | Glibenclamide 2.5mg tablets (Generics (UK) Ltd) (product) |
|  | 3883511000001100 | Starlix 180mg tablets (Novartis Pharmaceuticals UK Ltd) (product) |
|  | 393311000001107 | Tolbutamide 500mg tablets (Unichem Plc) (product) |
|  | 4029411000001100 | Hypurin Porcine 30/70 Mix 100units/ml suspension for injection 3ml cartridges (C P Pharmaceuticals Ltd) (product) |
|  | 4033311000001100 | Insulin soluble bovine 100units/ml solution for injection 3ml cartridges (product) |
|  | 4034911000001100 | Hypurin Porcine Neutral 100units/ml suspension for injection 3ml cartridges (C P Pharmaceuticals Ltd) (product) |
|  | 4053711000001100 | Insulin soluble porcine 100units/ml suspension for injection 3ml cartridges (product) |
|  | 409121008 | Product containing precisely metformin hydrochloride 500 milligram and rosiglitazone (as rosiglitazone maleate) 1 milligram/1 each conventional release oral tablet (clinical drug) |
|  | 409125004 | Product containing precisely metformin hydrochloride 1 gram and rosiglitazone (as rosiglitazone maleate) 4 milligram/1 each conventional release oral tablet (clinical drug) |
|  | 409361001 | Metformin hydrochloride 750mg m/r tablet (product) |
|  | 411533003 | Metformin hydrochloride 1g m/r tablet (product) |
|  | 419873003 | Product containing precisely metformin hydrochloride 850 milligram and pioglitazone (as pioglitazone hydrochloride) 15 milligram/1 each conventional release oral tablet (clinical drug) |
|  | 424345005 | Product containing precisely sitagliptin (as sitagliptin phosphate) 100 milligram/1 each conventional release oral tablet (clinical drug) |
|  | 443067000 | Product containing precisely saxagliptin 2.5 milligram/1 each conventional release oral tablet (clinical drug) |
|  | 443713000 | Product containing precisely saxagliptin 5 milligram/1 each conventional release oral tablet (clinical drug) |
|  | 4470711000001100 | Tolbutamide 500mg tablets (Generics (UK) Ltd) (product) |
|  | 464111000001100 | Glipizide 5mg tablets (Generics (UK) Ltd) (product) |
|  | 49111000001103 | Diaglyk 80mg tablets (Ashbourne Pharmaceuticals Ltd) (product) |
|  | 5268311000001100 | Humalog 100units/ml solution for injection 3ml cartridges (PI) (Waymade Ltd) (product) |
|  | 5268611000001100 | Humalog Mix25 Pen 100units/ml suspension for injection 3ml pre-filled pens (Waymade Healthcare Plc) (product) |
|  | 5269111000001100 | Humalog Mix50 Pen 100units/ml suspension for injection 3ml pre-filled pens (Waymade Healthcare Plc) (product) |
|  | 5270511000001100 | NovoRapid Penfill 100units/ml solution for injection 3ml cartridges (PI) (Waymade Ltd) (product) |
|  | 5295811000001100 | Humalog 100units/ml solution for injection 1.5ml cartridges (PI) (Dowelhurst Ltd) (product) |
|  | 5296011000001100 | Humalog 100units/ml solution for injection 10ml vials (PI) (Dowelhurst Ltd) (product) |
|  | 5302911000001100 | Avandamet 1mg/500mg tablets (GlaxoSmithKline) (product) |
|  | 5303611000001100 | Avandamet 2mg/500mg tablets (GlaxoSmithKline) (product) |
|  | 5330011000001100 | Actos 30mg tablets (PI) (Waymade Ltd) (product) |
|  | 5331011000001100 | Amaryl 4mg tablets (PI) (Waymade Ltd) (product) |
|  | 533211000001108 | Gliclazide 80mg tablets (Generics (UK) Ltd) (product) |
|  | 5336711000001100 | Avandia 4mg tablets (PI) (Waymade Ltd) (product) |
|  | 5355711000001100 | Daonil 5mg tablets (PI) (Waymade Ltd) (product) |
|  | 5369511000001100 | Glibenese 5mg tablets (PI) (Waymade Ltd) (product) |
|  | 5371611000001100 | Glucobay 50 tablets (PI) (Waymade Ltd) (product) |
|  | 5395711000001100 | NovoNorm 500microgram tablets (PI) (Waymade Ltd) (product) |
|  | 5396211000001100 | NovoNorm 2mg tablets (PI) (Waymade Ltd) (product) |
|  | 5415211000001100 | Starlix 180mg tablets (PI) (Waymade Ltd) (product) |
|  | 543011000001102 | Glyformin 500mg tablets (Dr Reddy's Labs) (product) |
|  | 5441111000001100 | Acarbose 100mg tablets (PI) (Dowelhurst Ltd) (product) |
|  | 5448611000001100 | Glimepiride 1mg tablets (PI) (Dowelhurst Ltd) (product) |
|  | 5481211000001100 | Minodiab 5mg tablets (PI) (Waymade Ltd) (product) |
|  | 5520811000001100 | Amaryl 3mg tablets (PI) (Dowelhurst Ltd) (product) |
|  | 622811000001108 | Gliclazide 80mg tablets (C P Pharmaceuticals Ltd) (product) |
|  | 639511000001103 | Tolbutamide 500mg tablets (Approved Prescription Services) (product) |
|  | 648011000001102 | Glibenclamide 2.5mg tablets (A A H Pharmaceuticals Ltd) (product) |
|  | 652411000001107 | Minodiab 5mg tablets (Pfizer Ltd) (product) |
|  | 656211000001109 | Glibenclamide 2.5mg tablets (Unichem Plc) (product) |
|  | 690811000001103 | Metformin 500mg tablets (Kent Pharmaceuticals Ltd) (product) |
|  | 695711000001101 | Glibenclamide 5mg tablets (Alpharma Limited) (product) |
|  | 697811000001104 | Glipizide 5mg tablets (IVAX Pharmaceuticals UK Ltd) (product) |
|  | 698511000001103 | Glipizide 5mg tablets (Sandoz Ltd) (product) |
|  | 7016011000001100 | Metformin 500mg tablets (IVAX Pharmaceuticals UK Ltd) (product) |
|  | 703682001 | Product containing precisely canagliflozin 100 milligram/1 each conventional release oral tablet (clinical drug) |
|  | 731211000001105 | Glipizide 5mg tablets (A A H Pharmaceuticals Ltd) (product) |
|  | 745211000001107 | Glibenclamide 5mg tablets (A A H Pharmaceuticals Ltd) (product) |
|  | 745911000001103 | Diamicron 80mg tablets (Servier Laboratories Limited) (product) |
|  | 7461611000001100 | Metformin 500mg tablets (Ranbaxy (UK) Ltd) (product) |
|  | 756411000001104 | Glipizide 5mg tablets (Alpharma Limited) (product) |
|  | 7589911000001100 | Levemir Penfill 100units/ml solution for injection 3ml cartridges (Novo Nordisk Pharmaceuticals Ltd) (product) |
|  | 7594211000001100 | Insulin detemir 100units/ml solution for injection 3ml pre-filled disposable devices (product) |
|  | 7597611000001100 | Insulin detemir 100units/ml solution for injection 3ml cartridges (product) |
|  | 788811000001108 | Glipizide 5mg tablets (Unichem Plc) (product) |
|  | 795311000001107 | Metformin 850mg tablets (Generics (UK) Ltd) (product) |
|  | 808711000001104 | Minodiab 2.5mg tablets (Pfizer Ltd) (product) |
|  | 8176311000001100 | Avandamet 2mg/1000mg tablets (GlaxoSmithKline) (product) |
|  | 822911000001107 | Glibenclamide 5mg tablets (Kent Pharmaceuticals Ltd) (product) |
|  | 8495311000001100 | Glibenclamide 7.5mg/5ml oral solution (Special Order) (product) |
|  | 8495611000001100 | Glibenclamide 5mg/5ml oral suspension (Special Order) (product) |
|  | 8495911000001100 | Glibenclamide 7.5mg/5ml oral suspension (Special Order) (product) |
|  | 8496611000001100 | Gliclazide 160mg/5ml oral suspension (Special Order) (product) |
|  | 8523711000001100 | Glibenclamide 5mg/5ml oral solution (product) |
|  | 8524011000001100 | Glibenclamide 7.5mg/5ml oral suspension (product) |
|  | 8524311000001100 | Gliclazide 80mg/5ml oral suspension (product) |
|  | 85711000001106 | Metformin 850mg tablets (Unichem Plc) (product) |
|  | 8614411000001100 | Metformin 250mg/5ml oral solution (Special Order) (product) |
|  | 8614811000001100 | Metformin 425mg/5ml oral suspension (Special Order) (product) |
|  | 8615611000001100 | Metformin 250mg/5ml oral suspension (Special Order) (product) |
|  | 8664211000001100 | Metformin 425mg/5ml oral suspension (product) |
|  | 868111000001101 | Vivazide 80mg tablets (Lexon UK Ltd) (product) |
|  | 882811000001109 | Metformin 500mg tablets (Alpharma Limited) (product) |
|  | 887211000001106 | Glibenclamide 5mg tablets (Generics (UK) Ltd) (product) |
|  | 903011000001101 | Glibenclamide 5mg tablets (IVAX Pharmaceuticals UK Ltd) (product) |
|  | 909311000001100 | Metformin 850mg tablets (Sandoz Ltd) (product) |
|  | 922211000001106 | Avandia 4mg tablets (GlaxoSmithKline) (product) |
|  | 929811000001102 | Gliclazide 80mg tablets (Sandoz Ltd) (product) |
|  | 9528811000001100 | Apidra 100units/ml solution for injection 10ml vials (sanofi-aventis) (product) |
|  | 9555911000001100 | Metformin 500mg tablets (Wockhardt UK Ltd) (product) |
|  | 9556111000001100 | Metformin 850mg tablets (Wockhardt UK Ltd) (product) |
|  | 9763311000001100 | Metformin 850mg tablets (The Boots Company) (product) |
|  | 9798211000001100 | Metformin 850mg tablets (Almus Pharmaceutical Ltd) (product) |
|  | 9801011000001100 | Tolbutamide 500mg tablets (Almus Pharmaceutical Ltd) (product) |
|  | 9801911000001100 | Gliclazide 80mg tablets (Almus Pharmaceutical Ltd) (product) |
|  | 34043411000001100 | Fiasp 100units/ml solution for injection 10ml vials (Novo Nordisk Ltd) (product) |
|  | 34188011000001100 | Metformin 850mg/5ml oral solution sugar free (A A H Pharmaceuticals Ltd) (product) |
|  | 34552811000001100 | Meijumet 750mg modified-release tablets (Medreich Plc) (product) |
|  | 34823611000001100 | Gliclazide 40mg tablets (Teva UK Ltd) (product) |
|  | 34956911000001100 | Gliclazide 40mg tablets (Almus Pharmaceuticals Ltd) (product) |
|  | 11119511000001100 | Hypurin Bovine Isophane 100units/ml suspension for injection 10ml vials (Waymade Ltd) (product) |
|  | 11119811000001100 | Hypurin Bovine Isophane 100units/ml suspension for injection 1.5ml cartridges (Waymade Ltd) (product) |
|  | 11982811000001100 | Chlorpropamide 250mg/5ml oral suspension (Special Order) (product) |
|  | 27692911000001100 | Chlorpropamide 100mg tablets (Special Order) (product) |
|  | 325213005 | Product containing precisely chlorpropamide 100 milligram/1 each conventional release oral tablet (clinical drug) |
|  | 325258004 | Product containing precisely tolazamide 250 milligram/1 each conventional release oral tablet (clinical drug) |
|  | 3279311000001100 | Hypurin Bovine Isophane 100units/ml suspension for injection 1.5ml cartridges (C P Pharmaceuticals Ltd) (product) |
|  | 3280011000001100 | Hypurin Bovine Isophane 100units/ml suspension for injection 10ml vials (C P Pharmaceuticals Ltd) (product) |
|  | 3472311000001100 | Insulin isophane bovine 100units/ml suspension for injection 10ml vials (product) |
|  | 374078008 | Product containing precisely tolazamide 500 milligram/1 each conventional release oral tablet (clinical drug) |
|  | 3949611000001100 | Chlorpropamide 100mg tablets (The Boots Company) (product) |
|  | 35214311000001100 | Humulin R KwikPen 500units/ml solution for injection 3ml pre-filled pens (Imported (United States)) (product) |
|  | 35215111000001100 | Insulin human 500units/ml solution for injection 3ml pre-filled disposable devices (product) |
|  | 30933011000001100 | Alogliptin 12.5mg / Metformin 1g tablets (Ennogen Healthcare Ltd) (product) |
|  | 30933211000001100 | Alogliptin 12.5mg tablets (Ennogen Healthcare Ltd) (product) |
|  | 30933611000001100 | Alogliptin 6.25mg tablets (Ennogen Healthcare Ltd) (product) |
|  | 32182811000001100 | Alogliptin 25mg tablets (Niche Pharma Ltd) (product) |
|  | 32183011000001100 | Alogliptin 6.25mg tablets (Niche Pharma Ltd) (product) |
|  | 35316811000001100 | Gliclazide 40mg tablets (NorthStar Healthcare Unlimited Company) (product) |
|  | 35547511000001100 | Yaltormin SR 500mg tablets (Wockhardt UK Ltd) (product) |
|  | 35548011000001100 | Yaltormin SR 750mg tablets (Wockhardt UK Ltd) (product) |
|  | 35563311000001100 | Insulin lispro 100units/ml solution for injection 3ml cartridges (Sanofi Pasteur) (product) |
|  | 35593311000001100 | Metformin 500mg tablets (RX Farma) (product) |
|  | 35593611000001100 | Metformin 850mg tablets (RX Farma) (product) |
|  | 35653211000001100 | Pioglitazone 15mg / Metformin 850mg tablets (Teva UK Ltd) (product) |
|  | 35672911000001100 | Pioglitazone 15mg / Metformin 850mg tablets (A A H Pharmaceuticals Ltd) (product) |
|  | 35776511000001100 | Insulin lispro Sanofi 100units/ml solution for injection 3ml cartridges (Sanofi) (product) |
|  | 35849011000001100 | Metuxtan SR 500mg tablets (Accord Healthcare Ltd) (product) |
|  | 36047011000001100 | Insulin aspart 100units/ml solution for injection 10ml vials (product) |
|  | 36047111000001100 | Insulin glargine 100units/ml solution for injection 10ml vials (product) |
|  | 36047211000001100 | Insulin glulisine 100units/ml solution for injection 10ml vials (product) |
|  | 36047711000001100 | Insulin lispro 100units/ml solution for injection 3ml cartridges (product) |
|  | 36047811000001100 | Insulin lispro 100units/ml solution for injection 3ml pre-filled disposable devices (product) |
|  | 36048011000001100 | Insulin soluble bovine 100units/ml solution for injection 1.5ml cartridges (product) |
|  | 36048211000001100 | Insulin soluble human 100units/ml solution for injection 1.5ml cartridges (product) |
|  | 36048311000001100 | Insulin soluble human 100units/ml solution for injection 10ml vials (product) |
|  | 36048711000001100 | Insulin soluble porcine 100units/ml solution for injection 10ml vials (product) |
|  | 36048911000001100 | Insulin zinc mixed human 100units/ml suspension for injection 10ml vials (product) |
|  | 36082811000001100 | Semglee 100units/ml solution for injection 3ml pre-filled pens (Mylan) (product) |
|  | 36468511000001100 | Pioglitazone 15mg / Metformin 850mg tablets (Alliance Healthcare (Distribution) Ltd) (product) |
|  | 36739111000001100 | Gliclazide 40mg tablets (Flamingo Pharma (UK) Ltd) (product) |
|  | 36763511000001100 | Byetta 10micrograms/0.04ml solution for injection 2.4ml pre-filled pens (Originalis B.V.) (product) |
|  | 36804511000001100 | Jardiance 10mg tablets (Originalis B.V.) (product) |
|  | 36832811000001100 | Pioglitazone 15mg / Metformin 850mg tablets (Torrent Pharma (UK) Ltd) (product) |
|  | 36856611000001100 | Pioglitazone 15mg tablets (Torrent Pharma (UK) Ltd) (product) |
|  | 36857111000001100 | Pioglitazone 30mg tablets (Torrent Pharma (UK) Ltd) (product) |
|  | 36893111000001100 | Forxiga 10mg tablets (Pharmaram Ltd) (product) |
|  | 36906311000001100 | Vipidia 6.25mg tablets (Originalis B.V.) (product) |
|  | 36911311000001100 | Toujeo 300units/ml solution for injection 3ml pre-filled DoubleStar pens (Sanofi) (product) |
|  | 36914911000001100 | Gliclazide 160mg tablets (product) |
|  | 36925911000001100 | Gliclazide 80mg tablets (Relonchem Ltd) (product) |
|  | 36931811000001100 | Insulin glargine 300units/ml solution for injection 3ml pre-filled disposable devices (product) |
|  | 37069611000001100 | Glimepiride 1mg tablets (Mawdsley-Brooks & Company Ltd) (product) |
|  | 37120411000001100 | Metformin 500mg tablets (Mawdsley-Brooks & Company Ltd) (product) |
|  | 36630511000001100 | Insulin glargine 100units/ml / Lixisenatide 33micrograms/ml solution for injection 3ml pre-filled disposable devices (product) |
|  | 36630611000001100 | Insulin glargine 100units/ml / Lixisenatide 50micrograms/ml solution for injection 3ml pre-filled disposable devices (product) |

|  | 37337011000001100 | Amglidia 0.6mg/ml oral suspension with 1ml oral syringe (Amring Pharmaceuticals Ltd) (product) |
| --- | --- | --- |
|  | 37337211000001100 | Amglidia 0.6mg/ml oral suspension with 5ml oral syringe (Amring Pharmaceuticals Ltd) (product) |
|  | 37337711000001100 | Amglidia 6mg/ml oral suspension with 5ml oral syringe (Amring Pharmaceuticals Ltd) (product) |
|  | 37355411000001100 | Byetta 10micrograms/0.04ml solution for injection 2.4ml pre-filled pens (Mawdsley-Brooks & Company Ltd) (product) |
|  | 37387311000001100 | Gliclazide 40mg tablets (DE Pharmaceuticals) (product) |
|  | 37405911000001100 | Glibenclamide 600micrograms/ml oral suspension sugar free (product) |
|  | 37417011000001100 | Humulin I 100units/ml suspension for injection 3ml cartridges (CST Pharma Ltd) (product) |
|  | 37417211000001100 | Humulin M3 100units/ml suspension for injection 3ml cartridges (CST Pharma Ltd) (product) |
|  | 37420411000001100 | Humulin I 100units/ml suspension for injection 10ml vials (CST Pharma Ltd) (product) |
|  | 37428011000001100 | Actos 15mg tablets (CST Pharma Ltd) (product) |
|  | 37428411000001100 | Actos 45mg tablets (CST Pharma Ltd) (product) |
|  | 37428811000001100 | Apidra 100units/ml solution for injection 3ml pre-filled SoloStar pens (CST Pharma Ltd) (product) |
|  | 37438411000001100 | Galvus 50mg tablets (CST Pharma Ltd) (product) |
|  | 37439111000001100 | Humalog KwikPen 100units/ml solution for injection 3ml pre-filled pens (CST Pharma Ltd) (product) |
|  | 37440411000001100 | Januvia 25mg tablets (CST Pharma Ltd) (product) |
|  | 37440811000001100 | Jardiance 10mg tablets (CST Pharma Ltd) (product) |
|  | 37441111000001100 | Jardiance 25mg tablets (CST Pharma Ltd) (product) |
|  | 37508611000001100 | Glucophage SR 750mg tablets (Mawdsley-Brooks & Company Ltd) (product) |
|  | 37508911000001100 | Glucophage 500mg tablets (Mawdsley-Brooks & Company Ltd) (product) |
|  | 37512111000001100 | Apidra 100units/ml solution for injection 3ml pre-filled SoloStar pens (Mawdsley-Brooks & Company Ltd) (product) |
|  | 37512611000001100 | Invokana 100mg tablets (Mawdsley-Brooks & Company Ltd) (product) |
|  | 37525711000001100 | Januvia 50mg tablets (Mawdsley-Brooks & Company Ltd) (product) |
|  | 37527211000001100 | Onglyza 2.5mg tablets (CST Pharma Ltd) (product) |
|  | 37527411000001100 | Onglyza 5mg tablets (CST Pharma Ltd) (product) |
|  | 37537911000001100 | Vipidia 12.5mg tablets (CST Pharma Ltd) (product) |
|  | 37538511000001100 | Xigduo 5mg/1000mg tablets (CST Pharma Ltd) (product) |
|  | 37550411000001100 | Humalog Mix50 KwikPen 100units/ml suspension for injection 3ml pre-filled pens (CST Pharma Ltd) (product) |
|  | 37550911000001100 | Humalog Mix25 100units/ml suspension for injection 3ml cartridges (CST Pharma Ltd) (product) |
|  | 37665511000001100 | Forxiga 10mg tablets (Pilsco Ltd) (product) |
|  | 37666211000001100 | Humalog Mix25 100units/ml suspension for injection 3ml cartridges (Pilsco Ltd) (product) |
|  | 37667011000001100 | Invokana 100mg tablets (Pilsco Ltd) (product) |
|  | 37667311000001100 | Janumet 50mg/1000mg tablets (Pilsco Ltd) (product) |
|  | 37667611000001100 | Januvia 100mg tablets (Pilsco Ltd) (product) |
|  | 37668011000001100 | Januvia 25mg tablets (Pilsco Ltd) (product) |
|  | 37668211000001100 | Januvia 50mg tablets (Pilsco Ltd) (product) |
|  | 37668511000001100 | Jardiance 10mg tablets (Pilsco Ltd) (product) |
|  | 37672211000001100 | Lantus 100units/ml solution for injection 3ml cartridges (Pilsco Ltd) (product) |
|  | 37695011000001100 | NovoMix 30 Penfill 100units/ml suspension for injection 3ml cartridges (Pilsco Ltd) (product) |
|  | 37696411000001100 | Onglyza 5mg tablets (Pilsco Ltd) (product) |
|  | 37708911000001100 | Victoza 6mg/ml solution for injection 3ml pre-filled pens (Pilsco Ltd) (product) |
|  | 37709511000001100 | Vipidia 25mg tablets (Pilsco Ltd) (product) |
|  | 37719711000001100 | Humulin M3 100units/ml suspension for injection 10ml vials (Pilsco Ltd) (product) |
|  | 37747911000001100 | Jentadueto 2.5mg/1000mg tablets (CST Pharma Ltd) (product) |
|  | 37751111000001100 | Tresiba FlexTouch 200units/ml solution for injection 3ml pre-filled pens (CST Pharma Ltd) (product) |
|  | 37775311000001100 | Pioglitazone 15mg / Metformin 850mg tablets (DE Pharmaceuticals) (product) |
|  | 10925511000001100 | Metformin 850mg / Pioglitazone 15mg tablets (product) |
|  | 11473511000001100 | Sitagliptin 100mg tablets (product) |
|  | 16037911000001100 | Saxagliptin 5mg tablets (product) |
|  | 325063005 | Insulin soluble human 100units/mL injection solution 3mL cartridge (product) |
|  | 325064004 | Insulin soluble human 100units/mL injection solution 10mL vial (product) |
|  | 325067006 | Insulin lispro 100units/mL injection solution 3mL cartridge (product) |
|  | 325070005 | Insulin lispro 100units/mL injection solution 3mL prefilled pen (product) |
|  | 325076004 | Insulin aspart 100units/mL injection solution 10mL vial (product) |
|  | 325111000 | Insulin isophane human 100units/mL injection suspension 3mL cartridge (product) |
|  | 3469111000001100 | Insulin glargine 100units/ml solution for injection 3ml cartridges (product) |
|  | 353987005 | Insulin soluble human 100units/mL injection solution 3mL prefilled pen (product) |
|  | 353988000 | Insulin isophane human 100units/mL injection suspension 3mL prefilled pen (product) |
|  | 371418007 | Insulin soluble bovine 100u/mL injection solution 10mL vial (product) |
|  | 371419004 | Insulin soluble bovine 100u/mL injection solution 1.5mL cartridge (product) |
|  | 371430001 | Insulin soluble porcine 100u/mL injection solution 10mL vial (product) |
|  | 371483009 | Insulin soluble human 100u/mL injection solution 5mL vial (product) |
|  | 371496005 | Insulin lispro 100u/mL injection solution 1.5mL cartridge (product) |
|  | 37853211000001100 | Synjardy 12.5mg/1000mg tablets (CST Pharma Ltd) (product) |
|  | 412448008 | Insulin glulisine 100units/mL injection 10mL vial (product) |
|  | 5322611000001100 | Metformin 500mg / Rosiglitazone 1mg tablets (product) |
|  | 8175511000001100 | Metformin 1g / Rosiglitazone 4mg tablets (product) |
|  | 8285611000001100 | Metformin 1g / Rosiglitazone 2mg tablets (product) |
|  | 37972011000001100 | Humalog 100units/ml solution for injection 10ml vials (Pharmaram Ltd) (product) |
|  | 37972211000001100 | Humalog Mix25 100units/ml suspension for injection 3ml cartridges (Pharmaram Ltd) (product) |
|  | 37973311000001100 | NovoRapid Penfill 100units/ml solution for injection 3ml cartridges (Pharmaram Ltd) (product) |
|  | 37974511000001100 | Vipidia 12.5mg tablets (Pharmaram Ltd) (product) |
|  | 37989611000001100 | Invokana 100mg tablets (Pharmaram Ltd) (product) |
|  | 37989911000001100 | Janumet 50mg/1000mg tablets (Pharmaram Ltd) (product) |
|  | 37990211000001100 | Januvia 100mg tablets (Pharmaram Ltd) (product) |
|  | 38060511000001100 | Bydureon BCise 2mg/0.85ml prolonged-release suspension for injection pre-filled pens (AstraZeneca UK Ltd) (product) |
|  | 38067111000001100 | Insulin soluble human 1unit/ml solution for infusion 30ml pre-filled syringes (Special Order) (product) |
|  | 38067711000001100 | Insulin soluble human 1unit/ml solution for infusion 30ml pre-filled syringes (product) |
|  | 38116611000001100 | Apidra 100units/ml solution for injection 3ml pre-filled SoloStar pens (DE Pharmaceuticals) (product) |
|  | 38123611000001100 | Byetta 10micrograms/0.04ml solution for injection 2.4ml pre-filled pens (DE Pharmaceuticals) (product) |
|  | 38135711000001100 | Forxiga 10mg tablets (DE Pharmaceuticals) (product) |
|  | 38138111000001100 | Glucophage SR 750mg tablets (DE Pharmaceuticals) (product) |
|  | 38138411000001100 | Glucophage SR 1000mg tablets (DE Pharmaceuticals) (product) |
|  | 38139711000001100 | Humulin M3 100units/ml suspension for injection 3ml cartridges (DE Pharmaceuticals) (product) |
|  | 38140911000001100 | Invokana 100mg tablets (DE Pharmaceuticals) (product) |
|  | 38141511000001100 | Janumet 50mg/1000mg tablets (DE Pharmaceuticals) (product) |
|  | 38141911000001100 | Januvia 50mg tablets (DE Pharmaceuticals) (product) |
|  | 38143211000001100 | Jentadueto 2.5mg/1000mg tablets (DE Pharmaceuticals) (product) |
|  | 38157811000001100 | Onglyza 2.5mg tablets (DE Pharmaceuticals) (product) |
|  | 38170511000001100 | Toujeo 300units/ml solution for injection 1.5ml pre-filled SoloStar pens (DE Pharmaceuticals) (product) |
|  | 38170711000001100 | Trajenta 5mg tablets (DE Pharmaceuticals) (product) |
|  | 38171211000001100 | Tresiba FlexTouch 200units/ml solution for injection 3ml pre-filled pens (DE Pharmaceuticals) (product) |
|  | 38174811000001100 | Vipidia 6.25mg tablets (DE Pharmaceuticals) (product) |
|  | 38175011000001100 | Vipidia 12.5mg tablets (DE Pharmaceuticals) (product) |
|  | 38176411000001100 | Xigduo 5mg/1000mg tablets (DE Pharmaceuticals) (product) |
|  | 38238111000001100 | Sukkarto SR 750mg tablets (Morningside Healthcare Ltd) (product) |
|  | 38292411000001100 | Acarbose 50mg tablets (Rivopharm (UK) Ltd) (product) |
|  | 38379411000001100 | Saxenda 6mg/ml solution for injection 3ml pre-filled pens (CST Pharma Ltd) (product) |
|  | 38533711000001100 | Metformin 750mg modified-release tablets (Morningside Healthcare Ltd) (product) |
|  | 10093311000001100 | Apidra 100units/ml solution for injection 3ml pre-filled OptiSet pens (Sanofi) (product) |
|  | 10097211000001100 | Insulin glulisine 100units/ml solution for injection 3ml pre-filled disposable devices (product) |
|  | 10250211000001100 | Lantus OptiClik 100units/ml solution for injection 3ml cartridges (sanofi-aventis) (product) |
|  | 10272911000001100 | Glimepiride 4mg tablets (Unichem Plc) (product) |
|  | 10274011000001100 | Glimepiride 2mg tablets (A A H Pharmaceuticals Ltd) (product) |
|  | 10292611000001100 | Metformin 500mg tablets (Relonchem Ltd) (product) |
|  | 10344911000001100 | Humalog Mix50 100units/ml suspension for injection 3ml cartridges (Eli Lilly & Co Ltd) (product) |
|  | 10352711000001100 | Insulin lispro biphasic 50/50 100units/ml suspension for injection 3ml cartridges (product) |
|  | 10445111000001100 | Glimepiride 1mg tablets (Niche Generics Ltd) (product) |
|  | 10445311000001100 | Glimepiride 2mg tablets (Niche Generics Ltd) (product) |
|  | 10445511000001100 | Glimepiride 3mg tablets (Niche Generics Ltd) (product) |
|  | 10459011000001100 | Actos 30mg tablets (PI) (Dowelhurst Ltd) (product) |
|  | 10464711000001100 | Avandia 4mg tablets (PI) (Dowelhurst Ltd) (product) |
|  | 10470611000001100 | Avandamet 2mg/500mg tablets (PI) (Dowelhurst Ltd) (product) |
|  | 10486711000001100 | Avandamet 1mg/500mg tablets (PI) (Waymade Ltd) (product) |
|  | 10486811000001100 | NovoNorm 1mg tablets (PI) (Dowelhurst Ltd) (product) |
|  | 10487411000001100 | Avandamet 2mg/1000mg tablets (PI) (Waymade Ltd) (product) |
|  | 10487711000001100 | Avandamet 2mg/500mg tablets (PI) (Waymade Ltd) (product) |
|  | 10508511000001100 | Glibenese 5mg tablets (PI) (Dowelhurst Ltd) (product) |
|  | 10517911000001100 | Glimepiride 2mg tablets (Winthrop Pharmaceuticals UK Ltd) (product) |
|  | 10529511000001100 | Diamicron 30mg MR tablets (PI) (Waymade Ltd) (product) |
|  | 10534811000001100 | NovoRapid 100units/ml solution for injection 10ml vials (PI) (Waymade Ltd) (product) |
|  | 10542211000001100 | Humalog Pen 100units/ml solution for injection 3ml pre-filled pens (Waymade Healthcare Plc) (product) |
|  | 10542911000001100 | NovoMix 30 Penfill 100units/ml suspension for injection 3ml cartridges (PI) (Waymade Ltd) (product) |
|  | 10617311000001100 | Glimepiride 1mg tablets (Somex Pharma) (product) |
|  | 10617811000001100 | Glimepiride 3mg tablets (Somex Pharma) (product) |
|  | 10618111000001100 | Glimepiride 4mg tablets (Somex Pharma) (product) |
|  | 10688811000001100 | Glimepiride 1mg tablets (Pliva Pharma Ltd) (product) |
|  | 10690511000001100 | Exubera 1mg inhalation powder blisters (Pfizer Ltd) (product) |
|  | 10690811000001100 | Exubera 3mg inhalation powder blisters (Pfizer Ltd) (product) |
|  | 10703011000001100 | Insulin human 1mg inhalation powder blisters (product) |
|  | 10703111000001100 | Insulin human 3mg inhalation powder blisters (product) |
|  | 107311000001107 | Glibenclamide 2.5mg tablets (Kent Pharmaceuticals Ltd) (product) |
|  | 10952411000001100 | Prandin 0.5mg tablets (Novo Nordisk Ltd) (product) |
|  | 10957811000001100 | Prandin 2mg tablets (Novo Nordisk Ltd) (product) |
|  | 11018611000001100 | Glimepiride 3mg tablets (Dr Reddy's Labs) (product) |
|  | 11113611000001100 | Pork Actrapid 100units/ml solution for injection 10ml vials (Waymade Ltd) (product) |
|  | 11114411000001100 | Pork Insulatard 100units/ml suspension for injection 10ml vials (Waymade Ltd) (product) |
|  | 11115011000001100 | Levemir FlexPen 100units/ml solution for injection 3ml pre-filled pens (Waymade Healthcare Plc) (product) |
|  | 11116811000001100 | Hypurin Porcine Neutral 100units/ml solution for injection 10ml vials (Waymade Ltd) (product) |
|  | 11117211000001100 | Hypurin Porcine Neutral 100units/ml solution for injection 1.5ml cartridges (Waymade Ltd) (product) |
|  | 11118411000001100 | Hypurin Porcine Isophane 100units/ml suspension for injection 10ml vials (Waymade Ltd) (product) |
|  | 11119311000001100 | Hypurin Bovine Lente 100units/ml suspension for injection 10ml vials (Waymade Ltd) (product) |
|  | 11122611000001100 | Mixtard 50 Penfill 100units/ml suspension for injection 3ml cartridges (Waymade Ltd) (product) |
|  | 11123011000001100 | Human Mixtard 50 Penfill 100units/ml suspension for injection 1.5ml cartridges (Waymade Ltd) (product) |
|  | 11123311000001100 | Mixtard 40 Penfill 100units/ml suspension for injection 3ml cartridges (Waymade Ltd) (product) |
|  | 11123611000001100 | Human Mixtard 40 Penfill 100units/ml suspension for injection 1.5ml cartridges (Waymade Ltd) (product) |
|  | 11124211000001100 | Mixtard 30 Penfill 100units/ml suspension for injection 3ml cartridges (Waymade Ltd) (product) |
|  | 11124611000001100 | Mixtard 20 Penfill 100units/ml suspension for injection 3ml cartridges (Waymade Ltd) (product) |
|  | 11132611000001100 | Actrapid 100units/ml solution for injection 10ml vials (Waymade Ltd) (product) |
|  | 11133811000001100 | Insulatard InnoLet 100units/ml suspension for injection 3ml pre-filled pens (Waymade Healthcare Plc) (product) |
|  | 11134011000001100 | Insulatard Penfill 100units/ml suspension for injection 3ml cartridges (Waymade Ltd) (product) |
|  | 11135211000001100 | Human Mixtard 10 Penfill 100units/ml suspension for injection 1.5ml cartridges (Waymade Ltd) (product) |
|  | 11135411000001100 | Human Mixtard 20 Penfill 100units/ml suspension for injection 1.5ml cartridges (Waymade Ltd) (product) |
|  | 11149811000001100 | Glimepiride 4mg tablets (Teva UK Ltd) (product) |
|  | 11343211000001100 | Glimepiride 2mg tablets (Sandoz Ltd) (product) |
|  | 11344011000001100 | Glimepiride 4mg tablets (Sandoz Ltd) (product) |
|  | 11411000001101 | Gliclazide 80mg tablets (Kent Pharmaceuticals Ltd) (product) |
|  | 11494111000001100 | Exenatide 10micrograms/0.04ml solution for injection 2.4ml pre-filled disposable devices (product) |
|  | 11494211000001100 | Exenatide 5micrograms/0.02ml solution for injection 1.2ml pre-filled disposable devices (product) |
|  | 115811000001106 | Metformin 850mg tablets (Alpharma Limited) (product) |
|  | 11599411000001100 | Metformin 500mg/5ml oral solution sugar free (Rosemont Pharmaceuticals Ltd) (product) |
|  | 11765811000001100 | Prandin 0.5mg tablets (Waymade Healthcare Plc) (product) |
|  | 120811000001108 | Glibenclamide 5mg tablets (C P Pharmaceuticals Ltd) (product) |
|  | 12107211000001100 | Glibenclamide 625micrograms/5ml oral suspension (Special Order) (product) |
|  | 12107811000001100 | Glibenclamide 15mg/5ml oral suspension (Special Order) (product) |
|  | 12108111000001100 | Glibenclamide 15mg/5ml oral solution (Special Order) (product) |
|  | 12109011000001100 | Gliclazide 100mg/5ml oral suspension (Special Order) (product) |
|  | 12109311000001100 | Gliclazide 120mg/5ml oral suspension (Special Order) (product) |
|  | 12109611000001100 | Gliclazide 20mg/5ml oral suspension (Special Order) (product) |
|  | 12135311000001100 | Glibenclamide 15mg/5ml oral solution (product) |
|  | 12135711000001100 | Glibenclamide 625micrograms/5ml oral solution (product) |
|  | 12136011000001100 | Gliclazide 120mg/5ml oral suspension (product) |
|  | 12136411000001100 | Gliclazide 60mg/5ml oral suspension (product) |
|  | 12147111000001100 | Glimepiride 1mg tablets (Kent Pharmaceuticals Ltd) (product) |
|  | 12147511000001100 | Glimepiride 3mg tablets (Kent Pharmaceuticals Ltd) (product) |
|  | 12147711000001100 | Glimepiride 4mg tablets (Kent Pharmaceuticals Ltd) (product) |
|  | 12792711000001100 | Metformin 1g/5ml oral solution (Special Order) (product) |
|  | 12793011000001100 | Metformin 5mg/5ml oral solution (Special Order) (product) |
|  | 12813711000001100 | Metformin 5mg/5ml oral solution (product) |
|  | 129411000001101 | Glipizide 5mg tablets (Pfizer Ltd) (product) |
|  | 13100411000001100 | Avandamet 2mg/1000mg tablets (Dowelhurst Ltd) (product) |
|  | 13158411000001100 | Diamicron 30mg MR tablets (Dowelhurst Ltd) (product) |
|  | 13168311000001100 | Januvia 100mg tablets (Dowelhurst Ltd) (product) |
|  | 13180011000001100 | NovoMix 30 FlexPen 100units/ml suspension for injection 3ml pre-filled pens (Dowelhurst Ltd) (product) |
|  | 13181211000001100 | NovoRapid FlexPen 100units/ml solution for injection 3ml pre-filled pens (Dowelhurst Ltd) (product) |
|  | 13182011000001100 | NovoRapid Penfill 100units/ml solution for injection 3ml cartridges (Dowelhurst Ltd) (product) |
|  | 13205311000001100 | Starlix 120mg tablets (Dowelhurst Ltd) (product) |
|  | 13317511000001100 | Tolbutamide 1g/5ml oral suspension (Special Order) (product) |
|  | 13324311000001100 | Tolbutamide 1g/5ml oral suspension (product) |
|  | 13324411000001100 | Tolbutamide 250mg/5ml oral suspension (product) |
|  | 13412311000001100 | Eucreas 50mg/1000mg tablets (Novartis Pharmaceuticals UK Ltd) (product) |
|  | 13412611000001100 | Eucreas 50mg/850mg tablets (Novartis Pharmaceuticals UK Ltd) (product) |
|  | 13412911000001100 | Vildagliptin 50mg tablets (product) |
|  | 13413011000001100 | Metformin 1g / Vildagliptin 50mg tablets (product) |
|  | 13435011000001100 | Pioglitazone 30mg/5ml oral suspension (Special Order) (product) |
|  | 13440011000001100 | Pioglitazone 30mg/5ml oral suspension (product) |
|  | 134609007 | Product containing precisely nateglinide 120 milligram/1 each conventional release oral tablet (clinical drug) |
|  | 134610002 | Product containing precisely nateglinide 60 milligram/1 each conventional release oral tablet (clinical drug) |
|  | 13578511000001100 | Glipizide 5mg tablets (Doncaster Pharmaceuticals Ltd) (product) |
|  | 13588911000001100 | Gliclazide 30mg modified-release tablets (UniChem Ltd) (product) |
|  | 13626511000001100 | Niddaryl 2mg tablets (Dee Pharmaceuticals Ltd) (product) |
|  | 13626911000001100 | Niddaryl 3mg tablets (Dee Pharmaceuticals Ltd) (product) |
|  | 13720511000001100 | Glimepiride 2mg tablets (Ranbaxy (UK) Ltd) (product) |
|  | 13721211000001100 | Glimepiride 4mg tablets (Ranbaxy (UK) Ltd) (product) |
|  | 13757811000001100 | Metformin 850mg tablets (Tillomed Laboratories Ltd) (product) |
|  | 13772811000001100 | Humulin I 100units/ml suspension for injection 3ml cartridges (Waymade Healthcare Plc) (product) |
|  | 13773011000001100 | Humulin S 100units/ml solution for injection 3ml cartridges (Waymade Healthcare Plc) (product) |
|  | 13833111000001100 | Avandamet 2mg/500mg tablets (Doncaster Pharmaceuticals Ltd) (product) |
|  | 13833811000001100 | Avandia 4mg tablets (Doncaster Pharmaceuticals Ltd) (product) |
|  | 13834011000001100 | Avandia 8mg tablets (Doncaster Pharmaceuticals Ltd) (product) |
|  | 13850511000001100 | Humulin R 100units/ml solution for injection 10ml vials (Imported (United States)) (product) |
|  | 13883511000001100 | Glucophage 500mg tablets (Doncaster Pharmaceuticals Ltd) (product) |
|  | 13883911000001100 | Glucophage 850mg tablets (Doncaster Pharmaceuticals Ltd) (product) |
|  | 13884711000001100 | Humalog Mix25 KwikPen 100units/ml suspension for injection 3ml pre-filled pens (Eli Lilly and Company Ltd) (product) |
|  | 14040411000001100 | Actos 30mg tablets (Sigma Pharmaceuticals Plc) (product) |
|  | 14056511000001100 | Amaryl 2mg tablets (Sigma Pharmaceuticals Plc) (product) |
|  | 140711000001103 | Semi-Daonil 2.5mg tablets (Aventis Pharma) (product) |
|  | 14122311000001100 | Glimepiride 2mg tablets (Consilient Health Ltd) (product) |
|  | 14122711000001100 | Glimepiride 4mg tablets (Consilient Health Ltd) (product) |
|  | 14183911000001100 | Bolamyn SR 500mg tablets (Teva UK Ltd) (product) |
|  | 14199911000001100 | Avandamet 2mg/1000mg tablets (Sigma Pharmaceuticals Plc) (product) |
|  | 14200111000001100 | Avandamet 4mg/1000mg tablets (Sigma Pharmaceuticals Plc) (product) |
|  | 14248411000001100 | Humalog Mix50 100units/ml suspension for injection 3ml cartridges (Sigma Pharmaceuticals Plc) (product) |
|  | 14248611000001100 | Humalog Mix25 Pen 100units/ml suspension for injection 3ml pre-filled pens (Sigma Pharmaceuticals Plc) (product) |
|  | 14249011000001100 | Humalog Pen 100units/ml solution for injection 3ml pre-filled pens (Sigma Pharmaceuticals Plc) (product) |
|  | 14249211000001100 | Humalog 100units/ml solution for injection 10ml vials (Sigma Pharmaceuticals Plc) (product) |
|  | 14313211000001100 | Diamicron 30mg MR tablets (Sigma Pharmaceuticals Plc) (product) |
|  | 14400811000001100 | Starlix 120mg tablets (Sigma Pharmaceuticals Plc) (product) |
|  | 14401211000001100 | Starlix 180mg tablets (Sigma Pharmaceuticals Plc) (product) |
|  | 14584711000001100 | Metformin 500mg modified-release tablets (A A H Pharmaceuticals Ltd) (product) |
|  | 14592411000001100 | Humulin I 100units/ml suspension for injection 10ml vials (Waymade Healthcare Plc) (product) |
|  | 14665711000001100 | NovoRapid 100units/ml solution for injection 10ml vials (Sigma Pharmaceuticals Plc) (product) |
|  | 14711111000001100 | Minodiab 5mg tablets (Sigma Pharmaceuticals Plc) (product) |
|  | 14759411000001100 | Lantus 100units/ml solution for injection 3ml cartridges (Sigma Pharmaceuticals Plc) (product) |
|  | 14773811000001100 | Metformin 500mg tablets (LPC Medical (UK) Ltd) (product) |
|  | 15090711000001100 | Gliclazide 30mg modified-release tablets (Sigma Pharmaceuticals Plc) (product) |
|  | 15091211000001100 | Glimepiride 2mg tablets (Sigma Pharmaceuticals Plc) (product) |
|  | 15091611000001100 | Glimepiride 4mg tablets (Sigma Pharmaceuticals Plc) (product) |
|  | 15334511000001100 | Edicil MR 30mg tablets (Ratiopharm UK Ltd) (product) |
|  | 15359011000001100 | Competact 15mg/850mg tablets (Waymade Healthcare Plc) (product) |
|  | 15367811000001100 | Glucophage SR 1000mg tablets (Merck Serono Ltd) (product) |
|  | 15373711000001100 | Glucophage 500mg oral powder sachets (Merck Serono Ltd) (product) |
|  | 15411311000001100 | Metformin 500mg oral powder sachets sugar free (product) |
|  | 15611411000001100 | Insulin human 500units/ml solution for injection 20ml vials (product) |
|  | 157911000001103 | Metformin 850mg tablets (Approved Prescription Services) (product) |
|  | 15859111000001100 | Liraglutide 18mg/3ml solution for injection pre-filled disposable devices (product) |
|  | 15973311000001100 | Metformin 500mg tablets (Zanza Laboratories Ltd) (product) |
|  | 15984111000001100 | Metformin 500mg/5ml oral solution sugar free (Actavis UK Ltd) (product) |
|  | 15993011000001100 | Onglyza 5mg tablets (Bristol-Myers Squibb Pharmaceuticals Ltd) (product) |
|  | 16007611000001100 | Pioglitazone 15mg/5ml oral suspension (Special Order) (product) |
|  | 16036811000001100 | Pioglitazone 15mg/5ml oral suspension (product) |
|  | 16053211000001100 | Gliclazide 80mg tablets (Bristol Laboratories Ltd) (product) |
|  | 16053511000001100 | Glimepiride 1mg tablets (Bristol Laboratories Ltd) (product) |
|  | 16053911000001100 | Glimepiride 3mg tablets (Bristol Laboratories Ltd) (product) |
|  | 16061711000001100 | Metformin 850mg tablets (Bristol Laboratories Ltd) (product) |
|  | 16125711000001100 | Glimepiride 2mg/5ml oral suspension (Special Order) (product) |
|  | 16137611000001100 | Amaryl 3mg tablets (Lexon (UK) Ltd) (product) |
|  | 16137811000001100 | Amaryl 4mg tablets (Lexon (UK) Ltd) (product) |
|  | 16158211000001100 | Diamicron 30mg MR tablets (Lexon (UK) Ltd) (product) |
|  | 16187611000001100 | Glucophage 850mg tablets (Lexon (UK) Ltd) (product) |
|  | 16256011000001100 | Actos 15mg tablets (Mawdsley-Brooks & Company Ltd) (product) |
|  | 16256611000001100 | Diamicron 30mg MR tablets (Mawdsley-Brooks & Company Ltd) (product) |
|  | 16271111000001100 | Avandamet 2mg/500mg tablets (Mawdsley-Brooks & Company Ltd) (product) |
|  | 16272111000001100 | Avandamet 4mg/1000mg tablets (Mawdsley-Brooks & Company Ltd) (product) |
|  | 16273111000001100 | Avandia 8mg tablets (Mawdsley-Brooks & Company Ltd) (product) |
|  | 164511000001108 | Amaryl 3mg tablets (Aventis Pharma) (product) |
|  | 16503111000001100 | Glucophage SR 500mg tablets (Mawdsley-Brooks & Company Ltd) (product) |
|  | 16516311000001100 | Humalog 100units/ml solution for injection 3ml cartridges (Mawdsley-Brooks & Company Ltd) (product) |
|  | 16518411000001100 | Humalog Mix25 Pen 100units/ml suspension for injection 3ml pre-filled pens (Mawdsley-Brooks & Company Ltd) (product) |
|  | 16519811000001100 | Mixtard 30 Penfill 100units/ml suspension for injection 3ml cartridges (Mawdsley-Brooks & Company Ltd) (product) |
|  | 16520511000001100 | Humulin M3 100units/ml suspension for injection 10ml vials (Mawdsley-Brooks & Company Ltd) (product) |
|  | 16521911000001100 | Humulin S 100units/ml solution for injection 10ml vials (Mawdsley-Brooks & Company Ltd) (product) |
|  | 16522311000001100 | NovoRapid FlexPen 100units/ml solution for injection 3ml pre-filled pens (Mawdsley-Brooks & Company Ltd) (product) |
|  | 16522711000001100 | NovoRapid Penfill 100units/ml solution for injection 3ml cartridges (Mawdsley-Brooks & Company Ltd) (product) |
|  | 16701411000001100 | Metformin 850mg tablets (Pfizer Ltd) (product) |
|  | 16702011000001100 | Gliclazide 40mg tablets (product) |
|  | 16727911000001100 | Levemir InnoLet 100units/ml solution for injection 3ml pre-filled pens (Waymade Healthcare Plc) (product) |
|  | 17199311000001100 | Gliclazide 80mg tablets (Sovereign Medical Ltd) (product) |
|  | 17205711000001100 | Tolbutamide 500mg tablets (Sovereign Medical Ltd) (product) |
|  | 17496211000001100 | Byetta 5micrograms/0.02ml solution for injection 1.2ml pre-filled pens (Mawdsley-Brooks & Company Ltd) (product) |
|  | 17503511000001100 | Eucreas 50mg/1000mg tablets (Mawdsley-Brooks & Company Ltd) (product) |
|  | 17503711000001100 | Eucreas 50mg/850mg tablets (Mawdsley-Brooks & Company Ltd) (product) |
|  | 17506511000001100 | Galvus 50mg tablets (Mawdsley-Brooks & Company Ltd) (product) |
|  | 17509911000001100 | Apidra 100units/ml solution for injection 3ml cartridges (Mawdsley-Brooks & Company Ltd) (product) |
|  | 17510911000001100 | Levemir Penfill 100units/ml solution for injection 3ml cartridges (Mawdsley-Brooks & Company Ltd) (product) |
|  | 17511111000001100 | Levemir FlexPen 100units/ml solution for injection 3ml pre-filled pens (Mawdsley-Brooks & Company Ltd) (product) |
|  | 17558911000001100 | Glucobay 100mg tablets (Sigma Pharmaceuticals Plc) (product) |
|  | 17559111000001100 | Humulin I 100units/ml suspension for injection 3ml cartridges (Sigma Pharmaceuticals Plc) (product) |
|  | 17567211000001100 | Januvia 100mg tablets (Sigma Pharmaceuticals Plc) (product) |
|  | 17581511000001100 | Amaryl 1mg tablets (Necessity Supplies Ltd) (product) |
|  | 17582111000001100 | Amaryl 2mg tablets (Necessity Supplies Ltd) (product) |
|  | 17582711000001100 | Amaryl 4mg tablets (Necessity Supplies Ltd) (product) |
|  | 17592711000001100 | Avandamet 2mg/1000mg tablets (Necessity Supplies Ltd) (product) |
|  | 17593011000001100 | Avandamet 2mg/500mg tablets (Necessity Supplies Ltd) (product) |
|  | 17593811000001100 | Avandamet 4mg/1000mg tablets (Necessity Supplies Ltd) (product) |
|  | 17594311000001100 | Avandia 4mg tablets (Necessity Supplies Ltd) (product) |
|  | 17601411000001100 | NovoMix 30 FlexPen 100units/ml suspension for injection 3ml pre-filled pens (Necessity Supplies Ltd) (product) |
|  | 17601611000001100 | NovoMix 30 Penfill 100units/ml suspension for injection 3ml cartridges (Necessity Supplies Ltd) (product) |
|  | 17601911000001100 | NovoRapid FlexPen 100units/ml solution for injection 3ml pre-filled pens (Necessity Supplies Ltd) (product) |
|  | 17609511000001100 | Humulin M3 KwikPen 100units/ml suspension for injection 3ml pre-filled pens (Eli Lilly and Company Ltd) (product) |
|  | 17621611000001100 | Prandin 0.5mg tablets (Necessity Supplies Ltd) (product) |
|  | 17821311000001100 | Tolbutamide 500mg tablets (Phoenix Healthcare Distribution Ltd) (product) |
|  | 17880511000001100 | Gliclazide 40mg tablets (A A H Pharmaceuticals Ltd) (product) |
|  | 17899911000001100 | Humulin M3 100units/ml suspension for injection 3ml cartridges (Sigma Pharmaceuticals Plc) (product) |
|  | 17900111000001100 | Onglyza 5mg tablets (Sigma Pharmaceuticals Plc) (product) |
|  | 17914711000001100 | Glibenclamide 2.5mg tablets (Phoenix Healthcare Distribution Ltd) (product) |
|  | 17915611000001100 | Gliclazide 80mg tablets (Phoenix Healthcare Distribution Ltd) (product) |
|  | 17918011000001100 | Glipizide 5mg tablets (Phoenix Healthcare Distribution Ltd) (product) |

|  | 17940011000001100 | Metformin 500mg tablets (Phoenix Healthcare Distribution Ltd) (product) |
| --- | --- | --- |
|  | 17970111000001100 | Metformin 500mg/5ml oral solution sugar free (Almus Pharmaceuticals Ltd) (product) |
|  | 17972811000001100 | Glibenclamide 2.5mg tablets (Almus Pharmaceuticals Ltd) (product) |
|  | 17973211000001100 | Glibenclamide 5mg tablets (Almus Pharmaceuticals Ltd) (product) |
|  | 18030311000001100 | Insuman Infusat 100units/ml solution for injection 10ml vials (Imported (Germany)) (product) |
|  | 18046311000001100 | Insuman Infusat 100units/ml solution for injection 3.15ml cartridges (Imported (Germany)) (product) |
|  | 18150311000001100 | Insuman Comb 25 100units/ml suspension for injection 3ml pre-filled SoloStar pens (Sanofi) (product) |
|  | 18155911000001100 | Starlix 120mg tablets (Mawdsley-Brooks & Company Ltd) (product) |
|  | 18212011000001100 | Lantus 100units/ml solution for injection 10ml vials (Mawdsley-Brooks & Company Ltd) (product) |
|  | 18224311000001100 | Competact 15mg/850mg tablets (Necessity Supplies Ltd) (product) |
|  | 18265111000001100 | Glucobay 100mg tablets (Necessity Supplies Ltd) (product) |
|  | 18309811000001100 | Acarbose 100mg tablets (A A H Pharmaceuticals Ltd) (product) |
|  | 18362011000001100 | Metformin 500mg modified-release tablets (Mawdsley-Brooks & Company Ltd) (product) |
|  | 18462111000001100 | Gliclazide 80mg tablets (Accord Healthcare Ltd) (product) |
|  | 18462611000001100 | Glimepiride 3mg tablets (Accord Healthcare Ltd) (product) |
|  | 18462811000001100 | Glimepiride 4mg tablets (Accord Healthcare Ltd) (product) |
|  | 184711000001100 | Gliclazide 80mg tablets (Sterwin Medicines) (product) |
|  | 18490911000001100 | Lantus 100units/ml solution for injection 3ml pre-filled OptiSet pens (Necessity Supplies Ltd) (product) |
|  | 18567011000001100 | Pioglitazone 45mg/5ml oral suspension (Special Order) (product) |
|  | 18580011000001100 | Glucophage SR 500mg tablets (Doncaster Pharmaceuticals Ltd) (product) |
|  | 18595311000001100 | Pioglitazone 45mg/5ml oral suspension (product) |
|  | 18624811000001100 | Acarbose 50mg tablets (Phoenix Healthcare Distribution Ltd) (product) |
|  | 18631911000001100 | Onglyza 5mg tablets (Mawdsley-Brooks & Company Ltd) (product) |
|  | 18678711000001100 | Vitile XL 30mg tablets (Actavis UK Ltd) (product) |
|  | 18745311000001100 | Humalog KwikPen 100units/ml solution for injection 3ml pre-filled pens (Sigma Pharmaceuticals Plc) (product) |
|  | 18745511000001100 | Humalog Mix50 KwikPen 100units/ml suspension for injection 3ml pre-filled pens (Sigma Pharmaceuticals Plc) (product) |
|  | 19274811000001100 | Bydureon 2mg powder and solvent for suspension for injection vials (Eli Lilly and Company Ltd) (product) |
|  | 19275411000001100 | Exenatide 2mg powder and solvent for suspension for injection vials (product) |
|  | 19276911000001100 | Repaglinide 1mg tablets (Aspire Pharma Ltd) (product) |
|  | 19277111000001100 | Repaglinide 500microgram tablets (Aspire Pharma Ltd) (product) |
|  | 19300511000001100 | Repaglinide 1mg tablets (Actavis UK Ltd) (product) |
|  | 19301211000001100 | Metabet SR 1000mg tablets (Actavis UK Ltd) (product) |
|  | 19306911000001100 | Enyglid 1mg tablets (Consilient Health Ltd) (product) |
|  | 19307311000001100 | Enyglid 2mg tablets (Consilient Health Ltd) (product) |
|  | 19354411000001100 | Insuman Basal 100units/ml suspension for injection 3ml pre-filled SoloStar pens (Sanofi) (product) |
|  | 19361111000001100 | Repaglinide 2mg tablets (Alliance Healthcare (Distribution) Ltd) (product) |
|  | 19361311000001100 | Pioglitazone 15mg tablets (A A H Pharmaceuticals Ltd) (product) |
|  | 19361711000001100 | Repaglinide 500microgram tablets (A A H Pharmaceuticals Ltd) (product) |
|  | 19362011000001100 | Repaglinide 1mg tablets (A A H Pharmaceuticals Ltd) (product) |
|  | 19362311000001100 | Repaglinide 2mg tablets (A A H Pharmaceuticals Ltd) (product) |
|  | 19469611000001100 | Pioglitazone 45mg tablets (Alliance Healthcare (Distribution) Ltd) (product) |
|  | 19473511000001100 | Pioglitazone 45mg tablets (A A H Pharmaceuticals Ltd) (product) |
|  | 19476411000001100 | Pioglitazone 30mg tablets (Consilient Health Ltd) (product) |
|  | 19476611000001100 | Pioglitazone 45mg tablets (Consilient Health Ltd) (product) |
|  | 19489511000001100 | Acarbose 100mg tablets (Alliance Healthcare (Distribution) Ltd) (product) |
|  | 19527011000001100 | Actos 45mg tablets (Waymade Healthcare Plc) (product) |
|  | 19570211000001100 | NovoRapid FlexTouch 100units/ml solution for injection 3ml pre-filled pens (Novo Nordisk Ltd) (product) |
|  | 19593311000001100 | Pioglitazone 45mg tablets (Dr Reddy's Laboratories (UK) Ltd) (product) |
|  | 19611511000001100 | Eucreas 50mg/850mg tablets (Doncaster Pharmaceuticals Ltd) (product) |
|  | 19613511000001100 | Humalog Mix50 KwikPen 100units/ml suspension for injection 3ml pre-filled pens (DE Pharmaceuticals) (product) |
|  | 19614111000001100 | Humalog 100units/ml solution for injection 10ml vials (Doncaster Pharmaceuticals Ltd) (product) |
|  | 19726511000001100 | Actos 45mg tablets (Mawdsley-Brooks & Company Ltd) (product) |
|  | 197511000001103 | Metformin 500mg tablets (Unichem Plc) (product) |
|  | 19818611000001100 | Glipizide 5mg tablets (Almus Pharmaceuticals Ltd) (product) |
|  | 19863911000001100 | Actos 45mg tablets (Lexon (UK) Ltd) (product) |
|  | 19957811000001100 | Glizofar 15mg tablets (Teva UK Ltd) (product) |
|  | 20022911000001100 | Glizofar 15mg tablets (Arrow Generics Ltd) (product) |
|  | 20023111000001100 | Glizofar 30mg tablets (Arrow Generics Ltd) (product) |
|  | 20023411000001100 | Glizofar 45mg tablets (Arrow Generics Ltd) (product) |
|  | 20114811000001100 | Januvia 50mg Tablets (Merck Sharp & Dohme Ltd) (product) |
|  | 20163611000001100 | NovoRapid Penfill 100units/ml solution for injection 3ml cartridges (Doncaster Pharmaceuticals Ltd) (product) |
|  | 20319611000001100 | Pioglitazone 15mg tablets (Sandoz Ltd) (product) |
|  | 20320211000001100 | Pioglitazone 30mg tablets (Sandoz Ltd) (product) |
|  | 20320611000001100 | Pioglitazone 45mg tablets (Sandoz Ltd) (product) |
|  | 20357611000001100 | Pioglitazone 30mg tablets (Actavis UK Ltd) (product) |
|  | 20566211000001100 | Repaglinide 500microgram tablets (Creo Pharma Ltd) (product) |
|  | 20566811000001100 | Repaglinide 2mg tablets (Creo Pharma Ltd) (product) |
|  | 20773611000001100 | Repaglinide 500microgram tablets (Accord Healthcare Ltd) (product) |
|  | 20778911000001100 | Repaglinide 2mg tablets (Accord Healthcare Ltd) (product) |
|  | 20925511000001100 | Onglyza 5mg tablets (Waymade Healthcare Plc) (product) |
|  | 21028111000001100 | Acarbose 100mg tablets (Arrow Generics Ltd) (product) |
|  | 21111411000001100 | Acarbose 50mg tablets (Actavis UK Ltd) (product) |
|  | 21208211000001100 | Jentadueto 2.5mg/850mg tablets (Boehringer Ingelheim Ltd) (product) |
|  | 21208511000001100 | Jentadueto 2.5mg/1000mg tablets (Boehringer Ingelheim Ltd) (product) |
|  | 21245011000001100 | Linagliptin 2.5mg / Metformin 1g tablets (product) |
|  | 21245111000001100 | Linagliptin 2.5mg / Metformin 850mg tablets (product) |
|  | 21366911000001100 | Glimepiride 6mg/5ml oral suspension (product) |
|  | 21409211000001100 | Gliclazide 80mg tablets (Medreich Plc) (product) |
|  | 21409811000001100 | Metabet SR 500mg tablets (Actavis UK Ltd) (product) |
|  | 21501311000001100 | Glucophage SR 1000mg tablets (Waymade Healthcare Plc) (product) |
|  | 21705311000001100 | Komboglyze 2.5mg/850mg tablets (Bristol-Myers Squibb Pharmaceuticals Ltd) (product) |
|  | 21705611000001100 | Komboglyze 2.5mg/1000mg tablets (Bristol-Myers Squibb Pharmaceuticals Ltd) (product) |
|  | 21711411000001100 | Saxagliptin 2.5mg / Metformin 1g tablets (product) |
|  | 21711511000001100 | Saxagliptin 2.5mg / Metformin 850mg tablets (product) |
|  | 21735211000001100 | Acarbose 100mg tablets (Waymade Healthcare Plc) (product) |
|  | 21763311000001100 | Pioglitazone 15mg tablets (Teva UK Ltd) (product) |
|  | 21764211000001100 | Pioglitazone 30mg tablets (Teva UK Ltd) (product) |
|  | 21764511000001100 | Pioglitazone 45mg tablets (Teva UK Ltd) (product) |
|  | 21793311000001100 | Metformin 1g modified-release tablets (Waymade Healthcare Plc) (product) |
|  | 21793711000001100 | Metformin 500mg tablets (Waymade Healthcare Plc) (product) |
|  | 21794511000001100 | Metformin 850mg tablets (Waymade Healthcare Plc) (product) |
|  | 21848111000001100 | Pioglitazone 15mg tablets (Waymade Healthcare Plc) (product) |
|  | 21848311000001100 | Pioglitazone 30mg tablets (Waymade Healthcare Plc) (product) |
|  | 21848511000001100 | Pioglitazone 45mg tablets (Waymade Healthcare Plc) (product) |
|  | 21880011000001100 | Repaglinide 500microgram tablets (Waymade Healthcare Plc) (product) |
|  | 21880611000001100 | Repaglinide 2mg tablets (Waymade Healthcare Plc) (product) |
|  | 21930011000001100 | Tresiba FlexTouch 100units/ml solution for injection 3ml pre-filled pens (Novo Nordisk Ltd) (product) |
|  | 21931911000001100 | Tresiba FlexTouch 200units/ml solution for injection 3ml pre-filled pens (Novo Nordisk Ltd) (product) |
|  | 21939611000001100 | Insulin degludec 100units/ml solution for injection 3ml pre-filled disposable devices (product) |
|  | 21939711000001100 | Insulin degludec 200units/ml solution for injection 3ml pre-filled disposable devices (product) |
|  | 21941511000001100 | Lyxumia 10micrograms/0.2ml solution for injection 3ml pre-filled pens (Sanofi) (product) |
|  | 21994811000001100 | Lixisenatide 20micrograms/0.2ml solution for injection 3ml pre-filled disposable devices (product) |
|  | 22026011000001100 | Glibenclamide 2.5mg tablets (Waymade Healthcare Plc) (product) |
|  | 22026511000001100 | Gliclazide 40mg tablets (Waymade Healthcare Plc) (product) |
|  | 22026711000001100 | Gliclazide 80mg tablets (Waymade Healthcare Plc) (product) |
|  | 22027911000001100 | Glimepiride 3mg tablets (Waymade Healthcare Plc) (product) |
|  | 22082511000001100 | Gliclazide 30mg modified-release tablets (Waymade Healthcare Plc) (product) |
|  | 22308811000001100 | Bolamyn SR 1000mg tablets (Teva UK Ltd) (product) |
|  | 22350011000001100 | Metformin 850mg tablets (Aurobindo Pharma Ltd) (product) |
|  | 22643511000001100 | Lantus 100units/ml solution for injection 3ml OptiClik cartridges (Waymade Healthcare Plc) (product) |
|  | 228111000001101 | Amaryl 1mg tablets (Aventis Pharma) (product) |
|  | 23369611000001100 | Glidipion 30mg tablets (Actavis UK Ltd) (product) |
|  | 23487411000001100 | Pioglitazone 15mg tablets (Accord Healthcare Ltd) (product) |
|  | 23487811000001100 | Pioglitazone 45mg tablets (Accord Healthcare Ltd) (product) |
|  | 23637311000001100 | Alogliptin 12.5mg tablets (product) |
|  | 23677911000001100 | Sitagliptin 50mg tablets (Waymade Healthcare Plc) (product) |
|  | 23678111000001100 | Sitagliptin 100mg tablets (Waymade Healthcare Plc) (product) |
|  | 23943211000001100 | Glibenclamide 2.5mg tablets (Doncaster Pharmaceuticals Ltd) (product) |
|  | 23943611000001100 | Gliclazide 80mg tablets (Doncaster Pharmaceuticals Ltd) (product) |
|  | 24018111000001100 | Xigduo 5mg/850mg tablets (Bristol-Myers Squibb Pharmaceuticals Ltd) (product) |
|  | 24018511000001100 | Xigduo 5mg/1000mg tablets (Bristol-Myers Squibb Pharmaceuticals Ltd) (product) |
|  | 24054711000001100 | Dapagliflozin 5mg / Metformin 850mg tablets (product) |
|  | 24088311000001100 | Invokana 300mg tablets (Janssen-Cilag Ltd) (product) |
|  | 24088611000001100 | Invokana 100mg tablets (Janssen-Cilag Ltd) (product) |
|  | 24106711000001100 | Glimepiride 3mg tablets (DE Pharmaceuticals) (product) |
|  | 24106911000001100 | Glimepiride 4mg tablets (DE Pharmaceuticals) (product) |
|  | 24134911000001100 | Pioglitazone 30mg tablets (Morningside Healthcare Ltd) (product) |
|  | 24137211000001100 | Pioglitazone 15mg tablets (Morningside Healthcare Ltd) (product) |
|  | 24380811000001100 | Metformin 1g modified-release tablets (DE Pharmaceuticals) (product) |
|  | 24381111000001100 | Metformin 500mg modified-release tablets (DE Pharmaceuticals) (product) |
|  | 24554511000001100 | Metformin 1g modified-release tablets (Actavis UK Ltd) (product) |
|  | 24568411000001100 | Sukkarto SR 500mg tablets (Morningside Healthcare Ltd) (product) |
|  | 25238811000001100 | Jardiance 10mg tablets (Boehringer Ingelheim Ltd) (product) |
|  | 25239711000001100 | Jardiance 25mg tablets (Boehringer Ingelheim Ltd) (product) |
|  | 25290511000001100 | Empagliflozin 10mg tablets (product) |
|  | 27117211000001100 | Tolbutamide 500mg tablets (Genesis Pharmaceuticals Ltd) (product) |
|  | 27879911000001100 | Metformin 500mg/5ml oral solution sugar free (Pinewood Healthcare) (product) |
|  | 27957011000001100 | Ziclaseg 30mg modified-release tablets (Lupin (Europe) Ltd) (product) |
|  | 28022511000001100 | Vokanamet 50mg/850mg tablets (Janssen-Cilag Ltd) (product) |
|  | 28024411000001100 | Vokanamet 50mg/1000mg tablets (Janssen-Cilag Ltd) (product) |
|  | 28049211000001100 | Canagliflozin 50mg / Metformin 1g tablets (product) |
|  | 28049311000001100 | Canagliflozin 50mg / Metformin 850mg tablets (product) |
|  | 28279611000001100 | Insulin degludec 100units/ml / Liraglutide 3.6mg/ml solution for injection 3ml pre-filled disposable devices (product) |
|  | 28420911000001100 | Vamju 60mg modified-release tablets (AMCo) (product) |
|  | 28425111000001100 | Forxiga 5mg tablets (Waymade Healthcare Plc) (product) |
|  | 28440211000001100 | Exenatide 2mg powder and solvent for suspension for injection pre-filled disposable devices (product) |
|  | 28462311000001100 | Trulicity 1.5mg/0.5ml solution for injection pre-filled pens (Eli Lilly and Company Ltd) (product) |
|  | 28775411000001100 | Canagliflozin 300mg tablets (Colorama Pharmaceuticals Ltd) (product) |
|  | 287811000001101 | Diabetamide 5mg tablets (Ashbourne Pharmaceuticals Ltd) (product) |
|  | 28789711000001100 | Dulaglutide 1.5mg/0.5ml solution for injection pre-filled disposable devices (product) |
|  | 28926811000001100 | Humalog KwikPen 200units/ml solution for injection 3ml pre-filled pens (Eli Lilly and Company Ltd) (product) |
|  | 28940811000001100 | Glidipion 15mg tablets (Actavis UK Ltd) (product) |
|  | 29700111000001100 | Liraglutide 6mg/ml solution for injection 3ml pre-filled disposable devices (J M McGill Ltd) (product) |
|  | 29737811000001100 | Onglyza 2.5mg tablets (Waymade Healthcare Plc) (product) |
|  | 29743011000001100 | Diabiom 30mg tablets (Tillomed Laboratories Ltd) (product) |
|  | 29743211000001100 | Diabiom 45mg tablets (Tillomed Laboratories Ltd) (product) |
|  | 29750011000001100 | Sitagliptin 25mg tablets (Waymade Healthcare Plc) (product) |
|  | 29754711000001100 | Acarbose 50mg tablets (Sigma Pharmaceuticals Plc) (product) |
|  | 29853611000001100 | Canagliflozin 100mg tablets (Niche Pharma Ltd) (product) |
|  | 29855111000001100 | Metformin 1g oral powder sachets sugar free (Niche Pharma Ltd) (product) |
|  | 29866811000001100 | Toujeo 300units/ml solution for injection 1.5ml pre-filled SoloStar pens (Sanofi) (product) |
|  | 29869711000001100 | Gliclazide 80mg tablets (Sigma Pharmaceuticals Plc) (product) |
|  | 29870011000001100 | Glipizide 5mg tablets (Sigma Pharmaceuticals Plc) (product) |
|  | 29903611000001100 | Insulin glargine 300units/ml solution for injection 1.5ml pre-filled disposable devices (product) |
|  | 29906011000001100 | Metformin 500mg tablets (Sigma Pharmaceuticals Plc) (product) |
|  | 29980811000001100 | Byetta 10micrograms/0.04ml solution for injection 2.4ml pre-filled pens (Lexon (UK) Ltd) (product) |
|  | 29981611000001100 | Bydureon 2mg powder and solvent for suspension for injection vials (Lexon (UK) Ltd) (product) |
|  | 29986211000001100 | Pioglitazone 45mg tablets (Sigma Pharmaceuticals Plc) (product) |
|  | 29988711000001100 | Acarbose 50mg tablets (Mawdsley-Brooks & Company Ltd) (product) |
|  | 29988911000001100 | Acarbose 100mg tablets (Mawdsley-Brooks & Company Ltd) (product) |
|  | 30056811000001100 | Gliclazide 30mg modified-release tablets (DE Pharmaceuticals) (product) |
|  | 30088811000001100 | Repaglinide 1mg tablets (DE Pharmaceuticals) (product) |
|  | 30089311000001100 | Repaglinide 2mg tablets (DE Pharmaceuticals) (product) |
|  | 30112311000001100 | Pioglitazone 45mg tablets (DE Pharmaceuticals) (product) |
|  | 30134911000001100 | Gliclazide 80mg tablets (Mawdsley-Brooks & Company Ltd) (product) |
|  | 30135211000001100 | Gliclazide 30mg modified-release tablets (Mawdsley-Brooks & Company Ltd) (product) |
|  | 30171811000001100 | Abasaglar KwikPen 100units/ml solution for injection 3ml pre-filled pens (Eli Lilly and Company Ltd) (product) |
|  | 30172211000001100 | Abasaglar 100units/ml solution for injection 3ml cartridges (Eli Lilly and Company Ltd) (product) |
|  | 30174111000001100 | Synjardy 5mg/1000mg tablets (Boehringer Ingelheim Ltd) (product) |
|  | 30175011000001100 | Synjardy 12.5mg/850mg tablets (Boehringer Ingelheim Ltd) (product) |
|  | 30175711000001100 | Synjardy 12.5mg/1000mg tablets (Boehringer Ingelheim Ltd) (product) |
|  | 30204011000001100 | Glipizide 5mg tablets (Mawdsley-Brooks & Company Ltd) (product) |
|  | 30208211000001100 | Eucreas 50mg/1000mg tablets (Waymade Healthcare Plc) (product) |
|  | 30214411000001100 | Vipdomet 12.5mg/1000mg tablets (Waymade Healthcare Plc) (product) |
|  | 30214611000001100 | Vipidia 12.5mg tablets (Waymade Healthcare Plc) (product) |
|  | 30269611000001100 | Byetta 5micrograms/0.02ml solution for injection 1.2ml pre-filled pens (Waymade Healthcare Plc) (product) |
|  | 30318211000001100 | Empagliflozin 12.5mg / Metformin 850mg tablets (product) |
|  | 30318311000001100 | Empagliflozin 5mg / Metformin 1g tablets (product) |
|  | 30318411000001100 | Empagliflozin 5mg / Metformin 850mg tablets (product) |
|  | 30763311000001100 | Metformin 500mg modified-release tablets (Almus Pharmaceuticals Ltd) (product) |
|  | 30828011000001100 | Metformin 1g modified-release tablets (Mawdsley-Brooks & Company Ltd) (product) |
|  | 30857211000001100 | Pioglitazone 30mg tablets (Mawdsley-Brooks & Company Ltd) (product) |
|  | 30857411000001100 | Pioglitazone 45mg tablets (Mawdsley-Brooks & Company Ltd) (product) |
|  | 30982411000001100 | Bilxona 30mg modified-release tablets (Accord Healthcare Ltd) (product) |
|  | 30989911000001100 | Metformin 500mg modified-release tablets (Alliance Healthcare (Distribution) Ltd) (product) |
|  | 30992711000001100 | Insulin soluble human 1unit/ml solution for injection 50ml pre-filled syringes (product) |
|  | 30993011000001100 | Insulin soluble human 1unit/ml solution for injection 50ml pre-filled syringes (Special Order) (product) |
|  | 31351711000001100 | Alogliptin 12.5mg / Metformin 1g tablets (J M McGill Ltd) (product) |
|  | 31352111000001100 | Alogliptin 25mg tablets (J M McGill Ltd) (product) |

|  | 32423011000001100 | Metformin 1g modified-release tablets (Kent Pharmaceuticals Ltd) (product) |
| --- | --- | --- |
|  | 32432111000001100 | Alogliptin 25mg tablets (Colorama Pharmaceuticals Ltd) (product) |
|  | 32493411000001100 | Glimepiride 1mg tablets (Brown & Burk UK Ltd) (product) |
|  | 32493611000001100 | Glimepiride 2mg tablets (Brown & Burk UK Ltd) (product) |
|  | 32494011000001100 | Glimepiride 4mg tablets (Brown & Burk UK Ltd) (product) |
|  | 32498311000001100 | Gliclazide 40mg tablets (Actavis UK Ltd) (product) |
|  | 325011000001101 | Metformin 850mg tablets (A A H Pharmaceuticals Ltd) (product) |
|  | 325219009 | Product containing precisely glibenclamide 5 milligram/1 each conventional release oral tablet (clinical drug) |
|  | 325251005 | Product containing precisely gliquidone 30 milligram/1 each conventional release oral tablet (clinical drug) |
|  | 325259007 | Product containing precisely glimepiride 2 milligram/1 each conventional release oral tablet (clinical drug) |
|  | 325261003 | Product containing precisely glimepiride 1 milligram/1 each conventional release oral tablet (clinical drug) |
|  | 325267004 | Product containing precisely tolbutamide 500 milligram/1 each conventional release oral tablet (clinical drug) |
|  | 325278007 | Product containing precisely metformin hydrochloride 500 milligram/1 each conventional release oral tablet (clinical drug) |
|  | 3256111000001100 | Insulatard ge 100units/ml suspension for injection 10ml vials (Novo Nordisk Pharmaceuticals Ltd) (product) |
|  | 3259411000001100 | Human Insulatard Penfill 100units/ml suspension for injection 1.5ml cartridges (Novo Nordisk Pharmaceuticals Ltd) (product) |
|  | 3259811000001100 | Insuman Comb 25 100units/ml suspension for injection 3ml pre-filled OptiSet pens (Sanofi) (product) |
|  | 326038002 | Product containing precisely acarbose 100 milligram/1 each conventional release oral tablet (clinical drug) |
|  | 326047005 | Product containing precisely repaglinide 500 microgram/1 each conventional release oral tablet (clinical drug) |
|  | 326061000 | Product containing precisely pioglitazone (as pioglitazone hydrochloride) 30 milligram/1 each conventional release oral tablet (clinical drug) |
|  | 3260811000001100 | Humulin I Pen 100units/ml suspension for injection 3ml pre-filled pens (Eli Lilly and Company Ltd) (product) |
|  | 3261411000001100 | Insulatard InnoLet 100units/ml suspension for injection 3ml pre-filled pens (Novo Nordisk Ltd) (product) |
|  | 3263011000001100 | Insuman Comb 25 100units/ml suspension for injection 5ml vials (Aventis Pharma) (product) |
|  | 3263611000001100 | Humulin M2 100units/ml suspension for injection 3ml cartridges (Eli Lilly & Co Ltd) (product) |
|  | 32638311000001100 | Sitagliptin 50mg/5ml oral solution (product) |
|  | 3264211000001100 | Humulin Isophane 100units/ml suspension for injection 3ml cartridges (Eli Lilly & Co Ltd) (product) |
|  | 3264411000001100 | Mixtard 20 Penfill 100units/ml suspension for injection 3ml cartridges (Novo Nordisk Pharmaceuticals Ltd) (product) |
|  | 3268711000001100 | Human Mixtard 10 Penfill 100units/ml suspension for injection 1.5ml cartridges (Novo Nordisk Pharmaceuticals Ltd) (product) |
|  | 3269711000001100 | Mixtard 10 NovoLet 100units/ml suspension for injection (Novo Nordisk Pharmaceuticals Ltd) (product) |
|  | 3269911000001100 | Hypurin Bovine Protamine Zinc 100units/ml suspension for injection 10ml vials (C P Pharmaceuticals Ltd) (product) |
|  | 3270211000001100 | Insuman Comb 15 100units/ml suspension for injection 3ml pre-filled OptiSet pens (Sanofi) (product) |
|  | 3270611000001100 | Human Mixtard 50 100units/ml suspension for injection 10ml vials (Novo Nordisk Pharmaceuticals Ltd) (product) |
|  | 3271311000001100 | Hypurin Bovine Neutral 100units/ml solution for injection 10ml vials (C P Pharmaceuticals Ltd) (product) |
|  | 3271611000001100 | Mixtard 30 InnoLet 100units/ml suspension for injection 3ml pre-filled pens (Novo Nordisk Ltd) (product) |
|  | 3271711000001100 | Insuman Comb 15 100units/ml suspension for injection 5ml vials (Aventis Pharma) (product) |
|  | 3272211000001100 | Humaject M3 Pen 100units/ml suspension for injection (Eli Lilly & Co Ltd) (product) |
|  | 3272411000001100 | Insuman Comb 50 100units/ml suspension for injection 3ml cartridges (Aventis Pharma) (product) |
|  | 3273111000001100 | Mixtard 30 Penfill 100units/ml suspension for injection 3ml cartridges (Novo Nordisk Pharmaceuticals Ltd) (product) |
|  | 3273411000001100 | Human Mixtard 50 Penfill 100units/ml suspension for injection 1.5ml cartridges (Novo Nordisk Pharmaceuticals Ltd) (product) |
|  | 3273911000001100 | Insuman Comb 15 100units/ml suspension for injection 3ml cartridges (Aventis Pharma) (product) |
|  | 3274011000001100 | Hypurin Bovine Neutral 100units/ml solution for injection 1.5ml cartridges (C P Pharmaceuticals Ltd) (product) |
|  | 3274811000001100 | Humulin M3 100units/ml suspension for injection 10ml vials (Eli Lilly & Co Ltd) (product) |
|  | 3275311000001100 | Human Mixtard 30 Penfill 100units/ml suspension for injection 1.5ml cartridges (Novo Nordisk Pharmaceuticals Ltd) (product) |
|  | 3275711000001100 | Humalog Mix25 100units/ml suspension for injection 3ml cartridges (Eli Lilly & Co Ltd) (product) |
|  | 3276011000001100 | Humalog Mix25 Pen 100units/ml suspension for injection 3ml pre-filled pens (Eli Lilly and Company Ltd) (product) |
|  | 3277811000001100 | NovoMix 30 Penfill 100units/ml suspension for injection 3ml cartridges (Novo Nordisk Pharmaceuticals Ltd) (product) |
|  | 3278611000001100 | Insuman Comb 50 100units/ml suspension for injection 3ml pre-filled OptiSet pens (Sanofi) (product) |
|  | 3279211000001100 | NovoRapid Penfill 100units/ml solution for injection 3ml cartridges (Novo Nordisk Pharmaceuticals Ltd) (product) |
|  | 3280611000001100 | Hypurin Bovine Lente 100units/ml suspension for injection 10ml vials (C P Pharmaceuticals Ltd) (product) |
|  | 3280711000001100 | NovoRapid 100units/ml solution for injection 10ml vials (Novo Nordisk Pharmaceuticals Ltd) (product) |
|  | 3281811000001100 | Humulin Zn 100units/ml suspension for injection 10ml vials (Eli Lilly & Co Ltd) (product) |
|  | 3282211000001100 | NovoRapid FlexPen 100units/ml solution for injection 3ml pre-filled pens (Novo Nordisk Ltd) (product) |
|  | 3282311000001100 | Ultratard 100units/ml suspension for injection 10ml vials (Novo Nordisk Pharmaceuticals Ltd) (product) |
|  | 3282611000001100 | Hypurin Porcine Isophane 100units/ml suspension for injection 1.5ml cartridges (C P Pharmaceuticals Ltd) (product) |
|  | 3284011000001100 | Hypurin Porcine Isophane 100units/ml suspension for injection 10ml vials (C P Pharmaceuticals Ltd) (product) |
|  | 3284111000001100 | Monotard 100units/ml suspension for injection 10ml vials (Novo Nordisk Pharmaceuticals Ltd) (product) |
|  | 3284211000001100 | Lantus 100units/ml solution for injection 3ml cartridges (Aventis Pharma) (product) |
|  | 3284311000001100 | Humalog 100units/ml solution for injection 3ml cartridges (Eli Lilly & Co Ltd) (product) |
|  | 3285511000001100 | Human Mixtard 40 Penfill 100units/ml suspension for injection 1.5ml cartridges (Novo Nordisk Pharmaceuticals Ltd) (product) |
|  | 3285611000001100 | Pork Actrapid 100units/ml solution for injection 10ml vials (Novo Nordisk Pharmaceuticals Ltd) (product) |
|  | 3287911000001100 | Lantus 100units/ml solution for injection 10ml vials (Aventis Pharma) (product) |
|  | 3291711000001100 | Insuman Rapid 100units/ml solution for injection 3ml pre-filled OptiSet pens (Sanofi) (product) |
|  | 3311311000001100 | Humulin S 100units/ml solution for injection 3ml cartridges (Eli Lilly & Co Ltd) (product) |
|  | 3311611000001100 | Insuman Rapid 100units/ml solution for injection 3ml cartridges (Aventis Pharma) (product) |
|  | 3312111000001100 | Actrapid 100units/ml solution for injection 10ml vials (Novo Nordisk Pharmaceuticals Ltd) (product) |
|  | 3312411000001100 | Velosulin 100units/ml solution for injection 10ml vials (Novo Nordisk Pharmaceuticals Ltd) (product) |
|  | 3333111000001100 | Pork Insulatard 100units/ml suspension for injection 10ml vials (Novo Nordisk Pharmaceuticals Ltd) (product) |
|  | 33425611000001100 | Metformin 750mg modified-release tablets (A A H Pharmaceuticals Ltd) (product) |
|  | 33428211000001100 | Gliclazide 40mg tablets (Alliance Healthcare (Distribution) Ltd) (product) |
|  | 33548011000001100 | Metformin 1g/5ml oral solution sugar free (Colonis Pharma Ltd) (product) |
|  | 33550911000001100 | Metformin 850mg/5ml oral solution sugar free (product) |
|  | 33598711000001100 | Pioglitazone 45mg tablets (Mylan Ltd) (product) |
|  | 33598911000001100 | Pioglitazone 30mg tablets (Mylan Ltd) (product) |
|  | 33613711000001100 | Repaglinide 500microgram tablets (Mylan Ltd) (product) |
|  | 33614311000001100 | Repaglinide 1mg tablets (Mylan Ltd) (product) |
|  | 33747711000001100 | Saxenda 6mg/ml solution for injection 3ml pre-filled pens (Novo Nordisk Ltd) (product) |
|  | 33766211000001100 | Zicron PR 30mg tablets (Bristol Laboratories Ltd) (product) |
|  | 33769011000001100 | Metformin 850mg/5ml oral solution sugar free (Alliance Healthcare (Distribution) Ltd) (product) |
|  | 33769211000001100 | Metformin 1g/5ml oral solution sugar free (Alliance Healthcare (Distribution) Ltd) (product) |
|  | 3468711000001100 | Insulin aspart 100units/ml solution for injection 3ml pre-filled disposable devices (product) |
|  | 3468811000001100 | Insulin aspart biphasic 30/70 100units/ml suspension for injection 3ml cartridges (product) |
|  | 3468911000001100 | Insulin aspart biphasic 30/70 100units/ml suspension for injection 3ml pre-filled disposable devices (product) |
|  | 3469311000001100 | Insulin isophane biphasic human 10/90 100units/ml suspension for injection 1.5ml cartridges (product) |
|  | 3469411000001100 | Insulin isophane biphasic human 10/90 100units/ml suspension for injection 3ml cartridges (product) |
|  | 3469511000001100 | Insulin isophane biphasic human 10/90 100units/ml suspension for injection 3ml pre-filled disposable devices (product) |
|  | 3469611000001100 | Insulin isophane biphasic human 15/85 100units/ml suspension for injection 3ml cartridges (product) |
|  | 3469711000001100 | Insulin isophane biphasic human 15/85 100units/ml suspension for injection 3ml pre-filled disposable devices (product) |
|  | 3469911000001100 | Insulin isophane biphasic human 20/80 100units/ml suspension for injection 1.5ml cartridges (product) |
|  | 3470011000001100 | Insulin isophane biphasic human 20/80 100units/ml suspension for injection 3ml cartridges (product) |
|  | 3470311000001100 | Insulin isophane biphasic human 25/75 100units/ml suspension for injection 3ml pre-filled disposable devices (product) |
|  | 3470411000001100 | Insulin isophane biphasic human 25/75 100units/ml suspension for injection 5ml vials (product) |
|  | 3470511000001100 | Insulin isophane biphasic human 30/70 100units/ml suspension for injection 1.5ml cartridges (product) |
|  | 3470911000001100 | Insulin isophane biphasic human 30/70 100units/ml suspension for injection 3ml cartridges (product) |
|  | 3471111000001100 | Insulin isophane biphasic human 40/60 100units/ml suspension for injection 1.5ml cartridges (product) |
|  | 3471211000001100 | Insulin isophane biphasic human 40/60 100units/ml suspension for injection 3ml cartridges (product) |
|  | 3471411000001100 | Insulin isophane biphasic human 50/50 100units/ml suspension for injection 1.5ml cartridges (product) |
|  | 3471511000001100 | Insulin isophane biphasic human 50/50 100units/ml suspension for injection 10ml vials (product) |
|  | 3471611000001100 | Insulin isophane biphasic human 50/50 100units/ml suspension for injection 3ml cartridges (product) |
|  | 3471811000001100 | Insulin isophane biphasic human 50/50 100units/ml suspension for injection 5ml vials (product) |
|  | 3471911000001100 | Insulin isophane biphasic porcine 30/70 100units/ml suspension for injection 1.5ml cartridges (product) |
|  | 3472911000001100 | Insulin isophane porcine 100units/ml suspension for injection 1.5ml cartridges (product) |
|  | 3473511000001100 | Insulin lispro biphasic 25/75 100units/ml suspension for injection 3ml pre-filled disposable devices (product) |
|  | 3473611000001100 | Insulin lispro biphasic 50/50 100units/ml suspension for injection 3ml pre-filled disposable devices (product) |
|  | 3473711000001100 | Insulin lispro biphasic 25/75 100units/ml suspension for injection 3ml cartridges (product) |
|  | 3474911000001100 | Insulin zinc mixed bovine 100units/ml suspension for injection 10ml vials (product) |
|  | 3650611000001100 | Starlix 60mg tablets (Novartis Pharmaceuticals UK Ltd) (product) |
|  | 3661311000001100 | Diamicron 30mg MR tablets (Servier Laboratories Limited) (product) |
|  | 374897009 | Product containing precisely pioglitazone (as pioglitazone hydrochloride) 45 milligram/1 each conventional release oral tablet (clinical drug) |
|  | 386047000 | Metformin hydrochloride 500mg m/r tablet (product) |
|  | 400780006 | Insulin glargine 100units/mL injection solution 3mL cartridge (product) |
|  | 400877001 | Insulin glargine 100units/mL injection solution 3mL prefilled disposable injection device (product) |
|  | 4028811000001100 | Hypurin Bovine Neutral 100units/ml solution for injection 3ml cartridges (C P Pharmaceuticals Ltd) (product) |
|  | 4033111000001100 | Insulin isophane biphasic porcine 30/70 100units/ml suspension for injection 3ml cartridges (product) |
|  | 4034311000001100 | Hypurin Porcine Isophane 100units/ml suspension for injection 3ml cartridges (C P Pharmaceuticals Ltd) (product) |
|  | 4053611000001100 | Insulin isophane porcine 100units/ml suspension for injection 3ml cartridges (product) |
|  | 409122001 | Product containing precisely metformin hydrochloride 500 milligram and rosiglitazone (as rosiglitazone maleate) 2 milligram/1 each conventional release oral tablet (clinical drug) |
|  | 409124000 | Product containing precisely metformin hydrochloride 1 gram and rosiglitazone (as rosiglitazone maleate) 2 milligram/1 each conventional release oral tablet (clinical drug) |
|  | 409197000 | Product containing precisely metformin hydrochloride 100 milligram/1 milliliter conventional release oral solution (clinical drug) |
|  | 423962004 | Product containing precisely sitagliptin (as sitagliptin phosphate) 25 milligram/1 each conventional release oral tablet (clinical drug) |
|  | 424513004 | Product containing precisely sitagliptin (as sitagliptin phosphate) 50 milligram/1 each conventional release oral tablet (clinical drug) |
|  | 444311000001106 | Glucobay 100 tablets (Bayer Plc) (product) |
|  | 446711000001101 | Actos 30mg tablets (Takeda UK Ltd) (product) |
|  | 450011000001104 | Tolbutamide 500mg tablets (A A H Pharmaceuticals Ltd) (product) |
|  | 494111000001103 | NovoNorm 1mg tablets (Novo Nordisk Pharmaceuticals Ltd) (product) |
|  | 498911000001107 | Gliclazide 80mg tablets (Alpharma Limited) (product) |
|  | 515111000001108 | Glibenese 5mg tablets (Pfizer Ltd) (product) |
|  | 5199411000001100 | Actos 45mg tablets (Takeda UK Ltd) (product) |
|  | 5268911000001100 | Humalog Mix25 100units/ml suspension for injection 3ml cartridges (PI) (Waymade Ltd) (product) |
|  | 5270711000001100 | NovoRapid Novolet 100units/ml solution for injection (PI) (Waymade Ltd) (product) |
|  | 5283711000001100 | NovoMix 30 FlexPen 100units/ml suspension for injection 3ml pre-filled pens (Waymade Healthcare Plc) (product) |
|  | 5329011000001100 | Actos 15mg tablets (PI) (Waymade Ltd) (product) |
|  | 5330711000001100 | Amaryl 2mg tablets (PI) (Waymade Ltd) (product) |
|  | 5330811000001100 | Amaryl 3mg tablets (PI) (Waymade Ltd) (product) |
|  | 5335311000001100 | Amaryl 1mg tablets (PI) (Waymade Ltd) (product) |
|  | 5337611000001100 | Avandia 8mg tablets (PI) (Waymade Ltd) (product) |
|  | 535811000001108 | Glibenclamide 5mg tablets (Approved Prescription Services) (product) |
|  | 5372211000001100 | Glucobay 100 tablets (PI) (Waymade Ltd) (product) |
|  | 5396011000001100 | NovoNorm 1mg tablets (PI) (Waymade Ltd) (product) |
|  | 5414811000001100 | Starlix 120mg tablets (PI) (Waymade Ltd) (product) |
|  | 5440011000001100 | Glucobay 100 tablets (PI) (Dowelhurst Ltd) (product) |
|  | 5449611000001100 | Glimepiride 3mg tablets (PI) (Dowelhurst Ltd) (product) |
|  | 5456111000001100 | Glibenclamide 5mg tablets (PI) (Dowelhurst Ltd) (product) |
|  | 5465811000001100 | Acarbose 50mg tablets (PI) (Dowelhurst Ltd) (product) |
|  | 551111000001104 | Gliclazide 80mg tablets (Unichem Plc) (product) |
|  | 5519911000001100 | Amaryl 1mg tablets (PI) (Dowelhurst Ltd) (product) |
|  | 5520411000001100 | Amaryl 2mg tablets (PI) (Dowelhurst Ltd) (product) |
|  | 5521211000001100 | Amaryl 4mg tablets (PI) (Dowelhurst Ltd) (product) |
|  | 5532711000001100 | Daonil 5mg tablets (PI) (Dowelhurst Ltd) (product) |
|  | 5548011000001100 | Glucobay 50 tablets (PI) (Dowelhurst Ltd) (product) |
|  | 58011000001106 | Actos 15mg tablets (Takeda UK Ltd) (product) |
|  | 593911000001106 | Metformin 500mg tablets (Sandoz Ltd) (product) |
|  | 61911000001108 | Glimil 80mg tablets (Milpharm Ltd) (product) |
|  | 637811000001108 | Metformin 850mg tablets (Sterwin Medicines) (product) |
|  | 700011000001100 | Glibenclamide 2.5mg tablets (Alpharma Limited) (product) |
|  | 7034411000001100 | Metformin 850mg tablets (IVAX Pharmaceuticals UK Ltd) (product) |
|  | 703679006 | Product containing precisely dapagliflozin propanediol 5 milligram/1 each conventional release oral tablet (clinical drug) |
|  | 703680009 | Product containing precisely dapagliflozin propanediol 10 milligram/1 each conventional release oral tablet (clinical drug) |
|  | 718311000001102 | Glibenclamide 5mg tablets (Unichem Plc) (product) |
|  | 7463711000001100 | Metformin 850mg tablets (Ranbaxy (UK) Ltd) (product) |
|  | 7589411000001100 | Levemir FlexPen 100units/ml solution for injection 3ml pre-filled pens (Novo Nordisk Ltd) (product) |
|  | 781411000001104 | Glucobay 50 tablets (Bayer Plc) (product) |
|  | 785311000001108 | Metformin 500mg tablets (Sterwin Medicines) (product) |
|  | 8093711000001100 | Glimepiride 2mg tablets (Unichem Plc) (product) |
|  | 8093911000001100 | Glimepiride 3mg tablets (Unichem Plc) (product) |
|  | 811811000001103 | Glipizide 5mg tablets (Kent Pharmaceuticals Ltd) (product) |
|  | 8174611000001100 | Avandamet 4mg/1000mg tablets (GlaxoSmithKline) (product) |
|  | 840811000001100 | NovoNorm 2mg tablets (Novo Nordisk Pharmaceuticals Ltd) (product) |
|  | 8494911000001100 | Glibenclamide 5mg/5ml oral solution (Special Order) (product) |
|  | 8496711000001100 | Gliclazide 80mg/5ml oral suspension (Special Order) (product) |
|  | 8497111000001100 | Gliclazide 40mg/5ml oral suspension (Special Order) (product) |
|  | 8523811000001100 | Glibenclamide 5mg/5ml oral suspension (product) |
|  | 8523911000001100 | Glibenclamide 7.5mg/5ml oral solution (product) |
|  | 8524111000001100 | Gliclazide 160mg/5ml oral suspension (product) |
|  | 8524211000001100 | Gliclazide 40mg/5ml oral suspension (product) |
|  | 859811000001106 | Metformin 850mg tablets (Kent Pharmaceuticals Ltd) (product) |
|  | 860011000001104 | Gliclazide 80mg tablets (IVAX Pharmaceuticals UK Ltd) (product) |
|  | 8614211000001100 | Metformin 425mg/5ml oral solution (Special Order) (product) |
|  | 8615911000001100 | Metformin 500mg/5ml oral solution (Special Order) (product) |
|  | 8616711000001100 | Metformin 500mg/5ml oral suspension (Special Order) (product) |
|  | 8617611000001100 | Metformin 850mg/5ml oral solution (Special Order) (product) |
|  | 8618911000001100 | Metformin 850mg/5ml oral suspension (Special Order) (product) |
|  | 864711000001108 | Tolbutamide 500mg tablets (Alpharma Limited) (product) |
|  | 8663911000001100 | Metformin 250mg/5ml oral solution (product) |
|  | 8664011000001100 | Metformin 250mg/5ml oral suspension (product) |
|  | 8664111000001100 | Metformin 425mg/5ml oral solution (product) |
|  | 8664411000001100 | Metformin 500mg/5ml oral suspension (product) |
|  | 8664611000001100 | Metformin 850mg/5ml oral solution (product) |
|  | 8664711000001100 | Metformin 850mg/5ml oral suspension (product) |
|  | 8712011000001100 | Tolbutamide 500mg/5ml oral suspension (Special Order) (product) |
|  | 8724011000001100 | Tolbutamide 500mg/5ml oral suspension (product) |
|  | 8759611000001100 | Metformin 850mg capsules (Special Order) (product) |
|  | 8796711000001100 | Metformin 850mg capsules (product) |
|  | 889611000001103 | Metformin 500mg tablets (Approved Prescription Services) (product) |
|  | 892811000001108 | Gliclazide 80mg tablets (Approved Prescription Services) (product) |
|  | 894411000001104 | Glibenclamide 2.5mg tablets (C P Pharmaceuticals Ltd) (product) |
|  | 8990711000001100 | Glucophage SR 500mg tablets (Merck Pharmaceuticals) (product) |
|  | 918011000001106 | Glibenclamide 2.5mg tablets (Approved Prescription Services) (product) |
|  | 923911000001109 | Glibenclamide 2.5mg tablets (IVAX Pharmaceuticals UK Ltd) (product) |
|  | 925311000001109 | Gliclazide 80mg tablets (Genus Pharmaceuticals) (product) |
|  | 938211000001105 | Amaryl 2mg tablets (Aventis Pharma) (product) |
|  | 9437511000001100 | Humulin M3 Pen 100units/ml suspension for injection 3ml pre-filled pens (Eli Lilly and Company Ltd) (product) |
|  | 9528311000001100 | Apidra 100units/ml solution for injection 3ml cartridges (sanofi-aventis) (product) |
|  | 9532111000001100 | Insulin glulisine 100units/ml solution for injection 3ml cartridges (product) |
|  | 9745811000001100 | Glipizide 5mg tablets (Teva UK Ltd) (product) |
|  | 9752811000001100 | Gliclazide 80mg tablets (Milpharm Ltd) (product) |
|  | 9797311000001100 | Metformin 500mg tablets (Almus Pharmaceutical Ltd) (product) |
|  | 34043011000001100 | Fiasp FlexTouch 100units/ml solution for injection 3ml pre-filled pens (Novo Nordisk Ltd) (product) |
|  | 34043211000001100 | Fiasp Penfill 100units/ml solution for injection 3ml cartridges (Novo Nordisk Ltd) (product) |
|  | 34188211000001100 | Metformin 1g/5ml oral solution sugar free (A A H Pharmaceuticals Ltd) (product) |
|  | 34346811000001100 | Metformin 850mg tablets (Crescent Pharma Ltd) (product) |
|  | 34347311000001100 | Metformin 500mg tablets (Crescent Pharma Ltd) (product) |
|  | 34552411000001100 | Meijumet 500mg modified-release tablets (Medreich Plc) (product) |
|  | 34553311000001100 | Meijumet 1000mg modified-release tablets (Medreich Plc) (product) |
|  | 12019111000001100 | Chlorpropamide 250mg/5ml oral suspension (product) |
|  | 31014311000001100 | Eperzan 30mg powder and solvent for solution for injection pre-filled pens (GlaxoSmithKline UK Ltd) (product) |

|  | 31014611000001100 | Eperzan 50mg powder and solvent for solution for injection pre-filled pens (GlaxoSmithKline UK Ltd) (product) |
| --- | --- | --- |
|  | 31015711000001100 | Albiglutide 30mg powder and solvent for solution for injection pre-filled disposable devices (product) |
|  | 31015811000001100 | Albiglutide 50mg powder and solvent for solution for injection pre-filled disposable devices (product) |
|  | 325214004 | Product containing precisely chlorpropamide 250 milligram/1 each conventional release oral tablet (clinical drug) |
|  | 32860911000001100 | Tolazamide 250mg tablets (Imported (United States)) (product) |
|  | 32861211000001100 | Tolazamide 500mg tablets (Imported (United States)) (product) |
|  | 3472211000001100 | Insulin isophane bovine 100units/ml suspension for injection 1.5ml cartridges (product) |
|  | 3950411000001100 | Chlorpropamide 250mg tablets (The Boots Company) (product) |
|  | 4028311000001100 | Hypurin Bovine Isophane 100units/ml suspension for injection 3ml cartridges (C P Pharmaceuticals Ltd) (product) |
|  | 4033211000001100 | Insulin isophane bovine 100units/ml suspension for injection 3ml cartridges (product) |
|  | 30933411000001100 | Alogliptin 25mg tablets (Ennogen Healthcare Ltd) (product) |
|  | 32181311000001100 | Alogliptin 12.5mg / Metformin 1g tablets (Niche Pharma Ltd) (product) |
|  | 32182511000001100 | Alogliptin 12.5mg tablets (Niche Pharma Ltd) (product) |
|  | 35216811000001100 | Humalog Junior KwikPen 100units/ml solution for injection 3ml pre-filled pens (Eli Lilly and Company Ltd) (product) |
|  | 35548311000001100 | Yaltormin SR 1000mg tablets (Wockhardt UK Ltd) (product) |
|  | 35563111000001100 | Insulin lispro 100units/ml solution for injection 3ml pre-filled pen (Sanofi Pasteur) (product) |
|  | 35563711000001100 | Insulin lispro 100units/ml solution for injection 10ml vials (Sanofi Pasteur) (product) |
|  | 35776411000001100 | Insulin lispro Sanofi 100units/ml solution for injection 10ml vials (Sanofi) (product) |
|  | 35776811000001100 | Insulin lispro Sanofi 100units/ml solution for injection 3ml pre-filled pens (Sanofi) (product) |
|  | 36047311000001100 | Insulin isophane human 100units/ml suspension for injection 3ml cartridges (product) |
|  | 36047411000001100 | Insulin isophane human 100units/ml suspension for injection 3ml pre-filled disposable devices (product) |
|  | 36047511000001100 | Insulin lispro 100units/ml solution for injection 1.5ml cartridges (product) |
|  | 36047611000001100 | Insulin lispro 100units/ml solution for injection 10ml vials (product) |
|  | 36047911000001100 | Insulin protamine zinc bovine 100units/ml suspension for injection 10ml vials (product) |
|  | 36048111000001100 | Insulin soluble bovine 100units/ml solution for injection 10ml vials (product) |
|  | 36048411000001100 | Insulin soluble human 100units/ml solution for injection 3ml cartridges (product) |
|  | 36048511000001100 | Insulin soluble human 100units/ml solution for injection 3ml pre-filled disposable devices (product) |
|  | 36048611000001100 | Insulin soluble human 100units/ml solution for injection 5ml vials (product) |
|  | 36048811000001100 | Insulin zinc crystalline human 100units/ml suspension for injection 10ml vials (product) |
|  | 36804811000001100 | Jardiance 25mg tablets (Originalis B.V.) (product) |
|  | 36857411000001100 | Pioglitazone 45mg tablets (Torrent Pharma (UK) Ltd) (product) |
|  | 36889711000001100 | Januvia 25mg tablets (Pharmaram Ltd) (product) |
|  | 36889911000001100 | Januvia 50mg tablets (Pharmaram Ltd) (product) |
|  | 36893611000001100 | Trajenta 5mg tablets (Pharmaram Ltd) (product) |
|  | 36904411000001100 | Victoza 6mg/ml solution for injection 3ml pre-filled pens (Originalis B.V.) (product) |
|  | 36910511000001100 | Glydex 160mg tablets (Medreich Plc) (product) |
|  | 37120611000001100 | Metformin 850mg tablets (Mawdsley-Brooks & Company Ltd) (product) |
|  | 36618311000001100 | Suliqua 100units/ml / 50micrograms/ml solution for injection 3ml pre-filled SoloStar pens (Sanofi) (product) |
|  | 36620611000001100 | Suliqua 100units/ml / 33micrograms/ml solution for injection 3ml pre-filled SoloStar pens (Sanofi) (product) |
|  | 37337511000001100 | Amglidia 6mg/ml oral suspension with 1ml oral syringe (Amring Pharmaceuticals Ltd) (product) |
|  | 37406011000001100 | Glibenclamide 6mg/ml oral suspension sugar free (product) |
|  | 37419911000001100 | Humulin M3 100units/ml suspension for injection 10ml vials (CST Pharma Ltd) (product) |
|  | 37428211000001100 | Actos 30mg tablets (CST Pharma Ltd) (product) |
|  | 37432611000001100 | Byetta 10micrograms/0.04ml solution for injection 2.4ml pre-filled pens (CST Pharma Ltd) (product) |
|  | 37436211000001100 | Eucreas 50mg/1000mg tablets (CST Pharma Ltd) (product) |
|  | 37437511000001100 | Forxiga 10mg tablets (CST Pharma Ltd) (product) |
|  | 37437811000001100 | Forxiga 5mg tablets (CST Pharma Ltd) (product) |
|  | 37439411000001100 | Humalog Mix25 KwikPen 100units/ml suspension for injection 3ml pre-filled pens (CST Pharma Ltd) (product) |
|  | 37440011000001100 | Janumet 50mg/1000mg tablets (CST Pharma Ltd) (product) |
|  | 37440211000001100 | Januvia 100mg tablets (CST Pharma Ltd) (product) |
|  | 37440611000001100 | Januvia 50mg tablets (CST Pharma Ltd) (product) |
|  | 37449011000001100 | Metformin 500mg/5ml oral solution sugar free (DE Pharmaceuticals) (product) |
|  | 37509111000001100 | Glucophage 850mg tablets (Mawdsley-Brooks & Company Ltd) (product) |
|  | 37512311000001100 | Victoza 6mg/ml solution for injection 3ml pre-filled pens (Mawdsley-Brooks & Company Ltd) (product) |
|  | 37512811000001100 | Invokana 300mg tablets (Mawdsley-Brooks & Company Ltd) (product) |
|  | 37523211000001100 | Janumet 50mg/1000mg tablets (Mawdsley-Brooks & Company Ltd) (product) |
|  | 37525111000001100 | Januvia 25mg tablets (Mawdsley-Brooks & Company Ltd) (product) |
|  | 37526411000001100 | Jardiance 10mg tablets (Mawdsley-Brooks & Company Ltd) (product) |
|  | 37526711000001100 | Jardiance 25mg tablets (Mawdsley-Brooks & Company Ltd) (product) |
|  | 37527011000001100 | Jentadueto 2.5mg/1000mg tablets (Mawdsley-Brooks & Company Ltd) (product) |
|  | 37528111000001100 | Jentadueto 2.5mg/850mg tablets (Mawdsley-Brooks & Company Ltd) (product) |
|  | 37528511000001100 | Komboglyze 2.5mg/1000mg tablets (Mawdsley-Brooks & Company Ltd) (product) |
|  | 37536011000001100 | Trajenta 5mg tablets (CST Pharma Ltd) (product) |
|  | 37538111000001100 | Vipidia 6.25mg tablets (CST Pharma Ltd) (product) |
|  | 37550311000001100 | Onglyza 2.5mg tablets (Mawdsley-Brooks & Company Ltd) (product) |
|  | 37590811000001100 | Apidra 100units/ml solution for injection 3ml cartridges (CST Pharma Ltd) (product) |
|  | 37591211000001100 | Bydureon 2mg powder and solvent for prolonged-release suspension for injection pre-filled pens (CST Pharma Ltd) (product) |
|  | 37618711000001100 | Bilxona 60mg modified-release tablets (Accord Healthcare Ltd) (product) |
|  | 37625811000001100 | Competact 15mg/850mg tablets (Pilsco Ltd) (product) |
|  | 37636511000001100 | Eucreas 50mg/1000mg tablets (Pilsco Ltd) (product) |
|  | 37672511000001100 | Lantus 100units/ml solution for injection 3ml pre-filled SoloStar pens (Pilsco Ltd) (product) |
|  | 37694811000001100 | NovoMix 30 FlexPen 100units/ml suspension for injection 3ml pre-filled pens (Pilsco Ltd) (product) |
|  | 37695311000001100 | NovoRapid FlexPen 100units/ml solution for injection 3ml pre-filled pens (Pilsco Ltd) (product) |
|  | 37695811000001100 | NovoRapid Penfill 100units/ml solution for injection 3ml cartridges (Pilsco Ltd) (product) |
|  | 37709111000001100 | Vipidia 12.5mg tablets (Pilsco Ltd) (product) |
|  | 37716411000001100 | Byetta 10micrograms/0.04ml solution for injection 2.4ml pre-filled pens (Pilsco Ltd) (product) |
|  | 37750911000001100 | Tresiba Penfill 100units/ml solution for injection 3ml cartridges (CST Pharma Ltd) (product) |
|  | 21635011000001100 | Dapagliflozin 10mg tablets (product) |
|  | 21635211000001100 | Dapagliflozin 5mg tablets (product) |
|  | 24104411000001100 | Canagliflozin 100mg tablets (product) |
|  | 325065003 | Insulin lispro 100units/mL injection solution 10mL vial (product) |
|  | 325100008 | Insulin zinc extended release 100units/mL injection 10mL vial (product) |
|  | 325143000 | Insulin protamine zinc bovine 100units/mL injection 10mL vial (product) |
|  | 3469211000001100 | Insulin glargine 100units/ml solution for injection 3ml pre-filled disposable devices (product) |
|  | 371485002 | Insulin soluble human 100u/mL injection solution 1.5mL cartridge (product) |
|  | 37852711000001100 | Synjardy 5mg/1000mg tablets (CST Pharma Ltd) (product) |
|  | 400844000 | Insulin zinc suspension mixed human 100units/ml injection 10ml vial (product) |
|  | 400847007 | Insulin glargine 100units/mL injection solution 10mL vial (product) |
|  | 5211211000001100 | Pioglitazone 45mg tablets (product) |
|  | 5322711000001100 | Metformin 500mg / Rosiglitazone 2mg tablets (product) |
|  | 8664311000001100 | Metformin 500mg/5ml oral solution (product) |
|  | 8991411000001100 | Metformin 500mg modified-release tablets (product) |
|  | 37971811000001100 | Humalog 100units/ml solution for injection 3ml cartridges (Pharmaram Ltd) (product) |
|  | 37973011000001100 | NovoRapid FlexPen 100units/ml solution for injection 3ml pre-filled pens (Pharmaram Ltd) (product) |
|  | 37974111000001100 | Vipidia 6.25mg tablets (Pharmaram Ltd) (product) |
|  | 37989011000001100 | Eucreas 50mg/1000mg tablets (Pharmaram Ltd) (product) |
|  | 37989211000001100 | Forxiga 5mg tablets (Pharmaram Ltd) (product) |
|  | 37992511000001100 | NovoMix 30 FlexPen 100units/ml suspension for injection 3ml pre-filled pens (Pharmaram Ltd) (product) |
|  | 37992711000001100 | Onglyza 2.5mg tablets (Pharmaram Ltd) (product) |
|  | 37992911000001100 | Onglyza 5mg tablets (Pharmaram Ltd) (product) |
|  | 38018211000001100 | Lamzarin 30mg modified-release tablets (Key Pharmaceuticals Ltd) (product) |
|  | 38018511000001100 | Lamzarin 60mg modified-release tablets (Key Pharmaceuticals Ltd) (product) |
|  | 38074211000001100 | Byetta 10micrograms/0.04ml solution for injection 2.4ml pre-filled pens (Pharmaram Ltd) (product) |
|  | 38074811000001100 | Tresiba FlexTouch 200units/ml solution for injection 3ml pre-filled pens (Pharmaram Ltd) (product) |
|  | 38082811000001100 | Exenatide 2mg/0.85ml prolonged-release suspension for injection pre-filled disposable devices (product) |
|  | 38123811000001100 | Byetta 5micrograms/0.02ml solution for injection 1.2ml pre-filled pens (DE Pharmaceuticals) (product) |
|  | 38135511000001100 | Forxiga 5mg tablets (DE Pharmaceuticals) (product) |
|  | 38139511000001100 | Humulin I 100units/ml suspension for injection 3ml cartridges (DE Pharmaceuticals) (product) |
|  | 38139911000001100 | Humulin M3 100units/ml suspension for injection 10ml vials (DE Pharmaceuticals) (product) |
|  | 38141711000001100 | Januvia 25mg tablets (DE Pharmaceuticals) (product) |
|  | 38142111000001100 | Jardiance 10mg tablets (DE Pharmaceuticals) (product) |
|  | 38142411000001100 | Jardiance 25mg tablets (DE Pharmaceuticals) (product) |
|  | 38142711000001100 | Jentadueto 2.5mg/850mg tablets (DE Pharmaceuticals) (product) |
|  | 38144011000001100 | Komboglyze 2.5mg/1000mg tablets (DE Pharmaceuticals) (product) |
|  | 38144911000001100 | Lantus 100units/ml solution for injection 3ml cartridges (DE Pharmaceuticals) (product) |
|  | 38145111000001100 | Lantus 100units/ml solution for injection 10ml vials (DE Pharmaceuticals) (product) |
|  | 38145311000001100 | Lantus 100units/ml solution for injection 3ml pre-filled SoloStar pens (DE Pharmaceuticals) (product) |
|  | 38158011000001100 | Onglyza 5mg tablets (DE Pharmaceuticals) (product) |
|  | 38174611000001100 | Vipdomet 12.5mg/1000mg tablets (DE Pharmaceuticals) (product) |
|  | 38175211000001100 | Vipidia 25mg tablets (DE Pharmaceuticals) (product) |
|  | 38190911000001100 | Repaglinide 1mg tablets (Rivopharm (UK) Ltd) (product) |
|  | 38191211000001100 | Repaglinide 2mg tablets (Rivopharm (UK) Ltd) (product) |
|  | 38293311000001100 | Acarbose 100mg tablets (Rivopharm (UK) Ltd) (product) |
|  | 38530511000001100 | Metformin 500mg modified-release tablets (Morningside Healthcare Ltd) (product) |
|  | 38543111000001100 | Metformin 1g modified-release tablets (Morningside Healthcare Ltd) (product) |
|  | 1066911000000100 | Diabetes monitoring short message service text message first invitation (procedure) |
|  | 1066921000000100 | Diabetes monitoring short message service text message second invitation (procedure) |
|  | 185756006 | Diabetes monitoring first letter (procedure) |
|  | 185757002 | Diabetes monitoring second letter (procedure) |
|  | 185758007 | Diabetes monitoring third letter (procedure) |
|  | 185759004 | Diabetes monitoring verbal invite (procedure) |
|  | 310425007 | Diabetes monitoring invitation (procedure) |
|  | 1066931000000100 | Diabetes monitoring short message service text message third invitation (procedure) |
|  | 1083111000000100 | Diabetes monitoring invitation email (procedure) |
|  | 1109921000000100 | Quality and Outcomes Framework quality indicator-related care invitation (procedure) |
|  | 1110921000000100 | Quality and Outcomes Framework diabetes mellitus quality indicator-related care invitation (procedure) |
|  | 143401000000102 | Quality and Outcomes Framework diabetes mellitus quality indicator-related care invitation using preferred method of communication (procedure) |
|  | 185760009 | Diabetes monitoring telephone invite (procedure) |
|  | 705072004 | Diabetes monitoring invitation by short message service text messaging (procedure) |

**Supplementary Table 6: Counts of Covid-19 cases in the new data resource compared with the Public Health England Covid-19 reports for 31 October 2020.**

|  | **Covid laboratory test data** | | **Hospitalised participants with Covid-19** | | **Deaths with Covid-19 on the death certificate** | |
| --- | --- | --- | --- | --- | --- | --- |
|  | Definition | Number of unique  participants | Definition | Number of unique  participants | Definition | Number of unique  participants |
| **Reported by Public Health England** | *Number of people with at least one positive Covid-19 test result (either lab-reported or lateral flow device), by (i) specimen date and (ii) date reported*^9^ | (i) 904,105 (ii) 1,011,660 | *Number of people admitted to hospital who tested positive for Covid-19 in the 14 days prior to admission, and those who tested positive in hospital after admission.*^10^ | 175,555 | *Number of deaths of people whose death certificate mentioned Covid-19 as one of the causes*^11^ | 53,102^†^ |
| **Counts from the new data resource** | Number of participants with a positive antigen test from Covid laboratory tests (i) with no restrictions and (ii) in the linked cohort^‡^ | (i) 884,311 (ii) 776,503 | Number of participants with a diagnosis ICD10 code (U07.1, U07.2, U071, U072) appearing in the hospital episodes (main or secondary diagnostic code position in HES-APC) in the linked cohort^‡^ | 126,349 | Number of participants with a death registration with a mention of a diagnosis ICD10 code (U07.1, U07.2, U071, U072) in the linked cohort^‡^ | 48,333 |

^†^Reported for 30 October

^‡^Amongst 54.4 million participants, representing 96% of the population of England.
